# Software Application Profile: T-Rx: A toolbox for reproducible processing of prescriptions (Rx) stored in electronic health record databases

**DOI:** 10.1101/2025.10.07.25336002

**Authors:** Chris Wai Hang Lo, Dale Handley, Oliver Pain, Michelle Kamp, Alexandra C. Gillett, Matthew H. Iveson, Chiara Fabbri, Katherine G. Young, AMBER Research Team, Cathryn M. Lewis

**Author notes:** **Correspondence** Professor Cathryn M. Lewis, Social, Genetic and Developmental Psychiatry Centre, Institute of Psychiatry, Psychology and Neuroscience, King’s College London, Memory Lane, London, United Kingdom, SE5 8AF).

## Abstract

**Motivation:** Linkage between population-wide biobanks and electronic health records (EHRs) opens new opportunities to study the genetic and epidemiological underpinnings of treatment outcomes. However, data extraction is often complex, and the lack of open, reproducible phenotyping algorithms makes it difficult to scale analyses and compare findings across studies.

**Implementation:** T-Rx is an open-source R package consisting of three modules: (1) Extraction and Imputation Module; (2) Exposure Ascertainment Module; and (3) Phenotyping Module.

**General features:** T-Rx allows the extraction and imputation of prescription details, such as product strength and prescribed quantity in UK-based EHR databases, including UK Biobank and CPRD. The Exposure Ascertainment Module converts discrete prescribing events into longitudinal exposure periods in one-line R commands. The Phenotyping Module inputs prescriptions and returns phenotype dataframes for downstream analyses.

**Availability:** T-Rx is available as a Github Repository at [https://github.com/chrislowh/T-Rx]. Documentation is available at: [https://chrislowh.github.io/T-Rx/].

## Introduction

Since the computerisation of healthcare records and electronic prescribing, healthcare systems worldwide have established extensive electronic health record (EHR) database initiatives ^1^. These EHR databases contain clinical information, which often comes as structured data (such as diagnoses and medications) or as unstructured free-text information in clinical notes. To enhance health data research, EHR databases have established data linkage to population-wide biobanks which include genetic and self-reported variables ^2^. This offers a rich combination of clinical and genetic data and opens new opportunities to study individual differences in treatment outcomes.

However, there are still major barriers for widespread use of prescribing information in EHR-linked biobanks. Data infrastructure varies across EHR databases, and full details of dosage, quantity, and frequency of administration might not be structured in readily usable formats ^3^, if available at all. Prescribing events are usually discrete and cross-sectional, thus not readily available for researchers to infer treatment episodes or perform longitudinal analysis based on medication exposure ^4^.

In EHR analyses, proxy measures attempt to capture treatment trajectories and outcomes, as symptom changes are often not tracked longitudinally in structured records ^5^. Treatment-resistant depression (TRD), for instance, can be defined from EHRs as at least two antidepressant switches during an episode of depression ^6^. This working definition enabled large-scale studies of non-response to antidepressants, overcoming the sample size limitations of clinical trials ^6^. However, these treatment outcomes are not harmonised across studies ^5,7^, making cross-validation of results challenging. Open-source platforms like the Health Data Research UK Phenotype Library ^8^ host reproducible phenotyping algorithms for diseases, but we lack a similar repository for standardisation of treatment outcomes. These are necessary to conduct scalable and reproducible analyses, and allow cross-validation of results across studies ^9,10^.

Here, we detail the features and applications of T-Rx, demonstrating its utility across EHRs and clinical areas, including psychiatric disorders and cardiometabolic diseases.

## Implementation

### Overview

T-Rx is available as an open-source R package (https://github.com/chrislowh/T-Rx), developed under R version 4.3.1 ^11^. The package is structured into three modules by functionality:

1. **Extraction and Imputation Module:** streamlining data extraction and wrangling processes from structured prescription data to ready-to-analyse prescriptions;
2. **Exposure Ascertainment Module:** aggregating cross-sectional prescribing events into longitudinal exposure episodes; and
3. **Phenotyping Module:** a library of reproducible algorithms for creating phenotypes related to treatment exposure and response.

**Figure 1A** gives an overview of T-Rx functionalities.

**Figure 1.**
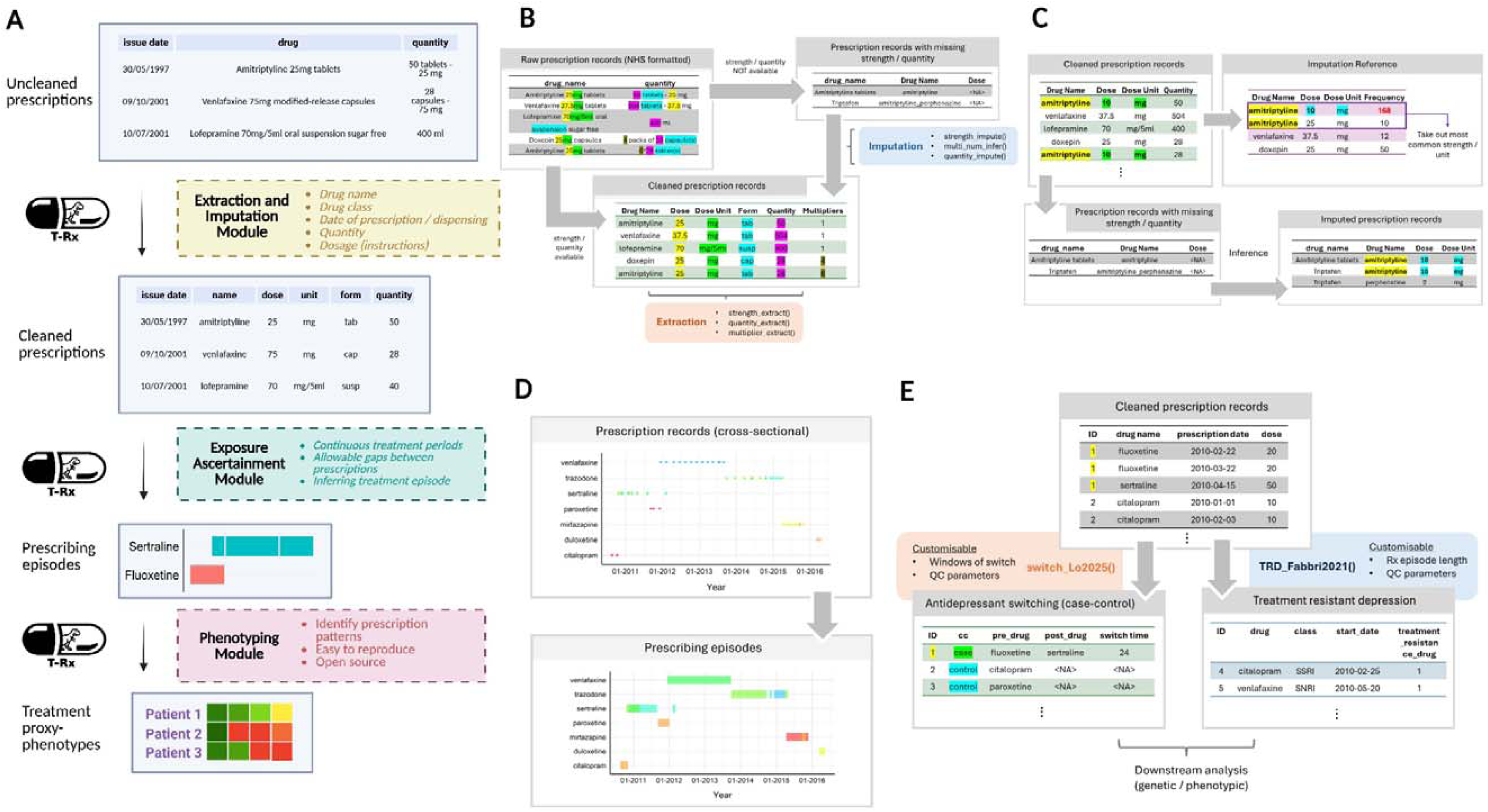
(A) Overview of T-Rx modules; (B) Extraction and Imputation Module of T-Rx, with UK Biobank primary care prescription records as an example; (C) Imputation of strengths and quantities with T-Rx; (D) Exposure Ascertainment Module of T-Rx; (E) Phenotyping Module of T-Rx, using antidepressant switching and treatment resistant depression as example functions, showing example prescriptions as input and phenotype data frames as outputs **Legends** Figures 1B/C demonstrate the processes of extracting and imputing product strengths from prescription strings, using UK Biobank primary care records as example. This can also be applied to other electronic health records, where dosage (i.e, the actual amount of medication taken) instead of product strengths are available. Created in BioRender. Lo, C. (2025) https://BioRender.com/2izbwra. **Abbreviations** QC = quality control; Rx = prescription.

### Installation

T-Rx can be installed as an R package cloned from the T-Rx GitHub repository, alongside website documentation. Details of installation and package dependencies are at https://chrislowh.github.io/T-Rx/installation.html, applicable to local machines and high-performance computing environments.

## Extraction and Imputation Module

### Extraction of prescription details

The extraction functions rely on regular expression (regex) patterns to extract strengths and quantities of medications in prescriptions, using strength units (e.g., *mg*, *mg/ml*) and dosage forms (e.g., *tab*, *suspension*) as *tags* for extraction (**Figure 1B**). This allows flexible capture of strings describing pharmaceutical products, and is not limited to specific coding systems in prescriptions. These functions include:

- *‘strength_extract()’*: extract strengths (highlighted in yellow in **Figure 1B**) as numbers that precede the user-input strength units, e.g., *mg* (in green);
- *‘quantity_extract()’:* extract quantities, e.g., number of tablets (in pink) as numbers that precede the user-input dosage forms, e.g., *tablets* (in blue); and
- *‘multiplier_extract()’:* extract multiplier information whenever necessary, e.g., *number of packs* (in grey).

### Imputation of prescription details

The imputation functions infer strength and quantity information from T-Rx extraction outputs or direct user inputs. A flowchart for imputation is described in **Figure 1C**. The two imputation functions are:

- ‘*strength_impute()*’: performs imputation on product strengths, and expands multi-strength products into multiple rows with strengths of each active ingredient mapped;
- ‘*quantity_impute()*’: performs imputation on prescribed quantities.

The validity of extraction and imputation functions were assessed using oral hypoglycaemic agent prescriptions in Clinical Practice Research Datalink (CPRD) Aurum, and antidepressant prescriptions in UK Biobank (UKB). Further details of technical documentation and validation are described in **Supplementary Methods**.

### Additional functionalities for inferring prescription information

EHR linkage was established with multiple providers in UKB ^12^, making the format of prescriptions particularly heterogeneous. Thus, further inference is required to gather the strength and quantity information. *‘multi_num_infer()’ and ‘duration_handling()’* are additional functionalities designed to handle the complexity of UKB prescription records (see **Supplementary Methods**).

## Exposure Ascertainment Module

Prescriptions are usually provided as a single row for each prescribing event, which need to be merged into treatment episodes (**Figure 1D**). One common approach is to assign two prescriptions into one treatment episode if the periods of coverage overlap (**Supplementary figure 4A**). A window between prescribing periods should be permitted to account for real-world treatment complexities, such as nudges in follow-up periods and medication stockpiling ^13^. When the duration of exposure is unavailable, assumptions of daily defined doses and prescription quantities are needed to infer treatment episodes **(Supplementary figure S4B)** ^14^.

T-Rx provides two algorithms to construct treatment episodes:

1. *‘rx_merge()’*: when the start and end dates (coverage) of each prescription are available, aggregate them into the same episode if the coverages overlap (**Supplementary figure S4D and S4E**);
2. *‘rx_infer()’*: when coverage of each prescription is not available, infer treatment duration and episodes by “repeated prescriptions” (**Supplementary figure S4B**).

Users can adjust function parameters, such as gaps allowed between prescriptions and default prescription duration, depending on the dataset and research question. After merging, the function returns a dataframe with prescribing episodes **(Supplementary figure S4F and S4G)**. Details of the functions are available in **Supplementary Materials.**

## Phenotyping Module

The Phenotyping Module hosts a library of algorithms to create proxy treatment outcomes, such as drug switching or treatment resistance. Using prescriptions as input, users can create treatment-related proxy phenotypes in one-line R commands with optional parameters (**Figure 2**). To contribute to open science, researchers can submit code for new phenotyping algorithms to T-Rx, enabling others to apply algorithms reproducibly.

**Figure 2.**
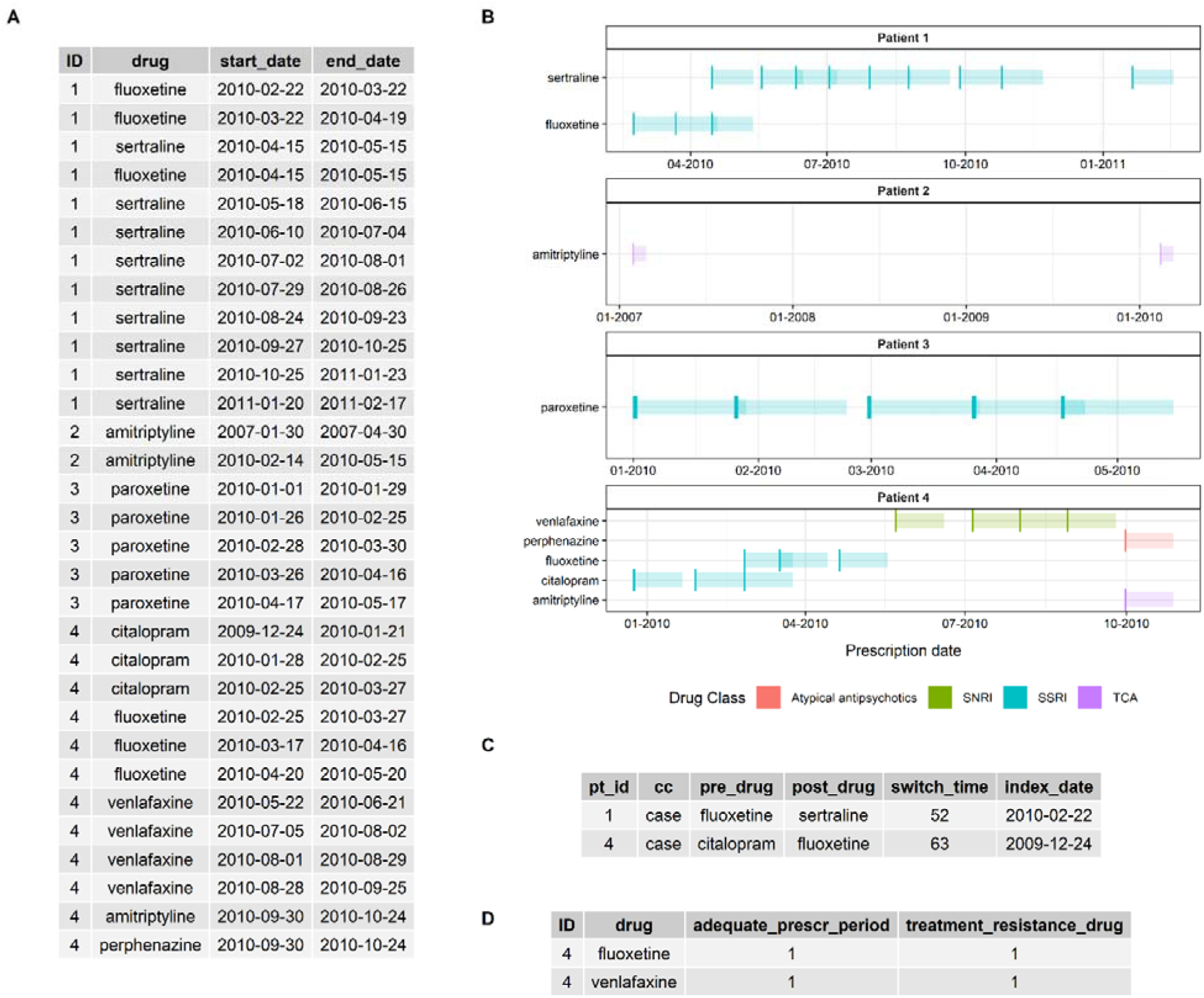
Sample use of T-Rx phenotyping functions, with (A) prescription data frame as user input, (B) visualisation of prescriptions from input, (C) output phenotype data frame with drug switching (switch_Lo2025()) and (D) output phenotype data frame with treatment resistant depression (TRD_Fabbri2021()). **Legends** Details of functions creating the two phenotypes are at: https://chrislowh.github.io/T-Rx/.

## Usage of T-Rx

### Datasets for illustration and validation

#### UK Biobank (UKB)

UKB is a prospective health study that recruited around 500,000 participants, with linkage to primary care prescriptions established for ∼230,000 participants ^12^. We illustrate the Extraction and Imputation Module using uncleaned antidepressant, antipsychotic and lithium prescriptions in UKB (extracted from codes in previous research ^6,12^, and **Supplementary table S1**).

### Clinical Practice Research Datalink (CPRD) Aurum

CPRD Aurum is a UK EHR database containing structured EHR data from primary care practices ^15^. The performance of T-Rx in strength extraction was tested using oral hypoglycemic agent and statin prescriptions in a randomly selected sample of participants with type 2 diabetes ^16^ (full details in **Supplementary Materials)**.

## Use Case 1: Product strength extraction

The strength extraction function ‘*strength_extract()*’utilises the strength units as “tags”, and the numbers preceding the “tags” are extracted as product strength (**Figure 1B**). Strings specifying strength units for both solid (e.g., *tablets, capsules*) and liquid (e.g., *injections, suspensions*) dosage forms need to be given as arguments (*liquid_strength_unit, solid_strength_unit)* based on prior knowledge of the medication(s) of interest. Example code for running ‘*strength_extract()*’ is shown below, for antidepressant, lithium and antipsychotic prescriptions in UKB.

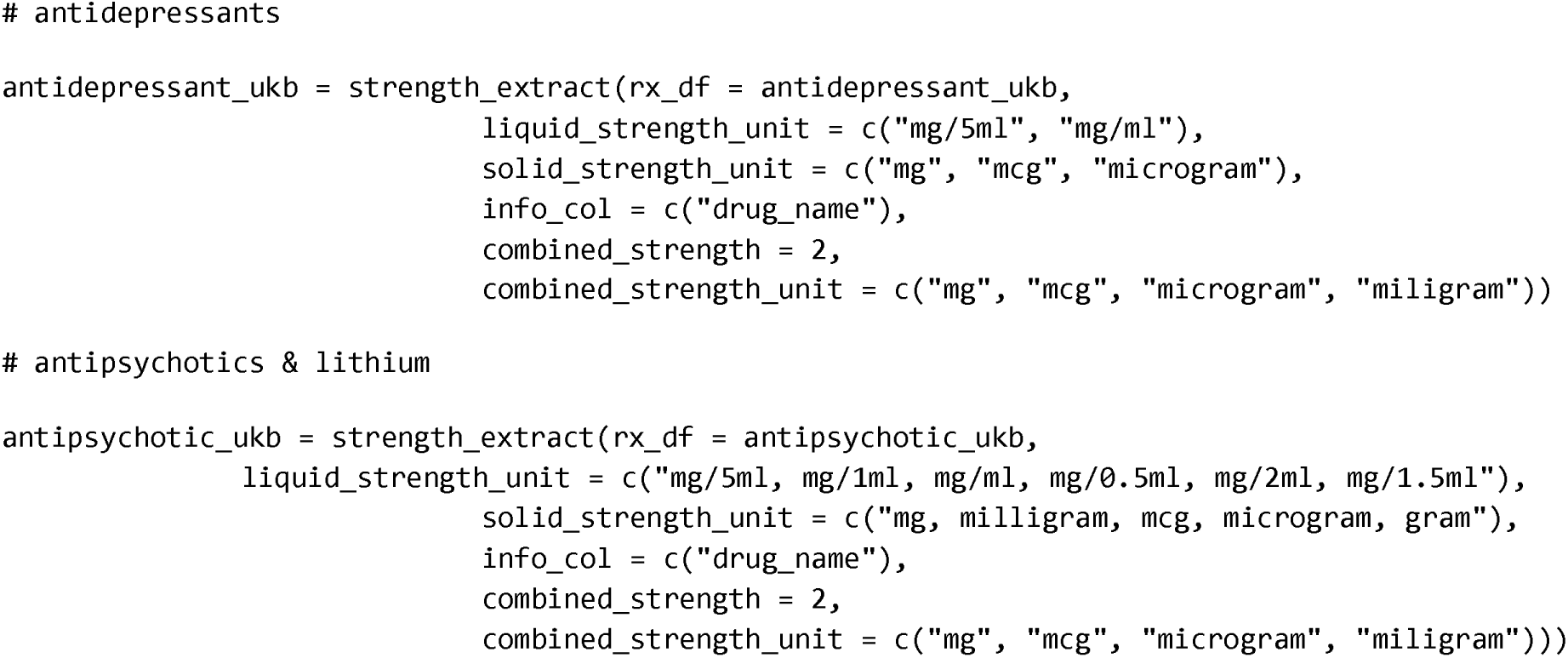

Running ‘*strength_extract()*’ returns a prescription dataframe with strengths of products extracted (**Figure 1B**). In UKB, the strengths of 2,320,226 (85.3%) antidepressant and 349,705 (81.2%) antipsychotic and lithium prescriptions were extracted (**Supplementary figure S5, table S9**). The missingness was due to strength and strength unit values being absent from prescriptions; these can be imputed using ‘*strength_impute()*’ (**Figure 1C**), providing strength values for over 99% of prescriptions (**Supplementary table S8**).

In CPRD Aurum, T-Rx extracted product strengths for prescriptions with information available, 98.6% of which matched dm+d outputs generated from CPRD Aurum (8,586,964/8,707,546). Product strengths of statins can also be extracted for 99.9% of prescriptions in the same sample, with further details described in **Supplementary Methods**. Log files with parameters and summary-level measures are provided in R to ensure reproducibility (**Supplementary Methods**).

## Use Case 2: Converting prescriptions into treatment-related phenotypes

We provide prescribing journeys for four synthetic patients on antidepressants, mimicking real-world prescription dataframes (**Figure 2**). From here, drug switching and TRD events can be created in one-line R commands, based on open-source codes in the literature (*‘switch_Lo2025()’* ^17^, and*‘TRD_Fabbri2021()’* ^6^). To run the phenotyping function ‘*switch_Lo2025()*’ (**Figure 1E**), users specify the prescription data frame, the column names for drug and prescription dates, windows to capture switching events and quality control parameters. These parameters allow users to adjust the specificity and sensitivity in capturing those who switch drug. Running ‘*switch_Lo2025()*’ returns a data frame of switchers (as wide format, **Figure 2C**), containing drug information before and after switching, the time to switch and index date, i.e., the first prescription date of the pre-switch drug. These inputs can used by ‘*TRD_Fabbri2021()*’, which returns a dataframe containing TRD patients, with details on antidepressant prescriptions received (**Figure 2D**). A schematic diagram of the process, including prescription data required, visualisations of output dataframes after running the functions are given in **Figure 2**.

## Discussion

To our knowledge, T-Rx is the most comprehensive tool to streamline standardisation of prescription data, to give ready-to-analyse outputs of medication exposure episodes and phenotypes capturing treatment and response patterns. T-Rx aims to facilitate analysis of treatment response by researchers who lack expertise in EHR data processing by providing a user-friendly and reproducible framework to create these datasets. This increases the usability of health data resources, enabling a broader range of researchers to work with prescription data to study treatment patterns and response.

The strength extraction and imputation functions in T-Rx make prescription dosages available in population-wide biobanks. This would facilitate future pharmacokinetic ^18^ and pharmacogenomic studies ^19^ for personalising medication dosing. T-Rx also hosts open-source algorithms to define treatment outcomes as simple R commands. These workflows are typically undertaken in pharmaco-epidemiological studies, which often require complex coding to assess prescriptions longitudinally instead of as discrete events, as well as quality control parameters imposed to ensure clinical relevance ^6,17^. The codes for clinical constructs such as TRD ^6,20^ often differ across studies and are rarely shared, hampering reproducibility. The absence of open-source platforms to host treatment outcome algorithms has made cross-study comparisons and validation challenging ^10^. We hope T-Rx will address such challenges and allow harmonisation of treatment definitions in real-world data ^10^.

Compared to T-Rx, most existing tools working with prescription data focus on a single purpose, such as estimating drug exposure duration ^21^ or visualising prescription data ^22–24^. T-Rx utilises regex patterns for the extraction of prescription details in contrast to natural language processing (NLP) workflows which focus on named-entity recognition tasks. These show excellent performance by mapping drug names to active pharmaceutical ingredients, but are less satisfactory for extracting prescribed dosages ^23^. NLP algorithms are also computationally intensive, which could be a barrier in resource-limited settings. Rule-based algorithms for extracting prescription details in T-Rx can also serve as a performance benchmark for the development of NLP algorithms.

T-Rx is currently limited by the scope of validation using prescription records across healthcare systems. In extraction functions, T-Rx demonstrated satisfactory performance in UK EHR health data resources (UKB, CPRD) but has not been evaluated in non-UK EHR databases, some of which provide readily structured data. Collaborative efforts from the research community can expand T-Rx functionalities beyond current phenotypes, so that new approaches for making inferences on prescription data can be developed, including medication non-adherence ^25^. With the growing use of prescribing data in healthcare research, T-Rx offers a framework for prescription processing that is adaptable across EHR datasets, enabling researchers to apply relevant modules to prepare the prescription data for analysis.

## Ethics approval

The UK Biobank has research ethics approval from the North West Multi-centre Research Ethics Committee (MREC; approval number 11/NW/0382) covering the United Kingdom. This current article has received approval to be conducted using the UK Biobank resource, under the project application number 82087. Ethical approval for observational studies using the Clinical Practice Research Datalink is granted by the NHS Health Research Authority (Derby Research Ethics Committee; REC reference 21/EM/0265). Individual patient consent is not required.

## Notes

AMBER Research Team: Mark J Adams (Institute for Neuroscience and Cardiovascular Research, University of Edinburgh, Edinburgh, UK), Clara Albiñana (Department of Psychiatry, University of Oxford, Oxford, UK), Iona Beange (Usher Institute, University of Edinburgh, Edinburgh, UK), Megan Calnan (Institute for Neuroscience and Cardiovascular Research, University of Edinburgh, Edinburgh, UK), Arlene Casey (Usher Institute, University of Edinburgh, Edinburgh, UK), Cristina Douglas (Usher Institute, University of Edinburgh, Edinburgh, UK), Matúš Falis (Institute for Neuroscience and Cardiovascular Research, University of Edinburgh, Edinburgh, UK; Usher Institute, University of Edinburgh, Edinburgh, UK), Sue Fletcher-Watson (Usher Institute, University of Edinburgh, Edinburgh, UK; Salvesen Mindroom Research Centre, University of Edinburgh, Edinburgh, UK), Anjali Henders (Institute for Molecular Bioscience, University of Queensland, Brisbane, QLD, Australia), Andrew M McIntosh (Institute for Neuroscience and Cardiovascular Research, University of Edinburgh, Edinburgh, UK; Centre for Genomic & Experimental Medicine, Institute for Genetics and Cancer, University of Edinburgh, Edinburgh, UK), Renata Nedel Pertile (Institute for Molecular Bioscience, University of Queensland, Brisbane, QLD, Australia), Quan Nguyen (Institute for Molecular Bioscience, University of Queensland, Brisbane, QLD, Australia), Emer O’Leary (Usher Institute, University of Edinburgh, Edinburgh, UK), Sonia Shah (Institute for Molecular Bioscience, University of Queensland, Brisbane, QLD, Australia), Madhurbain Singh (Social, Genetic and Developmental Psychiatry Centre, King’s College London, London, UK), Mark Somerville (Usher Institute, University of Edinburgh, Edinburgh, UK), Kate Stewart (Social, Genetic and Developmental Psychiatry Centre, King’s College London, London, UK), Vanessa Tsui (Institute for Molecular Bioscience, University of Queensland, Brisbane, QLD, Australia), Alicia Walker (Department of Psychiatry, University of Oxford, Oxford, UK), Heather C Whalley (Institute for Neuroscience and Cardiovascular Research, University of Edinburgh, Edinburgh, UK), Naomi R Wray (Department of Psychiatry, University of Oxford, Oxford, UK; Institute for Molecular Bioscience, University of Queensland, Brisbane, QLD, Australia; Queensland Brain Institute, University of Queensland, Brisbane, QLD, Australia).

## Acknowledgements

For the purposes of open access, the author has applied a Creative Commons Attribution (CC BY) licence to any Accepted Author Manuscript version arising from this submission. We extend our gratitude to the contributions of other investigators on the AMBER: Antidepressant Medications: Biology, Exposure & Response project, with the list of investigators provided in **Supplementary Materials**. A previous version of this article was published as a preprint on medRxiv: https://doi.org/10.1101/2025.10.07.25336002.

## Author Contributions

Chris Wai Hang Lo (Conceptualization, Data curation, Formal analysis, Methodology, Resources, Software, Validation, Visualization, Writing – original draft, Writing – review & editing), Dale Handley (Data curation, Formal analysis, Methodology, Validation, Writing – original draft, Writing – review & editing), Oliver Pain (Resources, Software, Writing – review & editing), Michelle Kamp (Methodology, Visualization, Writing – review & editing), Alexandra C. Gillett (Data curation, Methodology, Resources, Writing – review & editing), Matthew H. Iveson (Conceptualization, Methodology, Writing – review & editing), Chiara Fabbri (Data curation, Writing – review & editing), Katherine G. Young (Data curation, Resources, Writing – review & editing), Cathryn M. Lewis (Conceptualization, Funding acquisition, Methodology, Project administration, Resources, Supervision, Writing – original draft, Writing – review & editing).

## Supplementary data

Supplementary data are available at *IJE* online.

## Data availability

The data supporting this article is openly available from the King’s College London research data repository, KORDS, at https://doi.org/10.18742/33103934. Tutorials and instructions for T-Rx are provided at https://chrislowh.github.io/T-Rx/. The UK Biobank data was accessed via project 82087. Data access to UK Biobank is available for approved researchers at https://www.ukbiobank.ac.uk/enable-your-research. This project was conducted under CPRD study reference ID 23_002544. Researchers wishing to access CPRD data must obtain the necessary approvals and data access agreements.

## Use of Artificial intelligence (AI) tools

Artificial intelligence tools were used in editing R documentations for T-Rx functionalities to improve readability of technical details to users, as well as website development for T-Rx package descriptions.

