## Supplementary Materials for "Software Application Profile: T-Rx: A toolbox for reproducible processing of prescriptions (Rx) stored in electronic health record databases"

Chris Wai Hang Lo, BPharm^1^, Dale Handley, PhD^1^, Oliver Pain, PhD^2^, Michelle Kamp, PhD^1^, Alexandra C. Gillett, PhD^1,3^, Matthew H. Iveson, PhD^4^, Chiara Fabbri, PhD^5^, Katherine G. Young, PhD^6^, Cathryn M. Lewis, PhD^1,3,7*^, AMBER Research Team

^1^ Social, Genetic & Developmental Psychiatry Centre, Institute of Psychiatry, Psychology and Neuroscience, King's College London, London, United Kingdom

^2^ Department of Basic and Clinical Neuroscience, Institute of Psychiatry, Psychology and Neuroscience, King’s College London, London, United Kingdom

^3^ National Institute for Health Research Maudsley Biomedical Research Centre at South London and Maudsley NHS Foundation Trust and King’s College London, London, United Kingdom

^4^ School of Neurological and Cardiovascular Sciences, University of Edinburgh, Edinburgh, United Kingdom

^5^ Department of Biomedical and Neuromotor Sciences, University of Bologna, Bologna, Italy

^6^ Department of Clinical and Biomedical Sciences, University of Exeter, Exeter, United Kingdom

^7^ Department of Medical & Molecular Genetics, King’s College London, London, United Kingdom

**Software Application Profile: T-Rx: A toolbox for reproducible processing of prescriptions (Rx) from electronic health record databases**

[Supplementary figure S4. Exposure ascertainment module functionality for a three-prescription example: (A) sertraline prescriptions (with start and end dates of prescriptions available); (B) sertraline prescriptions (without end dates of prescriptions); (C) sertraline prescriptions (as table); (D) treatment episodes when sertraline prescriptions were merged into prescribing episodes, without gaps allowed between prescriptions; (E) treatment episodes when sertraline prescriptions were merged into prescribing episodes, with a 7-day gap allowed between prescriptions; (F), (G) treatment episodes as tables from (D) and (E), as outputs from *rx_merge()*.	83](#_Toc237218554)

**Supplementary Methods**

**Study Samples used in T-Rx development and validation**

**UK Biobank**

**Sample Description**

The UKB is a population-wide prospective cohort study in the United Kingdom (UK) recruiting around 500,000 participants, collecting genetic, demographic and self-assessed information at enrolment ^1^. All participants provided consent upon assessment, and completed self-assessment on health ^1^. Several waves of follow-up assessments were conducted after initial assessment, including physical and mental health measurements in the form of mental health questionnaire ^2^.

**Primary Care Records**

According to documentation from UKB, there is currently no national system for collecting primary care records ^3^. Therefore, at the current release, UKB liaised with data suppliers and other intermediaries to establish linkage to primary care data for ~230,000 (~45%) UKB participants, all of whom have provided written consent for linkage to their health-related records.

Prescriptions in UKB primary care records are available from the 1990s to 2018, with the start and end dates dependent on the databases linked to specific regions of practice. Only the dates of prescription are available with no information on dispensing. These prescriptions were recorded under READ v2, British National Formulary (BNF) and the NHS Dictionary of Medicines and Devices (dm+d) codes. Details of coding systems, including READ codes, BNF and dm+d codes are available in documentations from UKB ^3^ and previous work on treatment-resistant depression ^4^. Details of the data providers and the coding schema used are summarised below.

| **Country** | **GP Computer System Supplier** | **Approx no. of UK Biobank participants** | **Clinical coding classification** | **Prescription coding** |
| --- | --- | --- | --- | --- |
| Scotland ^a^ | EMIS ^b^ / Vision ^c^ | 27,000 | - Read v2 | - Read v2 - British National Formulary (BNF) |
| Wales ^d^ | EMIS / Vision | 21,000 | - Read v2 | - Read v2 |
| England | TPP^e^ | 165,000 | - Clinical Terms Version 3 (CTV3 or Read v3) | - BNF |
|  | Vision | 18,000 | - Read v2 | - Read v2 - Dictionary of - Medicines and Devices (dm+d) |

^a^ UK Biobank has engaged Albasoft (http://www.albasoft.co.uk/) (a third party data processor) to obtain data from GP practices in Scotland.

^b^ EMIS Health (https://www.emishealth.com/) is a computer system supplier to the NHS and provides the EMIS Web practice management system.

^c^ Vision Health (https://www.visionhealth.co.uk/) (previously InPS) is a computer system supplier and provides the Vision practice management system.

^d^ Data from Wales have been obtained via the SAIL Databank (https://saildatabank.com/) hosted by the University of Swansea.

^e^ TPP (https://www.tpp-uk.com/) is a computer system supplier and provides the SystmOne practice management system.

##### **Primary sample**

Antidepressant prescriptions in UKB primary care records are used as the primary sample for T-Rx function development in the **Extraction and Imputation Module** (see below), with the performance further evaluated using lithium and antipsychotic prescriptions in UKB. These drugs were selected for their complexity in prescription strings within the dataset, with the expectation that the functions should be applicable to most EHR databases. The results for performance of strength extraction and imputation functions are described in the main text, and in **Performance evaluation of strength extraction** functions in this Supplement (see below). Sample primary care prescriptions in UKB are illustrated in **Supplementary figure S1**, to illustrate the formats of prescription strings in EHR**.**

**Clinical Practice Research Datalink (CPRD) Aurum**

**Sample Description**

CPRD is a nationwide database of primary care medical records from patients in the United Kingdom starting from 1987 ^5–7^. CPRD collects anonymised data on diagnoses and treatment (as prescriptions) from participating general practices on a monthly basis, with further data linkage established to make demographic and hospital episode statistics available ^5^. CPRD Aurum provides comprehensive records for clinical events, demographic information, blood biochemistry, and prescription linkage ^6^. As of 2019, over 16 million individuals were registered in CPRD Aurum ^6^.

**Validation Sample**

All individuals with at least one Type 2 diabetes (T2D) code between the start of the study (1988) and 31/12/2025 were included in the initial CPRD Aurum data extract (n = 2,969,156) ^7^. A full description of the cohort extraction process is available online (<https://github.com/Exeter-Diabetes/CPRD-Cohort-scripts>). Oral hypoglycemic agent (OHA) and statin prescription records ^8^ were extracted from CPRD Aurum (**Supplementary tables S2 to S4**). These link to a product dictionary containing dm+d derived information, including product name and strength, where available.

To test for the validity of T-Rx in strength extraction in CPRD, we randomly selected OHA prescriptions in 100,000 individuals from the sample described above, where T-Rx extraction outputs were assessed for concordance with strengths obtained from linked dm+d codes (**Supplementary tables S2 and S3**). As additional validation, 847,007 statin prescriptions were extracted from randomly selected 10,000 individuals in the same validation sample (**Supplementary table S4**). Samples of CPRD prescriptions are presented in **Supplementary figure S2.** The numbers of CPRD participants and prescriptions summarized in **Supplementary tables S5 and S6**, and **Supplementary figure S3**.

**Extraction and Imputation Module**

#### **Extraction and imputation functions for product strengths**

Using antidepressant and antipsychotic/lithium prescriptions as examples, the code chunks below provide sample parameters to extract strengths of products from prescriptions using `*strength_extract()`*.

```
# antidepressants
antidepressant_ukb = strength_extract(rx_df = antidepressant_ukb,
				liquid_strength_unit = c("mg/5ml", "mg/ml"),
				solid_strength_unit = c("mg", "mcg", "microgram"),
				info_col = c("drug_name"),
				combined_strength = 2,
				combined_strength_unit = c("mg", "mcg", "microgram", "miligram"))
# antipsychotics
antipsychotic_ukb = strength_extract(rx_df = antipsychotic_ukb,
				liquid_strength_unit = c("mg/5ml, mg/1ml, mg/ml, mg/0.5ml, mg/2ml, mg/1.5ml"),
				solid_strength_unit = c("mg, milligram, mcg, microgram, gram"),
				info_col = c("drug_name"),
				combined_strength = 2,
				combined_strength_unit = c("mg", "mcg", "microgram", "miligram")))
```

Lists of strings specifying strength units for both solid (e.g., *tablets, capsules*) and liquid (e.g., *injections, suspensions*) dosage forms need to be given as arguments (*liquid_strength_unit, solid_strength_unit)* in the function. Here, we provided example lists of strength units which were evaluated for performance in main text, limited to specific therapeutic classes.

Running the `*strength_extract()*` functions returns a prescription dataframe with strengths of products extracted, with log messages to ensure reproducibility. An example of log message for *`strength_extract()`* is shown below, using a random sample of 5000 antidepressant prescriptions in UKB.

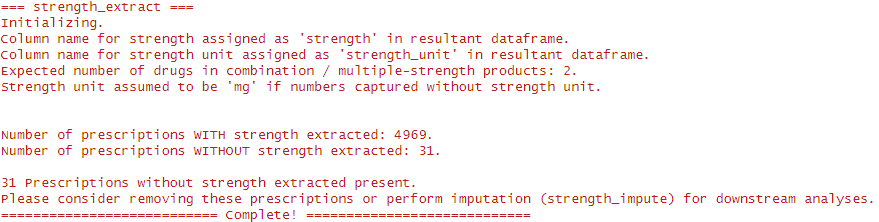

The log file prints out the parameters users specified, such as the column names of output produced (*`strength`*, *`strength_unit`*), number of drugs in multi-strength products expected, and assumptions for strength units in the prescriptions without strength units detected. Summary-level measures, such as number of prescriptions with or without strength information extracted are also reported.

Output dataframes from `*strength_extract()*` can be passed into `*strength_impute()*` for imputation, where the strengths of products cannot be extracted by strength unit tags.

```
# antidepressants
antidepressant_ukb = strength_impute(rx_df = antidepressant_ukb,
					    ref_drug_col = "chem_name",
					    strength_colname = "strength", 							    	    strength_unit_colname = "strength_unit")
# antipsychotics
antipsychotic_ukb = strength_impute(rx_df = antipsychotic_ukb,
					    ref_drug_col = "chem_name",
					    strength_colname = "strength", 							    	    strength_unit_colname = "strength_unit")
```

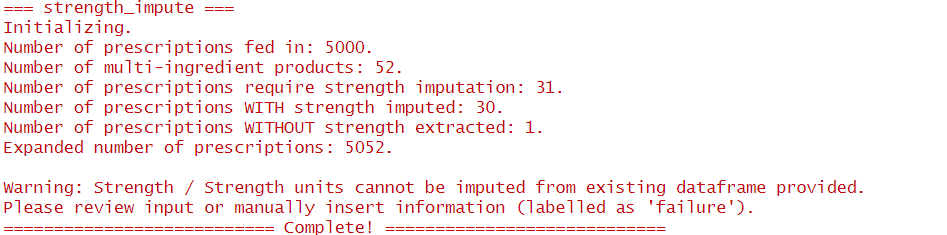
An example of log message for `*strength_impute()*` is shown below, using the same random sample of 5000 antidepressant prescriptions from above, after running `*strength_extract()*`.

The log message prints out summary-level measures, such as number of prescriptions in the input dataframe, number of prescriptions with multiple ingredients detected and number of prescriptions with strengths imputed. For prescriptions that cannot be expanded or imputed using `*strength_impute()*`, they would also be printed out as part of the log message and labelled as `*failure*` in the resultant dataframe. This can be due to inconsistent strength units or inability of the function to map correct strengths based on drug names (`*ref_drug_col*`), and would require manual input or quality control in data cleaning.

**Performance evaluation of strength extraction functions**

In UKB, 2,718,545 antidepressant prescriptions were available for 82,647 participants, and 430,705 antipsychotic and lithium prescriptions were available for 38,645 participants. **Supplementary table S7** and **Supplementary figure S6** describe the summary measures by each drug available in UKB. The performance of `*strength_extract()*` and `*strength_impute()*` are described in main text, with the strength distributions of products after running T-Rx are summarized in **Supplementary figure S5**, and **Supplementary tables S9 and S10**.

For oral antidiabetics in CPRD Aurum, product strengths were available for 8,707,513 of 8,707,546 prescriptions, all of which can also be extracted with T-Rx (**Supplementary figure S6**). Extraction outputs from T-Rx agreed with dm+d values for 98.6% of prescriptions (N = 8,586,964). Discrepancies between T-Rx and dm+d outputs were due to differences in rounding for concentrations of liquid dosage formulations (46,076; 0.5%), or differences in string formatting in multi-strength products (74,473; 0.9%). We provided multi-strength product codes in **Supplementary table S11**, showing the outputs from CPRD Aurum and T-Rx for comparison. For statins, product strengths were extracted by T-Rx in 99.9% (845,745/847,007) of prescriptions available in the validation sample (**Supplementary figure S6**).

#### **Validation of strength imputation functions**

Antidepressant prescriptions in UKB are used to evaluate the validity of `*strength_impute()*` in imputing product strengths, as a simulation for cases of poor data quality. Only the 2,320,226 (see main text) prescriptions in which the product strength information are complete and extracted by `*strength_extract()*` are used as comparison to imputation outputs.

Cross-validation was performed by randomly sampling different proportions of prescriptions (see below) as training dataset, with the remainder being testing dataset. The strengths and strength units for testing datasets were masked and imputed using `*strength_impute()*`, then matched to the respective values/strings extracted from `*strength_extract()*`.

For cross-validation, prescriptions are divided using training/testing proportions of 70%/30%, 90%/10% and 95%/5% respectively, to evaluate the stability of mode imputation in different scenarios of data quality, i.e.: missingness. Accuracy is assessed as the proportions of prescriptions with product strength values exactly matched to the strengths extracted, and imputation rate is assessed as the proportion of rows that can be imputed out of all masked prescriptions in the testing dataset. This process is done under the ‘boot’ R package version 1.3-32 ^9^. The cross-validation process is repeated for 100 (and/or 250) times to yield distributions of accuracy and imputations rates to estimate 95% confidence intervals for the respective metrics in the “boot” R package ^9^.

#### **Additional functions for primary care records in UKB**

**Extraction of multipliers**

In UKB primary care records, information on multipliers is sometimes included as part of prescription details. An example prescription is shown below, using antidepressant prescriptions. Information on multipliers was highlighted in blue.

| **drug_name** | **quantity** |
| --- | --- |
| Fluoxetine 20mg capsules (Teva UK Ltd) | 3 packs of 30 capsule(s) |
| Venlafaxine 37.5mg tablets | 2*56 tablets |
| Sertraline 50mg tablets (Sandoz Ltd) | 2 packs of 28 tablet(s) |

`*multiplier_extract()*` function in T-Rx requires users to specify strings expected to represent multipliers (e.g.: *pack*, *pack of*) and specified in the *multipliers* argument in the function. The function then recognizes these strings and extracts the numbers preceding this string, similar to `*strength_extract()`* and `*quantity_extract()`*. A sample code for `*multiplier_extract()*` is shown below.

```
# multiplier extraction
antidepressant_ukb = multiplier_extract(rx_df = antidepressant_ukb,
					multipliers = c("pack", "pack of" , "\\*"),
					alt_multipliers = c("x"),
					info_col = "quantity", 							    	       qc_remove = FALSE)
```

On the above function, the possible strings representing multipliers are specified by the `*multiplier*` argument in the function. We also further provide `*alt_multipliers*` as a more non-specific or loose search of multiplier information, expecting to capture strings such as “2x56” and “3x30”. Users have the option to not use these arguments depending on the data structure of the prescription records involved. If numeric values cannot be searched from these multiplier strings, *999* will be assigned for users’ further quality control. Below is an example of problematic prescription.

| **drug_name** | **quantity** | **chem_name** | **multiplier** |
| --- | --- | --- | --- |
| nefazodone starter pack | tablet(s) - 14 x 50 mg, 14 x 100 mg, 28 x 200 mg | nefazodone | 999 |
| nefazodone starter pack | 1 - tablets (14x50mg,14x100mg,28x200mg) | nefazodone | 999 |

On the above two prescriptions, it contains string patterns that cannot be resolved by `*multiplier_extract()*`, as there are multiple strings that matches to the multiplier pattern “x”. The “*multiplier*” in this case cannot be ascertained and need to be resolved manually. Furthermore, users can control whether to remove these problematic prescriptions using the `*qc_remove*` argument in the function.

A sample log file for `*multiplier_extract()*` is shown below, detailing message history if multiple strings matching multiplier patterns are found, and whether filters are applied for removing problematic prescriptions.

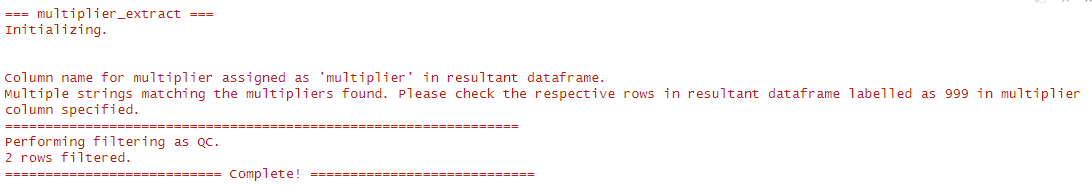

Details of `*multiplier_extract()*` and the respective functions of arguments are described on T-Rx website at: <https://chrislowh.github.io/T-Rx/>.

**Inferring quantities and duration of treatment from UKB-formatted strings**

The *`multi_num_infer()`* function in T-Rx is specifically designed to infer quantity information in UKB primary care prescription records. This leverages specific patterns of prescription records in UKB, where the integers typically represent one of product strengths, quantity or multipliers. This would extract quantities of prescriptions where dosage form strings were not specified.

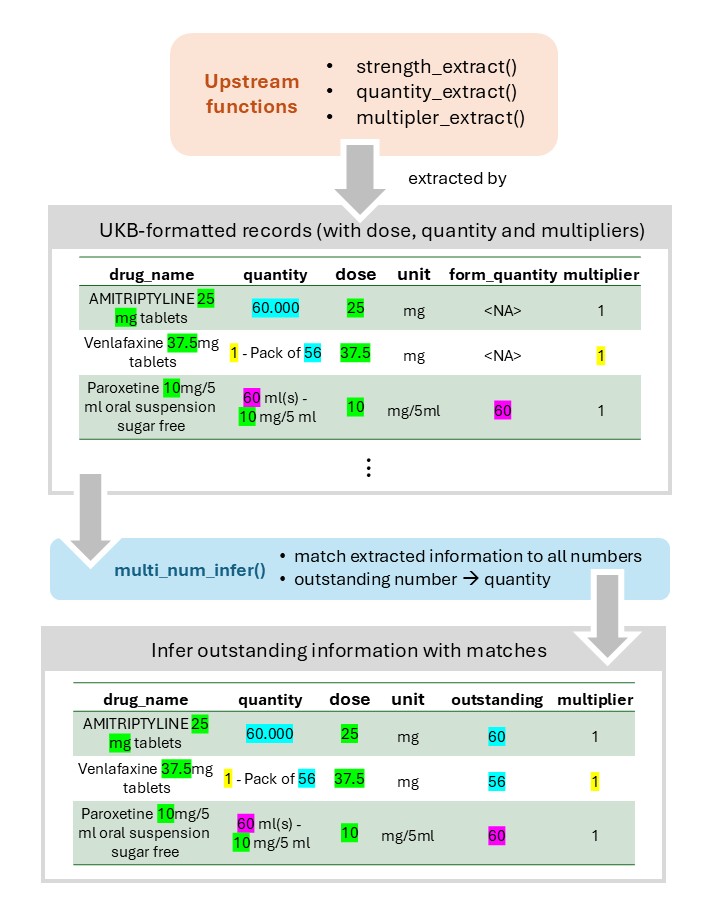

As an illustration, a sample prescription dataframe is shown below. The strengths and quantities of products were highlighted in yellow and blue respectively. Information on multipliers on products was highlighted in green.

| **drug_name** | **quantity** |
| --- | --- |
| AMITRIPTYLINE 25mg tablets | 60.000 |
| Venlafaxine 37.5mg tablets | 2 packs of 56 |
| Paroxetine 10mg/5ml oral suspension sugar free | 50 ml(s) – 10 mg/5 ml |

If strengths and multiplier information were extracted from upstream functions (with `*strength_extract()*` and *`multiplier_extract()`*, the remainder value, i.e.: quantity, can be inferred. An example output after running the above functions is shown below.

| **drug_name** | **quantity** | **strength** | **strength unit** | **multiplier** |
| --- | --- | --- | --- | --- |
| AMITRIPTYLINE 25mg tablets | 60.000 | 25 | mg | 1 |
| Venlafaxine 37.5mg tablets | 2 packs of 56 | 37.5 | mg | 2 |
| Paroxetine 10mg/5ml oral suspension sugar free | 50 ml(s) – 10 mg/5 ml | 10 | mg/5ml | 1 |

In the above prescriptions, the quantities are highlighted in blue, but `*quantity_extract()`* would not extract these numbers as quantity because of the absence of dosage form strings (e.g.: *tablets, capsules*). *`multi_num_infer()`* infers to extract these numbers as quantities by matching all integers shown in the *drug_name* and *quantity* columns to the integers that have already been extracted, i.e.: *strength* and *multiplier*. The outstanding number, given there is only one number outstanding, would be inferred as the quantity. The details and limitations of `*multi_num_infer()*` are described on T-Rx website at: <https://chrislowh.github.io/T-Rx/>.

##### **Handling duration-based strings in UKB primary care prescriptions**

In UKB primary care records, the quantities of prescriptions were sometimes described as prescription duration. An example is shown below, with duration-based strings highlighted in blue:

| **drug_name** | **quantity** |
| --- | --- |
| Cipramil 20mg tablets | 4/52 |
| Venlafaxine 37.5mg tablets | 1 – Pack of 56 |
| Fluoxetine 20mg capsules | 1 month – 20 mg |

These duration-based strings were commonly used in prescriptions, representing months, weeks and days respectively. Based on these strings, `*duration_handling()*` converts these strings into quantities of prescriptions.

| **Duration** | **Strings** |
| --- | --- |
| months | /12 |
| weeks | /52 |
| days | /365 |

### **Exposure Ascertainment Module**

#### **Merging prescriptions to exposure periods**

As described in the main text, `*rx_merge()*` function in T-Rx merges prescriptions to periods of exposure, with customisable gaps allowed between prescriptions. A graphical illustration is shown in **Supplementary figure S4.** This approach is preferred when the prescription durations are available in the prescription data, i.e.: the start dates of prescriptions, and one of the end dates or durations of prescriptions. The figure below shows possible differences in how exposure periods will be constructed by allowing different gaps between prescriptions, specified using the *gap* argument in `*rx_merge()*`.

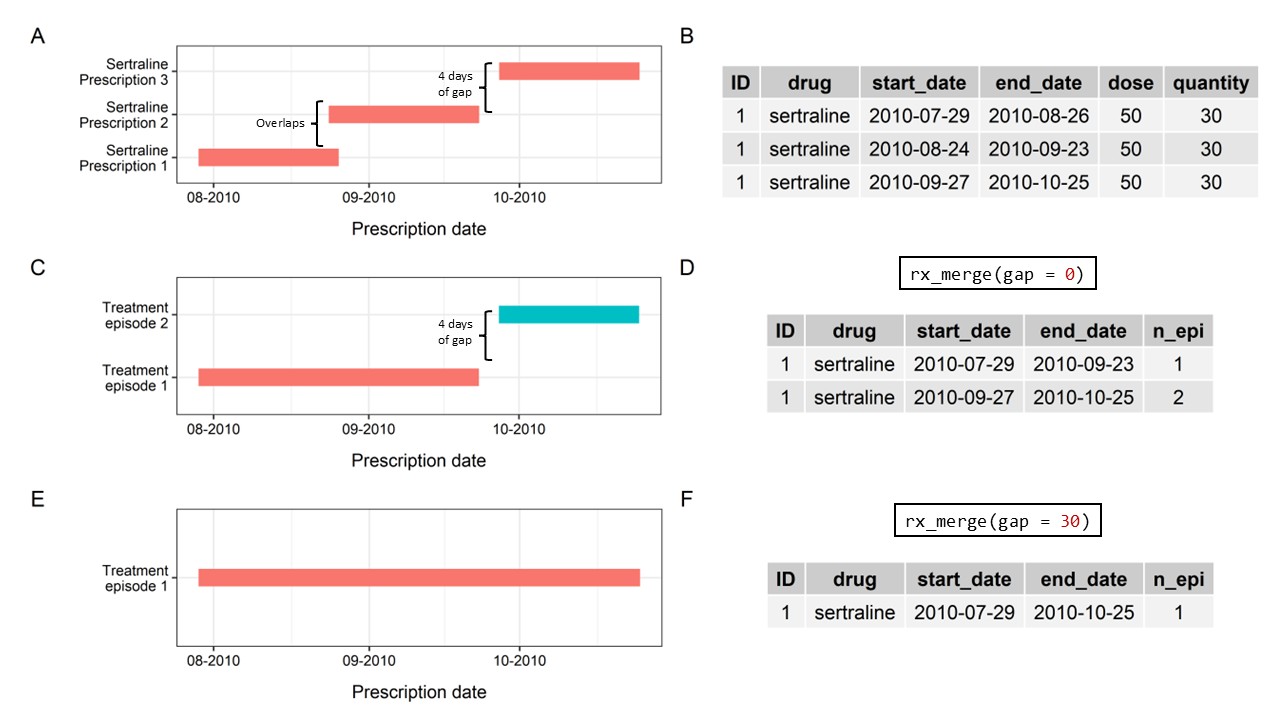

**Panel A/B** is a sample of three sertraline prescriptions, with the start and end dates of prescriptions available in the dataframe. If no gaps were allowed between prescriptions, two treatment episodes will be constructed (**Panel C/D**), due to the 4-day gap between sertraline prescriptions. Users can also allow for a gap (for real-life treatment complexities) – here we demonstrate a sample with 30-day gap, which would merge all three sertraline prescriptions into one treatment episode (**Panel E/F**).

##### **Inferring exposure periods from prescriptions without prescription durations**

In many EHR databases, the coverages of prescriptions were often not available (such as UKB primary care records). The `*rx_infer()*`function in T-Rx allows users to make inferences from “repeated prescriptions”.

The blocks of “repeated prescriptions” from consecutive prescriptions that were:

- of the same drug;
- of the same dosage (optional);
- of the same frequency or quantity (optional); and
- close enough in prescription dates (specified by *rx_window_days* argument).

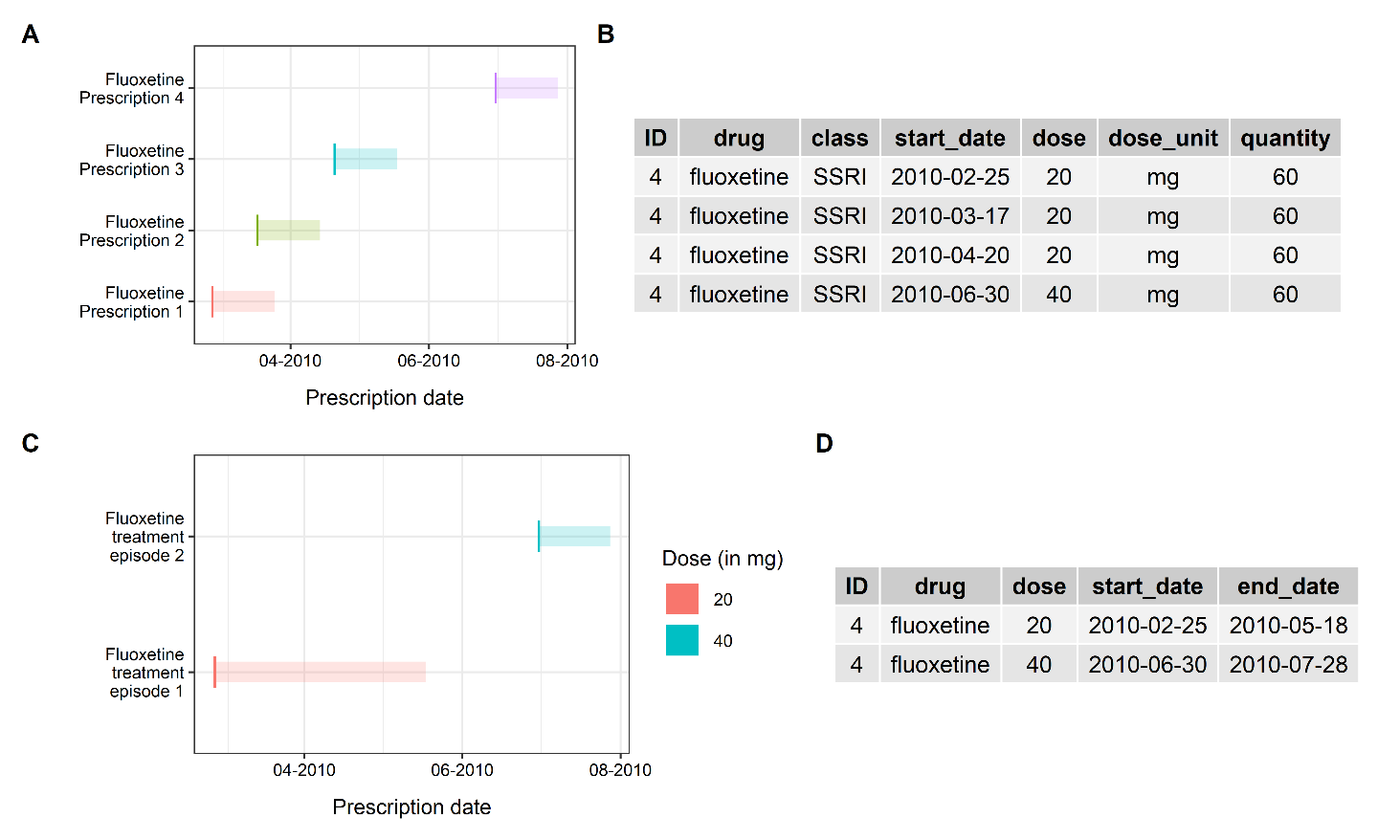

An example of “repeated prescription” block is shown above in the first three fluoxetine prescriptions, consisting of the same dosage (*20mg*) and quantity (*60*) (**Panel B**). The segments represent the expected exposure periods for each prescription assuming each prescription lasts for 28 days.

Using the above prescriptions as input, we can run `*rx_infer()*` as

```
fluoxetine_rx_infer = rx_infer(rx_df = fluoxetine_rx, id_col = "ID",
				  drug_col = "drug", dose_col = "dose",
				  date_col = "start_date",
                               rx_window_days = 98, assume_days = 28)
```

The *rx_window_days* argument controls how many days between the dates of two consecutive prescriptions are allowed to be treated as the same treatment episode, i.e.: how “far” the two prescriptions are apart. The “repeated prescription” blocks will be merged together into a longitudinal episode using `*rx_infer()*` (**Panel C/D**). The *assume_days* argument places assumptions on the length of the final or single prescription of the episode. Here, we used 28 days as an example for making such assumptions, yet the functions are flexible to different input days, depending on datasets and research areas of interest. As shown in **Panel C/D**, the final fluoxetine prescription is not treated as a “repeated prescription” because the dosage has been changed.

### **Phenotyping Module**

In the T-Rx phenotyping module, users can create proxy phenotypes with a one-line R command, by specifying the data frame of prescription records or episodes (with information on column names) and quality control parameters. In T-Rx, we provide two examples to create these phenotypes in antidepressant switching ^10^ and treatment-resistant depression events ^4^, described in published literature. The details in creating the phenotypes are summarized in main text.

#### **Antidepressant switching**

Switching is a proxy phenotype to capture medication non-response using prescription records, with phenotyping details described elsewhere ^10,11^. The phenotyping scripts have three steps to identify switching events, based on gaps between prescription dates of two different medications, imposing additional quality control criteria and identifying comparable controls ^10^.

#### **Treatment-resistant depression (TRD)**

TRD was defined and validated previously using at least two antidepressant switches within 14 weeks, with further quality control parameters ^4^. T-Rx concatenated the phenotyping algorithm for ease of use as `*TRD_Fabbri2021()*` (**Figure 1E**). This allows ready comparison with other TRD algorithms, given the heterogeneity of definitions in the literature ^12^.

**Supplementary Tables**

#### **Supplementary table S1. READ v2, BNF or dm+d codes codes used to extract antipsychotic prescriptions from UKB primary care records**

| **Code System** | **List of codes** |
| --- | --- |
| **READ v2** | d4t.., d4t1., d4t2., d4t3., d4t4., d4t5., d4t6., d4t7., d4tx., d4ty., d4tz., d4v.., d4v1., d4v2., d4v3., d4v4., d4v5., d4v6., d4v7., d4v8., d4v9., d4vr., d4vs., d4vt., d4vu., d4vv., d4vw., d4vx., d4vy., d4vz., d4x.., d4x1., d4x2., d4x3., d4x4., d42.., d421., d42z., dn3.., dn31., dn32., dn33., dn34., dn35., dn36., dn37., dn38., dn39., dn3a., dn3b., dn3c., dn3d., dn3D., dn3E., dn3J., dn3K., dn3v., dn3w., dn3x., dn3y., dn3z., d41.., d411., d412., d413., d414., d415., d41A., d41B., d41f., d41h., d41j., d41k., d41l., d41m., d41o., dh2.., o52.., o522., o52y., o52z., d4l.., d4l1., d4l2., d4l3., d4l4., d4l5., d4l6., d4l7., d4l8., d4l9., d4lA., d4lB., d4lC., d4lD., d4lE., d4lF., d4lG., d45.., d451., d45z., d51.., d511., d513., d516., d517., d518., d519., d51a., d51u., d51v., d51w., d51y., da1.., da11., da12., da1y., da1z., d46.., d46x., d46y., d46z., d52.., d521., d522., d524., d527., d528., d529., d52a., d52s., d52t., d52u., d52v., d52w., d52x., d531., d532., d47.., d471., d472., d473., d474., d475., d476., d47A., d47B., d47f., d47j., d47k., d47l., d47m., d47n., d47o., d47p., d47q., d47r., d47s., d47t., d47u., d47w., d47x., d47y., d55.., d551., d552., d553., d554., ds1.., d48.., d481., d483., d48y., d48z., d6..., d61.., d612., d613., d615., d616., d617., d618., d619., d61s., d61v., d61w., d61x., d61y., d61z., d62.., d622., d623., d624., d625., d62w., d62x., d62y., d62z., mb4.., mb41., mb43., d4k.., d4y.., d4y1., d4y2., d4y3., d4y4., d4y5., d4y6., d4r.., d4r1., d4r2., d4r3., d4r4., d4r5., d4r6., d4r7., d4r8., d4r9., d4rA., d4rB., d4rC., d4rD., d4rE., d4rF., d4rG., d4rH., d4rI., d4rJ., d4rK., d4rL., d4rM., d4rN., d4rO., d4rP., d4rt., d4ru., d4rv., d4rw., d4rx., d4ry., d4rz., d58.., d581., d582., d583., d58x., d58y., d58z., d4w.., d4w1., d4w2., d4w3., d4w5., d4w6., d4w7., d4w8., d4w9., d4wr., d4ws., d4wt., d4wu., d4wv., d4wx., d4wy., d4wz., d4a.., d4aw., d4ax., d4az., d4b.., d4b1., d4b2., d4b5., d4b6., d4bx., d4by., d916., o58z., d4c.., d4c2., d4cy., d56.., d561., d562., d563., d564., d4d.., dhe.., dhe1., dhe3., dhe4., dhea., dheb., dheB., dhec., dhed., dher., dhet., dheu., dhev., dhew., dhex., dhey., dhez., d4e.., d4e4., d4e5., d4ev., d4ew., d4ey., c8i.., c8i1., c8i2., c8i3., c8i4., c8i6., c8i8., c8i9., c8iu., c8iv., c8iw., c8ix., c8iy., c8iz., d19.., dhf.., dhg.., dhg1., dhg2., dhgz., djg5., o485., o59.., d4s.., d4s1., d4s2., d4s3., d4s4., d4s5., d4s6., d4s7., d4s8., d4s9., d4sa., d4sA., d4sb., d4sB., d4sc., d4sC., d4sd., d4sD., d4se., d4sE., d4sf., d4sF., d4sg., d4sG., d4sh., d4sH., d4si., d4sI., d4sj., d4sJ., d4sk., d4sK., d4sL., d4sM., d4sN., d4sO., d4sP., d4sQ., d4sR., d4ss., d4sS., d4st., d4sT., d4su., d4sU., d4sv., d4sV., d4sw., d4sW., d4sx., d4sX., d4sy., d4sY., d4sz., d4sZ., d4m.., d4m1., d4m2., d4p.., d4p1., d4p2., d4p3., d4p4., d4p5., d4p6., d4p7., d4p8., d4p9., d4pA., d4pB., d4pC., d4pD., d4pE., d4pF., d4pG., d4pH., d4pJ., d4pK., d4pL., d4pM., d4pN., d4pO., d4pP., d4pQ., d4pR., d4pS., d4pw., d4px., d4py., d4pz., d4q.., d4q1., d4q2., d4q3., d4q4., d4q5., d4q6., d4q7., d4q8., d4f.., d4f1., d4f5., d4f6., d4fw., d4fx., d4fy., d4fz., d4g.., d4g6., d4g7., d4gp., d4gq., d4gr., d4gs., d4gu., d4gv., d4gw., d4gx., d4gy., d4h.., d4h7., d4h9., d4hA., d4hr., d4hs., d4ht., d4hx., d4hy., d4hz., dhi.., dnb.., dnb1., dnb2., dnb3., dnb4., dnb5., dnb6., dnb7., dnb8., dnb9., dnbA., dnbB., dnbc., dnbC., dnbd., dnbD., dnbe., dnbE., dnbF., dnbG., dnbH., dnbI., dnbJ., dnbK., dnbL., dnbM., dnbn., dnbN., dnbo., dnbO., dnbp., dnbP., dnbq., dnbQ., dnbr., dnbR., dnbs., dnbS., dnbt., dnbT., dnbu., dnbU., dnbv., dnbw., dnbx., dnby., dnbz., dnh.., dnh1., dnh2., dnh3., dnh4., dnh5., dnh6., dnh7., dnh8., dnhy., dnhz., d4u.., d4j.., d4j1., d4j2., d4j3., d4jx., d4jy., d4jz., d4n.., d4n1., d4n2., d4n3., d4n4., d57.., d571., d573., d574., d575., d576., d577., d578., d57y., d57z. |
| **BNF** | 04.02.01.00.00, 04.02.02.00.00, 04.02.03.01.00, 04.02.03.02.00, 04.02.03.03.00, 04.02.03.04.00, 040201, 04020100, 04020101, 040201010AAAAAA, 04020102, 040201030AAAAAA, 040201030AAABAB, 040201030AAACAC, 040201030AAADAD, 040201030AAAEAE, 040201030AAAFAF, 040201030BBAAAA, 040201030BBACAC, 040201060AAAAAA, 040201060AAABAB, 040201060AAACAC, 040201060AAADAD, 0402010A0AAAAAA, 0402010A0AAABAB, 0402010ABAAABAB, 0402010ABAAACAC, 0402010ABAAADAD, 0402010D0AAABAB, 0402010D0AAAHAH, 0402010D0AAAIAI, 0402010D0AAAJAJ, 0402010D0AAAKAK, 0402010I0AAABAB, 0402010J0AAAAAA, 0402010J0AAAFAF, 0402010J0AAAJAJ, 0402010J0AAAKAK, 0402010J0AAALAL, 0402010L0BBAAAA, 0402010S0AAAEAE, 0402010T0BBAAAA, 0402010U0AAAEAE, 0402010U0AAAHAH, 0402010U0AAAQAQ, 0402010U0BBAAAH, 0402010W0AAAEAE, 0402010W0AAAFAF, 0402010W0AAAGAG, 0402010W0AAAHAH, 0402010W0BBAAAE, 0402010W0BBACAG, 0402010X0AAABAB, 0402010X0AAAFAF, 0402010X0AAAHAH, 0402010X0AAAIAI, 0402010X0AAAKAK, 0402010X0BBABAI, 0402010X0BBAEAC, 0402010X0BBAFAF, 040202, 04020200, 0402020G0BBABAC, 040203, 04020300, 0402030K0AAACAC, 0402030K0AAAFAF, 0402030K0AAAIAI, 0402030K0BBAAAC, 0402030K0BBABAF, 0402030K0BDAAAG, 0402030K0BFAAAF, 0402030K0BFABAI, 0402030P0AAAIAI, 0402030P0BCAAAI |
| **dm+d** | 406788008, 299275001000027136, 662611000001103, 261911000001101, 125111000001109, 9536311000001104, 64895001000027104, 133385001000027104, 133395001000027104, 95785001000027104, 516511000001106, 109725001000027104, 153375001000027104, 29535001000027108, 83015001000027104, 593211000001102, 184055001000027104 |

**Abbreviations**

BNF = British National Formulary; dm+d = Dictionary of Medicines and Devices; UKB = UK Biobank.

#### **Supplementary table S2. List of diabetes codes used for CPRD validation sample**

| **MedCodeId** | **Original Read Code** | **Cleansed Read Code** | **Term** | **SnomedCTConceptId** | **SnomedCTDescriptionId** |
| --- | --- | --- | --- | --- | --- |
| 356085010 | C10D-1 | C10D.11 | Maturity onset diabetes in youth type 2 | 237604008 | 356085010 |
| 483882011 | C10C | C10C.00 | Maturity-onset diabetes of the young | 609561005 | 2967884018 |
| 483886014 | C10C-1 | C10C.11 | Maturity onset diabetes in youth | 609561005 | 2967884018 |
| 2476117016 | C10C-2 | C10C.12 | Maturity onset diabetes in youth type 1 | 609562003 | 2967867012 |
| 840971000006112 | C10D | C10D.00 | Diabetes mellitus autosomal dominant type II | 237604008 | 356082013 |
| 1968641000006110 | C10Q | C10Q.00 | Maturity-onset diabetes of the young, type 5 | 609572000 | 2967853015 |
| 292495016 | C104y | C104y00 | Other specified diabetes mellitus with renal complications | 127013003 | 301016 |
| 293756010 | Cyu20 | Cyu2000 | [X]Other specified diabetes mellitus | 73211009 | 121589010 |
| 13751000006117 | C108y | C108y00 | Other specified diabetes mellitus with multiple comps | 385041000000108 | 760111000000115 |
| 13781000006113 | C10yy | C10yy00 | Other specified diabetes mellitus with other spec comps | 73211009 | 121589010 |
| 1233307015 | PKyP-1 | PKyP.11 | Wolfram syndrome | 70694009 | 1233307015 |
| 189711000000119 | C10M0 | C10M000 | Lipoatrophic diabetes mellitus without complication | 112991000000101 | 189711000000119 |
| 494831000000119 | C10N1 | C10N100 | Diabetes mellitus associated with cystic fibrosis | 426705001 | 2674608014 |
| 622221000000118 | C10FS | C10FS00 | Maternally inherited diabetes mellitus | 335621000000101 | 622221000000118 |
| 967701000006116 | C10M | C10M.00 | Lipoatrophic diabetes mellitus | 127012008 | 300015 |
| 1848531000006110 | PKyP | PKyP.00 | Diabetes insipidus, diabetes mellitus, optic atrophy and deafness | 70694009 | 3028935019 |
| 15518018 | C10N | C10N.00 | Secondary diabetes mellitus | 8801005 | 15518018 |
| 1230929011 | C10G | C10G.00 | Secondary pancreatic diabetes mellitus | 51002006 | 1230929011 |
| 2160090014 | C10H | C10H.00 | Diabetes mellitus induced by non-steroid drugs | 408540003 | 2160090014 |
| 189721000000113 | C10N0 | C10N000 | Secondary diabetes mellitus without complication | 8801005 | 15518018 |
| 198461000000116 | C10G0 | C10G000 | Diabetes mellitus associated with pancreatic disease | 51002006 | 84990019 |
| 967631000006115 | C10H0 | C10H000 | Diabetes mellitus induced by non-steroid drugs without complication | 413183008 | 2474729016 |
| 197984010 | C10E | C10E.00 | Type 1 diabetes mellitus | 46635009 | 197984010 |
| 292538019 | C10E5-1 | C10E511 | Type I diabetes mellitus with ulcer | 190368000 | 292538019 |
| 292540012 | C10E5 | C10E500 | Type 1 diabetes mellitus with ulcer | 190368000 | 292540012 |
| 292541011 | C10E6 | C10E600 | Type 1 diabetes mellitus with gangrene | 420825003 | 2618207019 |
| 292543014 | C10E6-1 | C10E611 | Type I diabetes mellitus with gangrene | 420825003 | 2618207019 |
| 292548017 | C10E8 | C10E800 | Type 1 diabetes mellitus - poor control | 444073006 | 2872487013 |
| 292550013 | C10E8-1 | C10E811 | Type I diabetes mellitus - poor control | 444073006 | 2872487013 |
| 292551012 | C10E9-1 | C10E911 | Type I diabetes mellitus maturity onset | 190372001 | 292551012 |
| 292553010 | C10E9 | C10E900 | Type 1 diabetes mellitus maturity onset | 190372001 | 292553010 |
| 429970018 | C10E4-1 | C10E411 | Unstable type I diabetes mellitus | 290002008 | 429970018 |
| 429971019 | C10E4 | C10E400 | Unstable type 1 diabetes mellitus | 290002008 | 429971019 |
| 457325013 | C10EA-1 | C10EA11 | Type I diabetes mellitus without complication | 313435000 | 457325013 |
| 457326014 | C10EA | C10EA00 | Type 1 diabetes mellitus without complication | 313435000 | 457326014 |
| 459161015 | C10EE | C10EE00 | Type 1 diabetes mellitus with hypoglycaemic coma | 314771006 | 459161015 |
| 459163017 | C10EE-1 | C10EE11 | Type I diabetes mellitus with hypoglycaemic coma | 314771006 | 459163017 |
| 459292011 | C10EH | C10EH00 | Type 1 diabetes mellitus with arthropathy | 314893005 | 459292011 |
| 459294012 | C10EH-1 | C10EH11 | Type I diabetes mellitus with arthropathy | 314893005 | 459294012 |
| 459296014 | C10EJ | C10EJ00 | Type 1 diabetes mellitus with neuropathic arthropathy | 71771000119100 | 3010513018 |
| 494564012 | C10E-1 | C10E.11 | Type I diabetes mellitus | 46635009 | 494564012 |
| 1780311019 | C10EL | C10EL00 | Type 1 diabetes mellitus with persistent microalbuminuria | 401110002 | 1780311019 |
| 299601000000114 | C10EQ | C10EQ00 | Gastroparesis with type 1 diabetes mellitus | 713702000 | 3698406016 |
| 913451000006117 | C10E0 | C10E000 | Type 1 diabetes mellitus with renal complications | 421893009 | 2618211013 |
| 913461000006115 | C10E0-1 | C10E011 | Type I diabetes mellitus with renal complications | 421893009 | 2623054013 |
| 913481000006113 | C10E1 | C10E100 | Type 1 diabetes mellitus with ophthalmic complications | 739681000 | 3537386015 |
| 913491000006111 | C10E1-1 | C10E111 | Type I diabetes mellitus with ophthalmic complications | 739681000 | 3698438014 |
| 913511000006117 | C10E2 | C10E200 | Type 1 diabetes mellitus with neurological complications | 421468001 | 2618197018 |
| 913521000006113 | C10E2-1 | C10E211 | Neurological disorder with type 1 diabetes mellitus | 421468001 | 3695401013 |
| 913541000006118 | C10E3 | C10E300 | Type 1 diabetes mellitus with multiple complications | 422228004 | 2618232017 |
| 913551000006116 | C10E3-1 | C10E311 | Type I diabetes mellitus with multiple complications | 422228004 | 2967771016 |
| 913661000006118 | C10E7 | C10E700 | Retinopathy with type 1 diabetes mellitus | 420789003 | 3699407019 |
| 913671000006113 | C10E7-1 | C10E711 | Type I diabetes mellitus with retinopathy | 420789003 | 3699407019 |
| 913781000006117 | C10EB | C10EB00 | Type 1 diabetes mellitus with mononeuropathy | 420918009 | 2618199015 |
| 913791000006119 | C10EB-1 | C10EB11 | Type I diabetes mellitus with mononeuropathy | 420918009 | 3697664014 |
| 913811000006115 | C10EC | C10EC00 | Type 1 diabetes mellitus with polyneuropathy | 713705003 | 3297342019 |
| 913821000006111 | C10EC-1 | C10EC11 | Type I diabetes mellitus with polyneuropathy | 713705003 | 3297390014 |
| 913841000006116 | C10ED | C10ED00 | Renal disorder associated with type 1 diabetes mellitus | 421893009 | 2623054013 |
| 913851000006119 | C10ED-1 | C10ED11 | Type I diabetes mellitus with nephropathy | 421893009 | 2623054013 |
| 913901000006112 | C10EF | C10EF00 | Type 1 diabetes mellitus with diabetic cataract | 421920002 | 2618234016 |
| 913911000006110 | C10EF-1 | C10EF11 | Type I diabetes mellitus with diabetic cataract | 421920002 | 3688521010 |
| 913931000006116 | C10EG | C10EG00 | Type 1 diabetes mellitus with peripheral angiopathy | 31211000119101 | 3315042015 |
| 913941000006114 | C10EG-1 | C10EG11 | Peripheral angiopathy due to type 1 diabetes mellitus | 31211000119101 | 3315042015 |
| 928461000006119 | C10EK | C10EK00 | Persistent proteinuria associated with type 1 diabetes mellitus | 420514000 | 2967914011 |
| 928471000006114 | C10EK-1 | C10EK11 | Type I diabetes mellitus with persistent proteinuria | 420514000 | 2967914011 |
| 928491000006110 | C10EL-1 | C10EL11 | Type I diabetes mellitus with persistent microalbuminuria | 401110002 | 1780311019 |
| 928501000006119 | C10EM | C10EM00 | Ketoacidosis in type 1 diabetes mellitus | 420270002 | 2967817017 |
| 928511000006116 | C10EM-1 | C10EM11 | Type I diabetes mellitus with ketoacidosis | 420270002 | 2967817017 |
| 928521000006112 | C10EN | C10EN00 | Ketoacidotic coma in type 1 diabetes mellitus | 421075007 | 2967758019 |
| 928531000006110 | C10EN-1 | C10EN11 | Type I diabetes mellitus with ketoacidotic coma | 421075007 | 2967758019 |
| 938301000006114 | C10EP | C10EP00 | Exudative maculopathy with type 1 diabetes mellitus | 420486006 | 3698521013 |
| 938311000006112 | C10EP-1 | C10EP11 | Type I diabetes mellitus with exudative maculopathy | 420486006 | 2618236019 |
| 1667941000000110 | C10EQ-1 | C10EQ11 | Type I diabetes mellitus with gastroparesis | 713702000 | 3698406016 |
| 2288011000000110 | C10P0 | C10P000 | Type I diabetes mellitus in remission | 703137001 | 3007238018 |
| 2288041000000110 | C10P0-1 | C10P011 | Type 1 diabetes mellitus in remission | 703137001 | 3007274010 |
| 72711000006117 | C1084-2 | C108412 | Unstable type 1 diabetes mellitus | 290002008 | 429971019 |
| 72721000006113 | C1084-1 | C108411 | Unstable type I diabetes mellitus | 290002008 | 429970018 |
| 84281000006115 | C108-2 | C108.12 | Type 1 diabetes mellitus | 46635009 | 197984010 |
| 84291000006117 | C1088-2 | C108812 | Type 1 diabetes mellitus - poor control | 444073006 | 2872487013 |
| 84301000006116 | C1089-2 | C108912 | Type 1 diabetes mellitus maturity onset | 190372001 | 292553010 |
| 84311000006118 | C108H-2 | C108H12 | Type 1 diabetes mellitus with arthropathy | 314893005 | 459292011 |
| 84321000006114 | C108F-2 | C108F12 | Type 1 diabetes mellitus with diabetic cataract | 421920002 | 2618234016 |
| 84331000006112 | C1086-2 | C108612 | Type 1 diabetes mellitus with gangrene | 420825003 | 2618207019 |
| 84341000006119 | C108E-2 | C108E12 | Type 1 diabetes mellitus with hypoglycaemic coma | 314771006 | 459161015 |
| 84361000006115 | C1083-2 | C108312 | Type 1 diabetes mellitus with multiple complications | 422228004 | 2618232017 |
| 84371000006110 | C108D-2 | C108D12 | Type 1 diabetes mellitus with nephropathy | 421893009 | 2623054013 |
| 84381000006113 | C1082-2 | C108212 | Type 1 diabetes mellitus with neurological complications | 421468001 | 2618197018 |
| 84391000006111 | C108J-2 | C108J12 | Type 1 diabetes mellitus with neuropathic arthropathy | 71771000119100 | 3010513018 |
| 84401000006113 | C1081-2 | C108112 | Type 1 diabetes mellitus with ophthalmic complications | 739681000 | 3537386015 |
| 84421000006115 | C108C-2 | C108C12 | Type 1 diabetes mellitus with polyneuropathy | 713705003 | 3297342019 |
| 84431000006117 | C1080-2 | C108012 | Type 1 diabetes mellitus with renal complications | 421893009 | 2618211013 |
| 84441000006110 | C1087-2 | C108712 | Type 1 diabetes mellitus with retinopathy | 420789003 | 3699407019 |
| 84451000006112 | C1085-2 | C108512 | Type 1 diabetes mellitus with ulcer | 190368000 | 292540012 |
| 84461000006114 | C108A-2 | C108A12 | Type 1 diabetes mellitus without complication | 313435000 | 457326014 |
| 84651000006114 | C108-3 | C108.13 | Type I diabetes mellitus | 46635009 | 494564012 |
| 84661000006111 | C1088-1 | C108811 | Type I diabetes mellitus poorly controlled | 444073006 | 2872487013 |
| 84671000006116 | C1089-1 | C108911 | Type I diabetes mellitus maturity onset | 190372001 | 292551012 |
| 84681000006118 | C108H-1 | C108H11 | Type I diabetes mellitus with arthropathy | 314893005 | 459294012 |
| 84691000006115 | C108F-1 | C108F11 | Cataract due to diabetes mellitus type 1 | 421920002 | 3688521010 |
| 84701000006115 | C1086-1 | C108611 | Type I diabetes mellitus with gangrene | 420825003 | 2618207019 |
| 84711000006117 | C108E-1 | C108E11 | Type I diabetes mellitus with hypoglycaemic coma | 314771006 | 459163017 |
| 84721000006113 | C108B-1 | C108B11 | Mononeuropathy with type 1 diabetes mellitus | 420918009 | 3697664014 |
| 84731000006111 | C1083-1 | C108311 | Multiple complications of type 1 diabetes mellitus | 422228004 | 2967771016 |
| 84741000006118 | C108D-1 | C108D11 | Type I diabetes mellitus with nephropathy | 421893009 | 2623054013 |
| 84751000006116 | C1082-1 | C108211 | Type I diabetes mellitus with neurological complications | 421468001 | 3695401013 |
| 84761000006119 | C108J-1 | C108J11 | Type I diabetes mellitus with neuropathic arthropathy | 71771000119100 | 3010513018 |
| 84771000006114 | C1081-1 | C108111 | Disorder of eye with type 1 diabetes mellitus | 739681000 | 3698438014 |
| 84791000006110 | C108C-1 | C108C11 | Polyneuropathy due to diabetes mellitus type I | 713705003 | 3297390014 |
| 84801000006111 | C1080-1 | C108011 | Type I diabetes mellitus with renal complications | 421893009 | 2623054013 |
| 84811000006114 | C1087-1 | C108711 | Type I diabetes mellitus with retinopathy | 420789003 | 3699407019 |
| 84821000006118 | C1085-1 | C108511 | Type I diabetes mellitus with ulcer | 190368000 | 292538019 |
| 84831000006115 | C108A-1 | C108A11 | Type I diabetes mellitus without complication | 313435000 | 457325013 |
| 616611000006111 | C10z0 | C10z000 | Disorder due to type 1 diabetes mellitus | 420868002 | 3013528019 |
| 616621000006115 | C1000 | C100000 | Type 1 diabetes mellitus without complication | 313435000 | 457326014 |
| 197761014 | C10F | C10F.00 | Type 2 diabetes mellitus | 44054006 | 197761014 |
| 292576013 | C10F3-1 | C10F311 | Type II diabetes mellitus with multiple complications | 190388001 | 292576013 |
| 292577016 | C10F3 | C10F300 | Type 2 diabetes mellitus with multiple complications | 190388001 | 292577016 |
| 292579018 | C10F4 | C10F400 | Type 2 diabetes mellitus with ulcer | 190389009 | 292579018 |
| 292581016 | C10F4-1 | C10F411 | Type II diabetes mellitus with ulcer | 190389009 | 292581016 |
| 292582011 | C10F5-1 | C10F511 | Type II diabetes mellitus with gangrene | 421631007 | 2618206011 |
| 292583018 | C10F5 | C10F500 | Type 2 diabetes mellitus with gangrene | 421631007 | 2618206011 |
| 292589019 | C10F7-1 | C10F711 | Type II diabetes mellitus - poor control | 443694000 | 2921019012 |
| 292590011 | C10F7 | C10F700 | Type II diabetes mellitus poorly controlled | 443694000 | 2921019012 |
| 457329019 | C10F9 | C10F900 | Type 2 diabetes mellitus without complication | 313436004 | 457329019 |
| 457330012 | C10F9-1 | C10F911 | Type II diabetes mellitus without complication | 313436004 | 457330012 |
| 459167016 | C10FD | C10FD00 | Type 2 diabetes mellitus with hypoglycaemic coma | 719216001 | 3316336015 |
| 459169018 | C10FD-1 | C10FD11 | Hypoglycaemic coma co-occurrent and due to diabetes mellitus type II | 719216001 | 3316336015 |
| 459306016 | C10FF-1 | C10FF11 | Type II diabetes mellitus with peripheral angiopathy | 314902007 | 459306016 |
| 459308015 | C10FF | C10FF00 | Type 2 diabetes mellitus with peripheral angiopathy | 314902007 | 459308015 |
| 459310018 | C10FG | C10FG00 | Type 2 diabetes mellitus with arthropathy | 314903002 | 459310018 |
| 459311019 | C10FG-1 | C10FG11 | Type II diabetes mellitus with arthropathy | 314903002 | 459311019 |
| 459312014 | C10FH-1 | C10FH11 | Type II diabetes mellitus with neuropathic arthropathy | 314904008 | 459312014 |
| 459313016 | C10FH | C10FH00 | Type 2 diabetes mellitus with neuropathic arthropathy | 314904008 | 459313016 |
| 493774016 | C10F-1 | C10F.11 | Type II diabetes mellitus | 44054006 | 493774016 |
| 1223147012 | C10FJ-1 | C10FJ11 | Insulin treated Type II diabetes mellitus | 237599002 | 1223147012 |
| 1488898011 | C10FK | C10FK00 | Hyperosmolar non-ketotic state in type 2 diabetes mellitus | 395204000 | 1488898011 |
| 299621000000117 | C10FR | C10FR00 | Gastroparesis with type 2 diabetes mellitus | 713703005 | 3698412014 |
| 840951000006119 | C109J | C109J00 | Insulin treated Type 2 diabetes mellitus | 237599002 | 2967820013 |
| 841351000006110 | C109J-2 | C109J12 | Insulin treated Type II diabetes mellitus | 237599002 | 1223147012 |
| 850691000006118 | C109K | C109K00 | Hyperosmolar non-ketotic state in type 2 diabetes mellitus | 395204000 | 1488898011 |
| 914031000006118 | C10F0 | C10F000 | Renal disorder associated with type II diabetes mellitus | 420279001 | 2618208012 |
| 914041000006111 | C10F0-1 | C10F011 | Type II diabetes mellitus with renal complications | 420279001 | 3013392012 |
| 914051000006113 | C10F1 | C10F100 | Disorder of eye with type 2 diabetes mellitus | 422099009 | 3698440016 |
| 914061000006110 | C10F1-1 | C10F111 | Type II diabetes mellitus with ophthalmic complications | 422099009 | 3698440016 |
| 914071000006115 | C10F2 | C10F200 | Neurologic disorder associated with type 2 diabetes mellitus | 421326000 | 3695406015 |
| 914081000006117 | C10F2-1 | C10F211 | Type II diabetes mellitus with neurological complications | 421326000 | 3695406015 |
| 914151000006112 | C10F6 | C10F600 | Retinopathy with type 2 diabetes mellitus | 422034002 | 3699410014 |
| 914161000006114 | C10F6-1 | C10F611 | Type II diabetes mellitus with retinopathy | 422034002 | 3699410014 |
| 914221000006113 | C10FA | C10FA00 | Type 2 diabetes mellitus with mononeuropathy | 420436000 | 2618201018 |
| 914231000006111 | C10FA-1 | C10FA11 | Type II diabetes mellitus with mononeuropathy | 420436000 | 3697667019 |
| 914241000006118 | C10FB | C10FB00 | Polyneuropathy due to type 2 diabetes mellitus | 713706002 | 3297353013 |
| 914251000006116 | C10FB-1 | C10FB11 | Type II diabetes mellitus with polyneuropathy | 713706002 | 3297353013 |
| 914261000006119 | C10FC | C10FC00 | Diabetes type 2 with nephropathy | 420279001 | 3035432019 |
| 914271000006114 | C10FC-1 | C10FC11 | Type II diabetes mellitus with nephropathy | 420279001 | 3013392012 |
| 914301000006111 | C10FE | C10FE00 | Type 2 diabetes mellitus with diabetic cataract | 420756003 | 3688522015 |
| 914311000006114 | C10FE-1 | C10FE11 | Type II diabetes mellitus with diabetic cataract | 420756003 | 3688522015 |
| 914391000006116 | C10FJ | C10FJ00 | Insulin treated Type 2 diabetes mellitus | 237599002 | 2967820013 |
| 928541000006117 | C10FL | C10FL00 | Persistent proteinuria associated with type 2 diabetes mellitus | 421986006 | 2967751013 |
| 928551000006115 | C10FL-1 | C10FL11 | Type II diabetes mellitus with persistent proteinuria | 421986006 | 2967751013 |
| 928561000006118 | C10FM | C10FM00 | Persistent microalbuminuria associated with type 2 diabetes mellitus | 420715001 | 2967769016 |
| 928571000006113 | C10FM-1 | C10FM11 | Type II diabetes mellitus with persistent microalbuminuria | 420715001 | 2967769016 |
| 928581000006111 | C10FN | C10FN00 | Ketoacidosis in type 2 diabetes mellitus | 421750000 | 2967754017 |
| 928591000006114 | C10FN-1 | C10FN11 | Type II diabetes mellitus with ketoacidosis | 421750000 | 2967754017 |
| 928601000006118 | C10FP | C10FP00 | Ketoacidotic coma in type 2 diabetes mellitus | 421847006 | 2618218019 |
| 928611000006115 | C10FP-1 | C10FP11 | Ketoacidotic coma in type II diabetes mellitus | 421847006 | 2618218019 |
| 938321000006116 | C10FQ | C10FQ00 | Exudative maculopathy associated with type 2 diabetes mellitus | 421779007 | 2618237011 |
| 938331000006118 | C10FQ-1 | C10FQ11 | Exudative maculopathy with type 2 diabetes mellitus | 421779007 | 3698529010 |
| 1667891000000110 | C10FK-1 | C10FK11 | Hyperosmolar non-ketotic state in type II diabetes mellitus | 395204000 | 1667891000000110 |
| 1667921000000110 | C10FR-1 | C10FR11 | Type II diabetes mellitus with gastroparesis | 713703005 | 3698412014 |
| 2288061000000110 | C10P1 | C10P100 | Type II diabetes mellitus in remission | 703138006 | 3007241010 |
| 2288071000000110 | C10P1-1 | C10P111 | Type 2 diabetes mellitus in remission | 703138006 | 3007259013 |
| 84471000006119 | C109-2 | C109.12 | Type 2 diabetes mellitus | 44054006 | 197761014 |
| 84481000006116 | C1097-2 | C109712 | Type 2 diabetes mellitus - poor control | 443694000 | 2921019012 |
| 84491000006118 | C109G-2 | C109G12 | Type 2 diabetes mellitus with arthropathy | 314903002 | 459310018 |
| 84501000006114 | C109E-2 | C109E12 | Type 2 diabetes mellitus with diabetic cataract | 420756003 | 3688522015 |
| 84511000006112 | C1095-2 | C109512 | Type 2 diabetes mellitus with gangrene | 421631007 | 2618206011 |
| 84521000006116 | C109D-2 | C109D12 | Type 2 diabetes mellitus with hypoglycaemic coma | 719216001 | 3316336015 |
| 84531000006118 | C109A-2 | C109A12 | Type 2 diabetes mellitus with mononeuropathy | 420436000 | 2618201018 |
| 84541000006111 | C1093-2 | C109312 | Type 2 diabetes mellitus with multiple complications | 190388001 | 292577016 |
| 84551000006113 | C109C-2 | C109C12 | Type 2 diabetes mellitus with nephropathy | 420279001 | 3035432019 |
| 84561000006110 | C1092-2 | C109212 | Type 2 diabetes mellitus with neurological complications | 421326000 | 3695406015 |
| 84571000006115 | C109H-2 | C109H12 | Type 2 diabetes mellitus with neuropathic arthropathy | 314904008 | 459313016 |
| 84581000006117 | C1091-2 | C109112 | Type 2 diabetes mellitus with ophthalmic complications | 422099009 | 3698440016 |
| 84591000006119 | C109F-2 | C109F12 | Type 2 diabetes mellitus with peripheral angiopathy | 314902007 | 459308015 |
| 84601000006110 | C109B-2 | C109B12 | Type 2 diabetes mellitus with polyneuropathy | 713706002 | 3297353013 |
| 84611000006113 | C1090-2 | C109012 | Type 2 diabetes mellitus with renal complications | 420279001 | 2618208012 |
| 84621000006117 | C1096-2 | C109612 | Type 2 diabetes mellitus with retinopathy | 422034002 | 3699410014 |
| 84631000006119 | C1094-2 | C109412 | Type 2 diabetes mellitus with ulcer | 190389009 | 292579018 |
| 84641000006112 | C1099-2 | C109912 | Type 2 diabetes mellitus without complication | 313436004 | 457329019 |
| 84841000006113 | C109-3 | C109.13 | Type II diabetes mellitus | 44054006 | 493774016 |
| 84851000006110 | C1097-1 | C109711 | Type II diabetes mellitus - poor control | 443694000 | 2921019012 |
| 84861000006112 | C109G-1 | C109G11 | Type II diabetes mellitus with arthropathy | 314903002 | 459311019 |
| 84871000006117 | C109E-1 | C109E11 | Cataract due to diabetes mellitus type 2 | 420756003 | 3688522015 |
| 84881000006119 | C1095-1 | C109511 | Type II diabetes mellitus with gangrene | 421631007 | 2618206011 |
| 84891000006116 | C109D-1 | C109D11 | Type II diabetes mellitus with hypoglycaemic coma | 719216001 | 3316336015 |
| 84901000006117 | C109A-1 | C109A11 | Mononeuropathy with type 2 diabetes mellitus | 420436000 | 3697667019 |
| 84911000006119 | C1093-1 | C109311 | Type II diabetes mellitus with multiple complications | 190388001 | 292576013 |
| 84921000006110 | C109C-1 | C109C11 | Renal disorder due to type 2 diabetes mellitus | 420279001 | 3013392012 |
| 84931000006113 | C1092-1 | C109211 | Neurological disorder with diabetes type 2 | 421326000 | 3695406015 |
| 84941000006115 | C109H-1 | C109H11 | Type II diabetes mellitus with neuropathic arthropathy | 314904008 | 459312014 |
| 84951000006118 | C1091-1 | C109111 | Disorder of eye with type 2 diabetes mellitus | 422099009 | 3698440016 |
| 84961000006116 | C109F-1 | C109F11 | Type II diabetes mellitus with peripheral angiopathy | 314902007 | 459306016 |
| 84971000006111 | C109B-1 | C109B11 | Type II diabetes mellitus with polyneuropathy | 713706002 | 3297353013 |
| 84981000006114 | C1090-1 | C109011 | Type II diabetes mellitus with renal complications | 420279001 | 3013392012 |
| 84991000006112 | C1096-1 | C109611 | Type II diabetes mellitus with retinopathy | 422034002 | 3699410014 |
| 85001000006117 | C1094-1 | C109411 | Type II diabetes mellitus with ulcer | 190389009 | 292581016 |
| 85011000006119 | C1099-1 | C109911 | Type II diabetes mellitus without complication | 313436004 | 457330012 |
| 281171000006119 | C1097 | C109700 | Type II diabetes mellitus uncontrolled | 443694000 | 2842387018 |
| 281211000006117 | C1095 | C109500 | Gangrene associated with type 2 diabetes mellitus | 421631007 | 2967779019 |
| 616501000006112 | C10z1 | C10z100 | Disorder due to type 2 diabetes mellitus | 422014003 | 3013072014 |
| 77727018 | C10E-2 | C10E.12 | Insulin dependent diabetes mellitus | 73211009 | 121589010 |
| 121589010 | C10 | C10..00 | Diabetes mellitus | 73211009 | 121589010 |
| 429972014 | C10E4-2 | C10E412 | Unstable insulin dependent diabetes mellitus | 11530004 | 19931010 |
| 457327017 | C10EA-2 | C10EA12 | Insulin-dependent diabetes without complication | 111552007 | 178796018 |
| 459162010 | C10EE-2 | C10EE12 | Insulin dependent diabetes mellitus with hypoglycaemic coma | 237632004 | 356141011 |
| 459293018 | C10EH-2 | C10EH12 | Insulin dependent diabetes mellitus with arthropathy | 39710007 | 1229552016 |
| 1223148019 | C109J-1 | C109J11 | Insulin treated non-insulin dependent diabetes mellitus | 237599002 | 1223148019 |
| 2674067015 | C10ER | C10ER00 | Latent autoimmune diabetes mellitus in adult | 426875007 | 2674067015 |
| 913441000006119 | C10E0-2 | C10E012 | Insulin-dependent diabetes mellitus with renal complications | 127013003 | 301016 |
| 913471000006110 | C10E1-2 | C10E112 | Insulin-dependent diabetes mellitus with ophthalmic comps | 25093002 | 3688467012 |
| 913501000006115 | C10E2-2 | C10E212 | Insulin-dependent diabetes mellitus with neurological comps | 422088007 | 3688477014 |
| 913531000006111 | C10E3-2 | C10E312 | Insulin dependent diabetes mellitus with multiple complicat | 385041000000108 | 760111000000115 |
| 913591000006110 | C10E5-2 | C10E512 | Skin ulcer associated with diabetes mellitus | 422183001 | 2623058011 |
| 913621000006112 | C10E6-2 | C10E612 | Insulin dependent diabetes mellitus with gangrene | 422275004 | 2616612015 |
| 913651000006115 | C10E7-2 | C10E712 | Insulin dependent diabetes mellitus with retinopathy | 4855003 | 9093013 |
| 913681000006111 | C10E8-2 | C10E812 | Insulin dependent diabetes mellitus - poor control | 268519009 | 401531012 |
| 913711000006112 | C10E9-2 | C10E912 | Insulin dependent diabetes maturity onset | 73211009 | 121589010 |
| 913771000006115 | C10EB-2 | C10EB12 | Insulin dependent diabetes mellitus with mononeuropathy | 230577008 | 345492010 |
| 913801000006118 | C10EC-2 | C10EC12 | Insulin dependent diabetes mellitus with polyneuropathy | 49455004 | 82373015 |
| 913831000006114 | C10ED-2 | C10ED12 | Insulin dependent diabetes mellitus with nephropathy | 127013003 | 301016 |
| 913891000006113 | C10EF-2 | C10EF12 | Insulin dependent diabetes mellitus with diabetic cataract | 43959009 | 73294018 |
| 913921000006119 | C10EG-2 | C10EG12 | Insulin dependent diab mell with peripheral angiopathy | 421895002 | 2618203015 |
| 913981000006115 | C10EJ-2 | C10EJ12 | Insulin dependent diab mell with neuropathic arthropathy | 201724008 | 309740011 |
| 2287971000000110 | C10P | C10P.00 | Diabetes mellitus in remission | 703136005 | 3007272014 |
| 302011 | G73y0 | G73y000 | Diabetic peripheral angiopathy | 127014009 | 302011 |
| 9093013 | F420 | F420.00 | Diabetic retinopathy | 4855003 | 9093013 |
| 19931010 | 66AJ1 | 66AJ100 | Brittle diabetes | 11530004 | 19931010 |
| 73466011 | C1001-2 | C100112 | Non-insulin dependent diabetes mellitus | 44054006 | 493773010 |
| 82373015 | F372-1 | F372.11 | Diabetic polyneuropathy | 49455004 | 82373015 |
| 205225016 | C104-1 | C104.11 | Diabetic nephropathy | 127013003 | 205225016 |
| 264716012 | 66AJz | 66AJz00 | Diabetic - poor control NOS | 268519009 | 401531012 |
| 292466013 | C100 | C100.00 | Diabetes mellitus without complication | 111552007 | 178796018 |
| 292475010 | C100z | C100z00 | Diabetes mellitus NOS with no mention of complication | 111552007 | 178796018 |
| 292496015 | C104z | C104z00 | Diabetes mellitus with nephropathy NOS | 127013003 | 301016 |
| 292565014 | C108z | C108z00 | Diabetes mellitus with multiple complications | 385041000000108 | 760111000000115 |
| 292621010 | C10yz | C10yz00 | Diabetes mellitus NOS with other specified manifestation | 73211009 | 121589010 |
| 293759015 | Cyu23 | Cyu2300 | Diabetic renal disease | 127013003 | 301016 |
| 297758012 | F420z | F420z00 | Diabetic retinopathy NOS | 4855003 | 9093013 |
| 345492010 | F3y0 | F3y0.00 | Diabetic mononeuropathy | 230577008 | 345492010 |
| 354316011 | C314-1 | C314.11 | Kidney disorder due to diabetes mellitus | 127013003 | 3688488015 |
| 401531012 | 66AJ | 66AJ.00 | Diabetic - poor control | 268519009 | 401531012 |
| 411891014 | 66AJ-1 | 66AJ.11 | Unstable diabetes | 11530004 | 19931010 |
| 457328010 | C1099 | C109900 | Non-insulin-dependent diabetes mellitus without complication | 313436004 | 457328010 |
| 459309011 | C109G | C109G00 | Non-insulin dependent diabetes mellitus with arthropathy | 314903002 | 459309011 |
| 493773010 | C109-1 | C109.11 | NIDDM - Non-insulin dependent diabetes mellitus | 44054006 | 493773010 |
| 72651000006114 | C1084 | C108400 | Unstable insulin dependent diabetes mellitus | 11530004 | 19931010 |
| 169731000006118 | 2BBF | 2BBF.00 | Retinal abnormality - diabetes-related | 4855003 | 1230609013 |
| 214921000006116 | F372 | F372.00 | Polyneuropathy in diabetes | 49455004 | 82373015 |
| 223291000000111 | C1001-1 | C100111 | Maturity onset diabetes | 44054006 | 493774016 |
| 280511000006113 | C109D | C109D00 | Non-insulin dependent diabetes mellitus with hypoglyca coma | 719216001 | 3316336015 |
| 280521000006117 | C109A | C109A00 | Non-insulin dependent diabetes mellitus with mononeuropathy | 420436000 | 3697667019 |
| 280531000006119 | C109C | C109C00 | Non-insulin dependent diabetes mellitus with nephropathy | 420279001 | 3035432019 |
| 280541000006112 | C109B | C109B00 | Non-insulin dependent diabetes mellitus with polyneuropathy | 713706002 | 3297353013 |
| 280551000006114 | C1094 | C109400 | Non-insulin-dependent diabetes mellitus with ulcer | 190389009 | 292580015 |
| 280561000006111 | C109F | C109F00 | Non-insulin-dependent diabetes mellitus with peripheral angiopathy | 314902007 | 459307013 |
| 280571000006116 | C109 | C109.00 | Non-insulin dependent diabetes mellitus | 44054006 | 493773010 |
| 280581000006118 | C1093 | C109300 | Non-insulin-dependent diabetes mellitus with multiple complications | 190388001 | 292578014 |
| 280591000006115 | C1092 | C109200 | Non-insulin-dependent diabetes mellitus with neuro comps | 421326000 | 3695406015 |
| 281161000006114 | C109E | C109E00 | Non-insulin depend diabetes mellitus with diabetic cataract | 420756003 | 3035281015 |
| 281181000006116 | C109H | C109H00 | Non-insulin dependent diabetes mellitus with neuropathic arthropathy | 314904008 | 459314010 |
| 377001000006117 | Cyu2 | Cyu2.00 | [X]Diabetes mellitus | 73211009 | 121589010 |
| 587111000006111 | C1091 | C109100 | Non-insulin-dependent diabetes mellitus with ophthalm comps | 422099009 | 3698440016 |
| 587521000006111 | C1090 | C109000 | Non-insulin-dependent diabetes mellitus with renal comps | 420279001 | 2615535015 |
| 616451000006110 | C106-3 | C106.13 | Diabetes mellitus with polyneuropathy | 49455004 | 82373015 |
| 616461000006112 | C104 | C104.00 | Diabetes mellitus with renal manifestation | 127013003 | 301016 |
| 616481000006119 | C1061 | C106100 | Diabetes mellitus, adult onset, + neurological manifestation | 421326000 | 2618196010 |
| 616491000006116 | C1051 | C105100 | Diabetes mellitus, adult onset, + ophthalmic manifestation | 422099009 | 2618233010 |
| 616511000006110 | C1001 | C100100 | Diabetes mellitus, adult onset, no mention of complication | 313436004 | 457329019 |
| 616531000006116 | C1011 | C101100 | Diabetes mellitus, adult onset, with ketoacidosis | 421750000 | 2618220016 |
| 616541000006114 | C1031 | C103100 | Diabetes mellitus, adult onset, with ketoacidotic coma | 421847006 | 2618218019 |
| 616551000006111 | C1041 | C104100 | Diabetes mellitus, adult onset, with renal manifestation | 420279001 | 2615535015 |
| 616561000006113 | C1072 | C107200 | Diabetes mellitus, adult with gangrene | 421631007 | 2618206011 |
| 616571000006118 | C10y1 | C10y100 | Diabetes mellitus, adult, + other specified manifestation | 73211009 | 121589010 |
| 616591000006117 | C1070 | C107000 | Diabetes mellitus, juvenile +peripheral circulatory disorder | 421365002 | 2618205010 |
| 616601000006113 | C1050 | C105000 | Diabetes mellitus, juvenile type, + ophthalmic manifestation | 739681000 | 3537386015 |
| 616641000006110 | C1010 | C101000 | Diabetes mellitus, juvenile type, with ketoacidosis | 420270002 | 2618216015 |
| 616651000006112 | C1030 | C103000 | Diabetes mellitus, juvenile type, with ketoacidotic coma | 421075007 | 2618217012 |
| 616661000006114 | C1040 | C104000 | Diabetes mellitus, juvenile type, with renal manifestation | 421893009 | 2618211013 |
| 616671000006119 | C1060 | C106000 | Diabetes mellitus, juvenile, + neurological manifestation | 421468001 | 2618197018 |
| 616681000006116 | C10y0 | C10y000 | Diabetes mellitus, juvenile, + other specified manifestation | 73211009 | 121589010 |
| 616971000006114 | N0301 | N030100 | Diabetic Charcot's arthropathy | 201724008 | 309739014 |
| 641581000006115 | C1096 | C109600 | Non-insulin-dependent diabetes mellitus with retinopathy | 422034002 | 3035295017 |
| 746791000006111 | C108A | C108A00 | Insulin-dependent diabetes without complication | 111552007 | 178796018 |
| 771371000006118 | C108B | C108B00 | Insulin dependent diabetes mellitus with mononeuropathy | 230577008 | 345492010 |
| 771381000006115 | C1083 | C108300 | Insulin dependent diabetes mellitus with multiple complicatn | 385041000000108 | 760111000000115 |
| 771391000006117 | C108D | C108D00 | Insulin dependent diabetes mellitus with nephropathy | 127013003 | 301016 |
| 771401000006115 | C108C | C108C00 | Insulin dependent diabetes mellitus with polyneuropathy | 49455004 | 82373015 |
| 771411000006117 | C1087 | C108700 | Insulin dependent diabetes mellitus with retinopathy | 4855003 | 9093013 |
| 771501000006119 | C1080 | C108000 | Insulin-dependent diabetes mellitus with renal complications | 127013003 | 301016 |
| 772131000006111 | C108J | C108J00 | Insulin dependent diab mell with neuropathic arthropathy | 201724008 | 309740011 |
| 772151000006116 | C1089 | C108900 | Insulin dependent diabetes maturity onset | 73211009 | 121589010 |
| 772161000006119 | C1000-1 | C100011 | Insulin dependent diabetes mellitus | 73211009 | 121589010 |
| 772171000006114 | C108 | C108.00 | Insulin dependent diabetes mellitus | 73211009 | 121589010 |
| 772181000006112 | C1088 | C108800 | Insulin dependent diabetes mellitus - poor control | 268519009 | 401531012 |
| 787111000006112 | C108-1 | C108.11 | IDDM-Insulin dependent diabetes mellitus | 73211009 | 121589010 |
| 881441000006111 | C1000-99 | C100099 | Diabetes mellitus - juvenile | 73211009 | 881441000006111 |
| 881481000006117 | C104-99 | C104.99 | Diabetes + nephropathy | 127013003 | 881481000006117 |

**Abbreviations**

CPRD = Clinical Practice Research Datalink.

#### **Supplementary table S3. List of oral hypoglycaemic agents DM+D/Product code list included in CPRD prescriptions as validation**

| **ProdCodeId** | **dmdid** | **Term from EMIS** | **ProductName** | | | | **Formulation** | **DrugSubstanceName** | | **SubstanceStrength** | | **BNFChapter** |
| --- | --- | --- | --- | --- | --- | --- | --- | --- | --- | --- | --- | --- |
| 11441000033114 | 42080211000001104 | Acarbose 100mg tablets | Acarbose 100mg tablets | | | | Tablet/ Oral Tablet | Acarbose | | 100.000mg | | 6010203 |
| 11541000033110 | 42080311000001104 | Acarbose 50mg tablets | Acarbose 50mg tablets | | | | Tablet/ Oral Tablet | Acarbose | | 50.000mg | | 6010203 |
| 58841000033113 | 938211000001105 | Amaryl 2mg tablets (Zentiva Pharma UK Ltd) | Amaryl 2mg tablets | | | | Tablet/ Oral Tablet | Glimepiride | | 2.000mg | |  |
| 59541000033116 | 228111000001101 | Amaryl 1mg tablets (Zentiva Pharma UK Ltd) | Amaryl 1mg tablets | | | | Tablet/ Oral Tablet | Glimepiride | | 1.000mg | |  |
| 59641000033115 | 164511000001108 | Amaryl 3mg tablets (Zentiva Pharma UK Ltd) | Amaryl 3mg tablets | | | | Tablet/ Oral Tablet | Glimepiride | | 3.000mg | |  |
| 59741000033112 | 259611000001101 | Amaryl 4mg tablets (Zentiva Pharma UK Ltd) | Amaryl 4mg tablets | | | | Tablet/ Oral Tablet | Glimepiride | | 4.000mg | |  |
| 249441000033114 | 42081111000001104 | Chlorpropamide 100mg tablets | Chlorpropamide 100mg tablets | | | | Tablet/ Oral Tablet | Chlorpropamide | | 100.000mg | | 6010201 |
| 249541000033110 | 42081211000001104 | Chlorpropamide 250mg tablets | Chlorpropamide 250mg tablets | | | | Tablet/ Oral Tablet | Chlorpropamide | | 250.000mg | | 6010201 |
| 406841000033114 | 234011000001108 | Daonil 5mg tablets (Sanofi) | Daonil 5mg tablets | | | | Tablet/ Oral Tablet | Glibenclamide | | 5.000mg | |  |
| 462041000033119 | 13511000001103 | Diabetamide 2.5mg tablets (Ashbourne Pharmaceuticals Ltd) | Diabetamide 2.5mg tablets | | | | Tablet/ Oral Tablet | Glibenclamide | | 2.500mg | |  |
| 462141000033115 | 287811000001101 | Diabetamide 5mg tablets (Ashbourne Pharmaceuticals Ltd) | Diabetamide 5mg tablets | | | | Tablet/ Oral Tablet | Glibenclamide | | 5.000mg | |  |
| 463541000033118 | 745911000001103 | Diamicron 80mg tablets (Servier Laboratories Ltd) | Diamicron 80mg tablets | | | | Tablet/ Oral Tablet | Gliclazide | | 80.000mg | |  |
| 557241000033113 | 233411000001102 | Euglucon 2.5mg tablets (Aventis Pharma) | Euglucon 2.5mg tablets | | | | Tablet/ Oral Tablet | Glibenclamide | | 2.500mg | |  |
| 557341000033115 | 322511000001103 | Euglucon 5mg tablets (Sanofi) | Euglucon 5mg tablets | | | | Tablet/ Oral Tablet | Glibenclamide | | 5.000mg | |  |
| 644541000033111 | 42082711000001104 | Glibenclamide 2.5mg tablets | Glibenclamide 2.5mg tablets | | | | Tablet/ Oral Tablet | Glibenclamide | | 2.500mg | | 6010201 |
| 644641000033112 | 42082811000001104 | Glibenclamide 5mg tablets | Glibenclamide 5mg tablets | | | | Tablet/ Oral Tablet | Glibenclamide | | 5.000mg | | 6010201 |
| 644741000033115 | 515111000001108 | Glibenese 5mg tablets (Pfizer Ltd) | Glibenese 5mg tablets | | | | Tablet/ Oral Tablet | Glipizide | | 5.000mg | |  |
| 644841000033113 | 42083511000001104 | Glipizide 5mg tablets | Glipizide 5mg tablets | | | | Tablet/ Oral Tablet | Glipizide | | 5.000mg | | 6010201 |
| 644941000033117 | 363211000001102 | Glucophage 500mg tablets (Merck Serono Ltd) | Glucophage 500mg tablets | | | | Tablet/ Oral Tablet | Metformin hydrochloride | | 500.000mg | |  |
| 645041000033117 | 365111000001109 | Glucophage 850mg tablets (Merck Serono Ltd) | Glucophage 850mg tablets | | | | Tablet/ Oral Tablet | Metformin hydrochloride | | 850.000mg | |  |
| 645241000033113 | 444311000001106 | Glucobay 100mg tablets (Bayer Plc) | Glucobay 100mg tablets | | | | Tablet/ Oral Tablet | Acarbose | | 100.000mg | |  |
| 645341000033115 | 781411000001104 | Glucobay 50mg tablets (Bayer Plc) | Glucobay 50mg tablets | | | | Tablet/ Oral Tablet | Acarbose | | 50.000mg | |  |
| 645841000033112 | 42083111000001104 | Glimepiride 2mg tablets | Glimepiride 2mg tablets | | | | Tablet/ Oral Tablet | Glimepiride | | 2.000mg | | 6010201 |
| 645941000033116 | 42083011000001104 | Glimepiride 1mg tablets | Glimepiride 1mg tablets | | | | Tablet/ Oral Tablet | Glimepiride | | 1.000mg | | 6010201 |
| 646041000033114 | 42083211000001104 | Glimepiride 3mg tablets | Glimepiride 3mg tablets | | | | Tablet/ Oral Tablet | Glimepiride | | 3.000mg | | 6010201 |
| 646141000033113 | 42083311000001104 | Glimepiride 4mg tablets | Glimepiride 4mg tablets | | | | Tablet/ Oral Tablet | Glimepiride | | 4.000mg | | 6010201 |
| 646441000033117 | 42082911000001104 | Gliclazide 80mg tablets | Gliclazide 80mg tablets | | | | Tablet/ Oral Tablet | Gliclazide | | 80.000mg | | 6010201 |
| 646541000033116 | 42083411000001104 | Glipizide 2.5mg tablets | Glipizide 2.5mg tablets | | | | Tablet/ Oral Tablet | Glipizide | | 2.500mg | | 6010201 |
| 646841000033119 | 3763311000001100 | Glurenorm 30mg tablets (Sanofi) | Glurenorm 30mg tablets | | | | Tablet/ Oral Tablet | Gliquidone | | 30.000mg | |  |
| 646941000033110 | 42083611000001104 | Gliquidone 30mg tablets | Gliquidone 30mg tablets | | | | Tablet/ Oral Tablet | Gliquidone | | 30.000mg | | 6010201 |
| 896941000033112 | 42084911000001104 | Metformin 500mg tablets | Metformin 500mg tablets | | | | Tablet/ Oral Tablet | Metformin hydrochloride | | 500.000mg | | 6010202 |
| 897041000033113 | 42085111000001104 | Metformin 850mg tablets | Metformin 850mg tablets | | | | Tablet/ Oral Tablet | Metformin hydrochloride | | 850.000mg | | 6010202 |
| 920141000033112 | 808711000001104 | Minodiab 2.5mg tablets (Pfizer Ltd) | Minodiab 2.5mg tablets | | | | Tablet/ Oral Tablet | Glipizide | | 2.500mg | |  |
| 920241000033117 | 652411000001107 | Minodiab 5mg tablets (Pfizer Ltd) | Minodiab 5mg tablets | | | | Tablet/ Oral Tablet | Glipizide | | 5.000mg | |  |
| 1278041000033110 | 140711000001103 | Semi-Daonil 2.5mg tablets (Sanofi) | Semi-Daonil 2.5mg tablets | | | | Tablet/ Oral Tablet | Glibenclamide | | 2.500mg | |  |
| 1450941000033110 | 42090611000001104 | Tolbutamide 500mg tablets | Tolbutamide 500mg tablets | | | | Tablet/ Oral Tablet | Tolbutamide | | 500.000mg | | 6010201 |
| 1602941000033110 | 49111000001103 | Diaglyk 80mg tablets (Ashbourne Pharmaceuticals Ltd) | Diaglyk 80mg tablets | | | | Tablet/ Oral Tablet | Gliclazide | | 80.000mg | |  |
| 1659941000033110 | 152311000001101 | NovoNorm 500microgram tablets (Novo Nordisk Ltd) | NovoNorm 500microgram tablets | | | | Tablet/ Oral Tablet | Repaglinide | | 500.000microgram | |  |
| 1660041000033110 | 494111000001103 | NovoNorm 1mg tablets (Novo Nordisk Ltd) | NovoNorm 1mg tablets | | | | Tablet/ Oral Tablet | Repaglinide | | 1.000mg | |  |
| 1660141000033110 | 840811000001100 | NovoNorm 2mg tablets (Novo Nordisk Ltd) | NovoNorm 2mg tablets | | | | Tablet/ Oral Tablet | Repaglinide | | 2.000mg | |  |
| 1672741000033110 | 42088111000001104 | Repaglinide 1mg tablets | Repaglinide 1mg tablets | | | | Tablet/ Oral Tablet | Repaglinide | | 1.000mg | | 6010203 |
| 1672841000033110 | 42088211000001104 | Repaglinide 2mg tablets | Repaglinide 2mg tablets | | | | Tablet/ Oral Tablet | Repaglinide | | 2.000mg | | 6010203 |
| 1672941000033110 | 42088311000001104 | Repaglinide 500microgram tablets | Repaglinide 500microgram tablets | | | | Tablet/ Oral Tablet | Repaglinide | | 500.000microgram | | 6010203 |
| 2147541000033110 | 42089111000001104 | Rosiglitazone 4mg tablets | Rosiglitazone 4mg tablets | | | | Tablet/ Oral Tablet | Rosiglitazone maleate | | 4.000mg | | 6010203 |
| 2147641000033110 | 42089211000001104 | Rosiglitazone 8mg tablets | Rosiglitazone 8mg tablets | | | | Tablet/ Oral Tablet | Rosiglitazone maleate | | 8.000mg | | 6010203 |
| 2147741000033110 | 922211000001106 | Avandia 4mg tablets (GlaxoSmithKline UK Ltd) | Avandia 4mg tablets | | | | Tablet/ Oral Tablet | Rosiglitazone maleate | | 4.000mg | |  |
| 2147841000033110 | 270611000001105 | Avandia 8mg tablets (GlaxoSmithKline UK Ltd) | Avandia 8mg tablets | | | | Tablet/ Oral Tablet | Rosiglitazone maleate | | 8.000mg | |  |
| 2190741000033110 | 42086211000001104 | Pioglitazone 15mg tablets | Pioglitazone 15mg tablets | | | | Tablet/ Oral Tablet | Pioglitazone hydrochloride | | 15.000mg | | 6010203 |
| 2190841000033110 | 42086311000001104 | Pioglitazone 30mg tablets | Pioglitazone 30mg tablets | | | | Tablet/ Oral Tablet | Pioglitazone hydrochloride | | 30.000mg | | 6010203 |
| 2190941000033110 | 58011000001106 | Actos 15mg tablets (Neon Healthcare Ltd) | Actos 15mg tablets | | | | Tablet/ Oral Tablet | Pioglitazone hydrochloride | | 15.000mg | |  |
| 2191041000033110 | 446711000001101 | Actos 30mg tablets (Neon Healthcare Ltd) | Actos 30mg tablets | | | | Tablet/ Oral Tablet | Pioglitazone hydrochloride | | 30.000mg | |  |
| 2288241000033110 | 42085911000001104 | Nateglinide 60mg tablets | Nateglinide 60mg tablets | | | | Tablet/ Oral Tablet | Nateglinide | | 60.000mg | | 6010203 |
| 2288341000033110 | 42085711000001104 | Nateglinide 120mg tablets | Nateglinide 120mg tablets | | | | Tablet/ Oral Tablet | Nateglinide | | 120.000mg | | 6010203 |
| 2288441000033110 | 42085811000001104 | Nateglinide 180mg tablets | Nateglinide 180mg tablets | | | | Tablet/ Oral Tablet | Nateglinide | | 180.000mg | | 6010203 |
| 2288541000033110 | 3650611000001100 | Starlix 60mg tablets (Novartis Pharmaceuticals UK Ltd) | Starlix 60mg tablets | | | | Tablet/ Oral Tablet | Nateglinide | | 60.000mg | |  |
| 2288641000033110 | 3651211000001100 | Starlix 120mg tablets (Novartis Pharmaceuticals UK Ltd) | Starlix 120mg tablets | | | | Tablet/ Oral Tablet | Nateglinide | | 120.000mg | |  |
| 2288741000033110 | 3883511000001100 | Starlix 180mg tablets (Novartis Pharmaceuticals UK Ltd) | Starlix 180mg tablets | | | | Tablet/ Oral Tablet | Nateglinide | | 180.000mg | |  |
| 2289441000033110 | 38896211000001104 | Gliclazide 30mg modified-release tablets | Gliclazide 30mg modified-release tablets | | | | Modified-release tablet | Gliclazide | | 30.000mg | | 6010201 |
| 2289941000033110 | 3661311000001100 | Diamicron 30mg MR tablets (Servier Laboratories Ltd) | Diamicron 30mg MR tablets | | | | Modified-release tablet | Gliclazide | | 30.000mg | |  |
| 2620241000033110 | 8664411000001100 | Metformin 500mg/5ml oral suspension | Metformin 500mg/5ml oral suspension | | | | Oral suspension | Metformin hydrochloride | | 100.000mg/1.000ml | | 6010202 |
| 2995141000033110 | 5302911000001100 | Avandamet 1mg/500mg tablets (GlaxoSmithKline UK Ltd) | Avandamet 1mg/500mg tablets | | | | Tablet/ Oral Tablet | Metformin hydrochloride/ Rosiglitazone maleate | | 500.000mg + 1.000mg | |  |
| 2995241000033110 | 5303611000001100 | Avandamet 2mg/500mg tablets (GlaxoSmithKline UK Ltd) | Avandamet 2mg/500mg tablets | | | | Tablet/ Oral Tablet | Metformin hydrochloride/ Rosiglitazone maleate | | 500.000mg + 2.000mg | |  |
| 2995941000033110 | 42086411000001104 | Pioglitazone 45mg tablets | Pioglitazone 45mg tablets | | | | Tablet/ Oral Tablet | Pioglitazone hydrochloride | | 45.000mg | | 6010203 |
| 2996041000033110 | 5199411000001100 | Actos 45mg tablets (Neon Healthcare Ltd) | Actos 45mg tablets | | | | Tablet/ Oral Tablet | Pioglitazone hydrochloride | | 45.000mg | |  |
| 3191341000033110 | 42088711000001104 | Rosiglitazone 1mg / Metformin 500mg tablets | Rosiglitazone 1mg / Metformin 500mg tablets | | | | Tablet/ Oral Tablet | Metformin hydrochloride/ Rosiglitazone maleate | | 500.000mg + 1.000mg | | 6010203 |
| 3191441000033110 | 42088911000001104 | Rosiglitazone 2mg / Metformin 500mg tablets | Rosiglitazone 2mg / Metformin 500mg tablets | | | | Tablet/ Oral Tablet | Metformin hydrochloride/ Rosiglitazone maleate | | 500.000mg + 2.000mg | | 6010203 |
| 3200541000033110 | 42088811000001104 | Rosiglitazone 2mg / Metformin 1g tablets | Rosiglitazone 2mg / Metformin 1g tablets | | | | Tablet/ Oral Tablet | Metformin hydrochloride/ Rosiglitazone maleate | | 1.000gram + 2.000mg | | 6010203 |
| 3200641000033110 | 42089011000001104 | Rosiglitazone 4mg / Metformin 1g tablets | Rosiglitazone 4mg / Metformin 1g tablets | | | | Tablet/ Oral Tablet | Metformin hydrochloride/ Rosiglitazone maleate | | 1.000gram + 4.000mg | | 6010203 |
| 3200741000033110 | 8176311000001100 | Avandamet 2mg/1000mg tablets (GlaxoSmithKline UK Ltd) | Avandamet 2mg/1000mg tablets | | | | Tablet/ Oral Tablet | Metformin hydrochloride/ Rosiglitazone maleate | | 1.000gram + 2.000mg | |  |
| 3200841000033110 | 8174611000001100 | Avandamet 4mg/1000mg tablets (GlaxoSmithKline UK Ltd) | Avandamet 4mg/1000mg tablets | | | | Tablet/ Oral Tablet | Metformin hydrochloride/ Rosiglitazone maleate | | 1.000gram + 4.000mg | |  |
| 3228141000033110 | 39113511000001104 | Metformin 500mg modified-release tablets | Metformin 500mg modified-release tablets | | | | Modified-release tablet | Metformin hydrochloride | | 500.000mg | | 6010202 |
| 3228241000033110 | 8990711000001100 | Glucophage SR 500mg tablets (Merck Serono Ltd) | Glucophage SR 500mg tablets | | | | Modified-release tablet | Metformin hydrochloride | | 500.000mg | |  |
| 3953041000033110 | 8524311000001100 | Gliclazide 80mg/5ml oral suspension | Gliclazide 80mg/5ml oral suspension | | | | Oral suspension | Gliclazide | | 16.000mg/1.000ml | | 6010201 |
| 3982341000033110 | 10750111000001100 | Metformin 500mg/5ml oral solution sugar free | Metformin 500mg/5ml oral solution sugar free | | | | Oral solution | Metformin hydrochloride | | 100.000mg/1.000ml | | 6010202 |
| 3982441000033110 | 10741611000001100 | Metsol 500mg/5ml oral solution (Kappin Ltd) | Metsol 500mg/5ml oral solution | | | | Oral solution | Metformin hydrochloride | | 100.000mg/1.000ml | |  |
| 3983941000033110 | 42086111000001104 | Pioglitazone 15mg / Metformin 850mg tablets | Pioglitazone 15mg / Metformin 850mg tablets | | | | Tablet/ Oral Tablet | Metformin hydrochloride/ Pioglitazone hydrochloride | | 850.000mg + 15.000mg | | 6010203 |
| 3984241000033110 | 10922311000001100 | Competact 15mg/850mg tablets (Neon Healthcare Ltd) | Competact 15mg/850mg tablets | | | | Tablet/ Oral Tablet | Metformin hydrochloride/ Pioglitazone hydrochloride | | 850.000mg + 15.000mg | |  |
| 3993941000033110 | 10952411000001100 | Prandin 0.5mg tablets (Novo Nordisk Ltd) | Prandin 0.5mg tablets | | | | Tablet/ Oral Tablet | Repaglinide | | 500.000microgram | |  |
| 3994041000033110 | 10956911000001100 | Prandin 1mg tablets (Novo Nordisk Ltd) | Prandin 1mg tablets | | | | Tablet/ Oral Tablet | Repaglinide | | 1.000mg | |  |
| 3994141000033110 | 10957811000001100 | Prandin 2mg tablets (Novo Nordisk Ltd) | Prandin 2mg tablets | | | | Tablet/ Oral Tablet | Repaglinide | | 2.000mg | |  |
| 4133241000033110 | 42089711000001104 | Sitagliptin 100mg tablets | Sitagliptin 100mg tablets | | | | Tablet/ Oral Tablet | Sitagliptin | | 100.000mg | | 6010203 |
| 4133341000033110 | 11473711000001100 | Januvia 100mg tablets (Merck Sharp & Dohme (UK) Ltd) | Januvia 100mg tablets | | | | Tablet/ Oral Tablet | Sitagliptin | | 100.000mg | |  |
| 4269741000033110 | 13412911000001100 | Vildagliptin 50mg tablets | Vildagliptin 50mg tablets | | | | Tablet/ Oral Tablet | Vildagliptin | | 50.000mg | | 6010203 |
| 4269941000033110 | 13411611000001100 | Galvus 50mg tablets (Novartis Pharmaceuticals UK Ltd) | Galvus 50mg tablets | | | | Tablet/ Oral Tablet | Vildagliptin | | 50.000mg | |  |
| 4452341000033110 | 13413111000001100 | Vildagliptin 50mg / Metformin 850mg tablets | Vildagliptin 50mg / Metformin 850mg tablets | | | | Tablet/ Oral Tablet | Metformin hydrochloride/ Vildagliptin | | 850.000mg + 50.000mg | | 6010203 |
| 4452441000033110 | 13413011000001100 | Vildagliptin 50mg / Metformin 1g tablets | Vildagliptin 50mg / Metformin 1g tablets | | | | Tablet/ Oral Tablet | Metformin hydrochloride/ Vildagliptin | | 1.000gram + 50.000mg | | 6010203 |
| 4452541000033110 | 13412311000001100 | Eucreas 50mg/1000mg tablets (Novartis Pharmaceuticals UK Ltd) | Eucreas 50mg/1000mg tablets | | | | Tablet/ Oral Tablet | Metformin hydrochloride/ Vildagliptin | | 1.000gram + 50.000mg | |  |
| 4452641000033110 | 13412611000001100 | Eucreas 50mg/850mg tablets (Novartis Pharmaceuticals UK Ltd) | Eucreas 50mg/850mg tablets | | | | Tablet/ Oral Tablet | Metformin hydrochloride/ Vildagliptin | | 850.000mg + 50.000mg | |  |
| 4549141000033110 | 38893811000001104 | Metformin 750mg modified-release tablets | Metformin 750mg modified-release tablets | | | | Modified-release tablet | Metformin hydrochloride | | 750.000mg | | 6010202 |
| 4549241000033110 | 13748611000001100 | Glucophage SR 750mg tablets (Merck Serono Ltd) | Glucophage SR 750mg tablets | | | | Modified-release tablet | Metformin hydrochloride | | 750.000mg | |  |
| 4941241000033110 | 13456711000001100 | Nazdol MR 30mg tablets (Krka UK Ltd) | Nazdol MR 30mg tablets | | | | Modified-release tablet | Gliclazide | | 30.000mg | |  |
| 4945241000033110 | 38893711000001104 | Metformin 1g modified-release tablets | Metformin 1g modified-release tablets | | | | Modified-release tablet | Metformin hydrochloride | | 1.000gram | | 6010202 |
| 4945341000033110 | 15367811000001100 | Glucophage SR 1000mg tablets (Merck Serono Ltd) | Glucophage SR 1000mg tablets | | | | Modified-release tablet | Metformin hydrochloride | | 1.000gram | |  |
| 4957741000033110 | 14183911000001100 | Bolamyn SR 500mg tablets (Teva UK Ltd) | Bolamyn SR 500mg tablets | | | | Modified-release tablet | Metformin hydrochloride | | 500.000mg | |  |
| 5007641000033110 | 15411311000001100 | Metformin 500mg oral powder sachets sugar free | Metformin 500mg oral powder sachets sugar free | | | | Powder for oral solution/ Powder | Metformin hydrochloride | | 500.000mg | | 6010202 |
| 5007741000033110 | 15411211000001100 | Metformin 1g oral powder sachets sugar free | Metformin 1g oral powder sachets sugar free | | | | Powder for oral solution/ Powder | Metformin hydrochloride | | 1.000gram | | 6010202 |
| 5007841000033110 | 15373711000001100 | Glucophage 500mg oral powder sachets (Merck Serono Ltd) | Glucophage 500mg oral powder sachets | | | | Powder for oral solution/ Powder | Metformin hydrochloride | | 500.000mg | |  |
| 5007941000033110 | 15374311000001100 | Glucophage 1000mg oral powder sachets (Merck Serono Ltd) | Glucophage 1000mg oral powder sachets | | | | Powder for oral solution/ Powder | Metformin hydrochloride | | 1.000gram | |  |
| 5131241000033110 | 15859111000001100 | Liraglutide 6mg/ml solution for injection 3ml pre-filled disposable devices | Liraglutide 6mg/ml solution for injection 3ml pre-filled disposable devices | | | | Solution for injection | Liraglutide | | 6.000mg/1.000ml | | 6010203 |
| 5131341000033110 | 15858611000001100 | Victoza 6mg/ml solution for injection 3ml pre-filled pens (Novo Nordisk Ltd) | Victoza 6mg/ml solution for injection 3ml pre-filled pens | | | | Solution for injection | Liraglutide | | 6.000mg/1.000ml | |  |
| 5132241000033110 | 17060511000001100 | Janumet 50mg/1000mg tablets (Merck Sharp & Dohme (UK) Ltd) | Janumet 50mg/1000mg tablets | | | | Tablet/ Oral Tablet | Metformin hydrochloride/ Sitagliptin | | 1.000gram + 50.000mg | |  |
| 5316641000033110 | 39703911000001104 | Saxagliptin 5mg tablets | Saxagliptin 5mg tablets | | | | Tablet/ Oral Tablet | Saxagliptin hydrochloride | | 5.000mg | | 6010203 |
| 5316741000033110 | 15993011000001100 | Onglyza 5mg tablets (AstraZeneca UK Ltd) | Onglyza 5mg tablets | | | | Tablet/ Oral Tablet | Saxagliptin hydrochloride | | 5.000mg | |  |
| 5378141000033110 | 16536211000001100 | Dacadis MR 30mg tablets (Viatris UK Healthcare Ltd) | Dacadis MR 30mg tablets | | | | Modified-release tablet | Gliclazide | | 30.000mg | |  |
| 5576041000033110 | 17071811000001100 | Metformin 1g / Sitagliptin 50mg tablets | Metformin 1g / Sitagliptin 50mg tablets | | | | Tablet/ Oral Tablet | Metformin hydrochloride/ Sitagliptin | | 1.000gram + 50.000mg | | 6010203 |
| 5709741000033110 | 16702011000001100 | Gliclazide 40mg tablets | Gliclazide 40mg tablets | | | | Tablet/ Oral Tablet | Gliclazide | | 40.000mg | | 6010201 |
| 5709841000033110 | 16677511000001100 | Zicron 40mg tablets (Bristol Laboratories Ltd) | Zicron 40mg tablets | | | | Tablet/ Oral Tablet | Gliclazide | | 40.000mg | |  |
| 5815341000033110 | 15334511000001100 | Edicil MR 30mg tablets (Teva UK Ltd) | Edicil MR 30mg tablets | | | | Modified-release tablet | Gliclazide | | 30.000mg | |  |
| 5869241000033110 | 8524211000001100 | Gliclazide 40mg/5ml oral suspension | Gliclazide 40mg/5ml oral suspension | | | | Oral suspension | Gliclazide | | 8.000mg/1.000ml | | 6010201 |
| 5968741000033110 | 8523711000001100 | Glibenclamide 5mg/5ml oral solution | Glibenclamide 5mg/5ml oral solution | | | | Oral solution | Glibenclamide | | 1.000mg/1.000ml | | 6010201 |
| 5968841000033110 | 8523811000001100 | Glibenclamide 5mg/5ml oral suspension | Glibenclamide 5mg/5ml oral suspension | | | | Oral suspension | Glibenclamide | | 1.000mg/1.000ml | | 6010201 |
| 5997041000033110 | 42085011000001104 | Metformin 500mg/5ml oral solution | Metformin 500mg/5ml oral solution | | | | Oral solution | Metformin hydrochloride | | 100.000mg/1.000ml | | 6010202 |
| 6029841000033110 | 18141911000001100 | Metabet SR 500mg tablets (Morningside Healthcare Ltd) | Metabet SR 500mg tablets | | | | Modified-release tablet | Metformin hydrochloride | | 500.000mg | |  |
| 6123241000033110 | 39703811000001104 | Saxagliptin 2.5mg tablets | Saxagliptin 2.5mg tablets | | | | Tablet/ Oral Tablet | Saxagliptin hydrochloride | | 2.500mg | | 6010203 |
| 6123341000033110 | 18596311000001100 | Onglyza 2.5mg tablets (AstraZeneca UK Ltd) | Onglyza 2.5mg tablets | | | | Tablet/ Oral Tablet | Saxagliptin hydrochloride | | 2.500mg | |  |
| 6279741000033110 | 18885611000001100 | Metabet SR 1000mg tablets (Morningside Healthcare Ltd) | Metabet SR 1000mg tablets | | | | Modified-release tablet | Metformin hydrochloride | | 1.000gram | |  |
| 6391041000033110 | 19308111000001100 | Glucient SR 500mg tablets (Consilient Health Ltd) | Glucient SR 500mg tablets | | | | Modified-release tablet | Metformin hydrochloride | | 500.000mg | |  |
| 6391141000033110 | 19306711000001100 | Enyglid 0.5mg tablets (Krka UK Ltd) | Enyglid 0.5mg tablets | | | | Tablet/ Oral Tablet | Repaglinide | | 500.000microgram | |  |
| 6391341000033110 | 19307311000001100 | Enyglid 2mg tablets (Krka UK Ltd) | Enyglid 2mg tablets | | | | Tablet/ Oral Tablet | Repaglinide | | 2.000mg | |  |
| 6444041000033110 | 19525211000001100 | Linagliptin 5mg tablets | Linagliptin 5mg tablets | | | | Tablet/ Oral Tablet | Linagliptin | | 5.000mg | | 6010203 |
| 6444141000033110 | 19492811000001100 | Trajenta 5mg tablets (Boehringer Ingelheim Ltd) | Trajenta 5mg tablets | | | | Tablet/ Oral Tablet | Linagliptin | | 5.000mg | |  |
| 6526041000033110 | 20023111000001100 | Glizofar 30mg tablets (Arrow Generics Ltd) | Glizofar 30mg tablets | | | | Tablet/ Oral Tablet | Pioglitazone hydrochloride | | 30.000mg | |  |
| 7687241000033110 | 42089911000001104 | Sitagliptin 50mg tablets | Sitagliptin 50mg tablets | | | | Tablet/ Oral Tablet | Sitagliptin | | 50.000mg | | 6010203 |
| 7687341000033110 | 20114811000001100 | Januvia 50mg tablets (Merck Sharp & Dohme (UK) Ltd) | Januvia 50mg tablets | | | | Tablet/ Oral Tablet | Sitagliptin | | 50.000mg | |  |
| 7687441000033110 | 42089811000001104 | Sitagliptin 25mg tablets | Sitagliptin 25mg tablets | | | | Tablet/ Oral Tablet | Sitagliptin | | 25.000mg | | 6010203 |
| 7687541000033110 | 20115111000001100 | Januvia 25mg tablets (Merck Sharp & Dohme (UK) Ltd) | Januvia 25mg tablets | | | | Tablet/ Oral Tablet | Sitagliptin | | 25.000mg | |  |
| 7874941000033110 | 20552511000001100 | Diagemet XL 500mg tablets (Genus Pharmaceuticals Ltd) | Diagemet XL 500mg tablets | | | | Modified-release tablet | Metformin hydrochloride | | 500.000mg | |  |
| 8115741000033110 | 21245011000001100 | Linagliptin 2.5mg / Metformin 1g tablets | Linagliptin 2.5mg / Metformin 1g tablets | | | | Tablet/ Oral Tablet | Linagliptin/ Metformin hydrochloride | | 2.500mg + 1000.000mg | | 6010203 |
| 8115841000033110 | 21245111000001100 | Linagliptin 2.5mg / Metformin 850mg tablets | Linagliptin 2.5mg / Metformin 850mg tablets | | | | Tablet/ Oral Tablet | Linagliptin/ Metformin hydrochloride | | 2.500mg + 850.000mg | | 6010203 |
| 8115941000033110 | 21208511000001100 | Jentadueto 2.5mg/1000mg tablets (Boehringer Ingelheim Ltd) | Jentadueto 2.5mg/1000mg tablets | | | | Tablet/ Oral Tablet | Linagliptin/ Metformin hydrochloride | | 2.500mg + 1000.000mg | |  |
| 8116041000033110 | 21208211000001100 | Jentadueto 2.5mg/850mg tablets (Boehringer Ingelheim Ltd) | Jentadueto 2.5mg/850mg tablets | | | | Tablet/ Oral Tablet | Linagliptin/ Metformin hydrochloride | | 2.500mg + 850.000mg | |  |
| 8125241000033110 | 21366911000001100 | Glimepiride 6mg/5ml oral suspension | Glimepiride 6mg/5ml oral suspension | | | | Oral suspension | Glimepiride | | 1.200mg/1.000ml | | 6010201 |
| 8199341000033110 | 39689911000001104 | Dapagliflozin 5mg tablets | Dapagliflozin 5mg tablets | | | | Tablet/ Oral Tablet | Dapagliflozin propanediol monohydrate | | 5.000mg | | 6010203 |
| 8199441000033110 | 39689811000001104 | Dapagliflozin 10mg tablets | Dapagliflozin 10mg tablets | | | | Tablet/ Oral Tablet | Dapagliflozin propanediol monohydrate | | 10.000mg | | 6010203 |
| 8199541000033110 | 21609511000001100 | Forxiga 5mg tablets (AstraZeneca UK Ltd) | Forxiga 5mg tablets | | | | Tablet/ Oral Tablet | Dapagliflozin propanediol monohydrate | | 5.000mg | |  |
| 8199641000033110 | 21609811000001100 | Forxiga 10mg tablets (AstraZeneca UK Ltd) | Forxiga 10mg tablets | | | | Tablet/ Oral Tablet | Dapagliflozin propanediol monohydrate | | 10.000mg | |  |
| 8242541000033110 | 21711511000001100 | Saxagliptin 2.5mg / Metformin 850mg tablets | Saxagliptin 2.5mg / Metformin 850mg tablets | | | | Tablet/ Oral Tablet | Metformin hydrochloride/ Saxagliptin hydrochloride | | 850.000mg + 2.500mg | | 6010203 |
| 8242641000033110 | 21711411000001100 | Saxagliptin 2.5mg / Metformin 1g tablets | Saxagliptin 2.5mg / Metformin 1g tablets | | | | Tablet/ Oral Tablet | Metformin hydrochloride/ Saxagliptin hydrochloride | | 1.000gram + 2.500mg | | 6010203 |
| 8242741000033110 | 21705311000001100 | Komboglyze 2.5mg/850mg tablets (AstraZeneca UK Ltd) | Komboglyze 2.5mg/850mg tablets | | | | Tablet/ Oral Tablet | Metformin hydrochloride/ Saxagliptin hydrochloride | | 850.000mg + 2.500mg | |  |
| 8242841000033110 | 21705611000001100 | Komboglyze 2.5mg/1000mg tablets (AstraZeneca UK Ltd) | Komboglyze 2.5mg/1000mg tablets | | | | Tablet/ Oral Tablet | Metformin hydrochloride/ Saxagliptin hydrochloride | | 1.000gram + 2.500mg | |  |
| 8267341000033110 | 21994611000001100 | Lixisenatide 10micrograms/0.2ml solution for injection 3ml pre-filled disposable devices | Lixisenatide 10micrograms/0.2ml solution for injection 3ml pre-filled disposable devices | | | | Solution for injection | Lixisenatide | | 50.000microgram/1.000ml | | 6010203 |
| 8267441000033110 | 21994811000001100 | Lixisenatide 20micrograms/0.2ml solution for injection 3ml pre-filled disposable devices | Lixisenatide 20micrograms/0.2ml solution for injection 3ml pre-filled disposable devices | | | | Solution for injection | Lixisenatide | | 100.000microgram/1.000ml | | 6010203 |
| 8267541000033110 | 21941511000001100 | Lyxumia 10micrograms/0.2ml solution for injection 3ml pre-filled pens (Sanofi) | Lyxumia 10micrograms/0.2ml solution for injection 3ml pre-filled pens | | | | Solution for injection | Lixisenatide | | 50.000microgram/1.000ml | |  |
| 8267641000033110 | 21941011000001100 | Lyxumia 20micrograms/0.2ml solution for injection 3ml pre-filled pens (Sanofi) | Lyxumia 20micrograms/0.2ml solution for injection 3ml pre-filled pens | | | | Solution for injection | Lixisenatide | | 100.000microgram/1.000ml | |  |
| 8298141000033110 | 22226111000001100 | Gliclazide 60mg modified-release tablets | Gliclazide 60mg modified-release tablets | | | | Modified-release tablet | Gliclazide | | 60.000mg | | 6010201 |
| 8298241000033110 | 22225011000001100 | Laaglyda MR 60mg tablets (Krka UK Ltd) | Laaglyda MR 60mg tablets | | | | Modified-release tablet | Gliclazide | | 60.000mg | |  |
| 8348941000033110 | 22308811000001100 | Bolamyn SR 1000mg tablets (Teva UK Ltd) | Bolamyn SR 1000mg tablets | | | | Modified-release tablet | Metformin hydrochloride | | 1.000gram | |  |
| 8839341000033110 | 23369611000001100 | Glidipion 30mg tablets (Actavis UK Ltd) | Glidipion 30mg tablets | | | | Tablet/ Oral Tablet | Pioglitazone hydrochloride | | 30.000mg | |  |
| 8959241000033110 | 23637511000001100 | Alogliptin 6.25mg tablets | Alogliptin 6.25mg tablets | | | | Tablet/ Oral Tablet | Alogliptin benzoate | | 6.250mg | | 6010203 |
| 8959341000033110 | 23637311000001100 | Alogliptin 12.5mg tablets | Alogliptin 12.5mg tablets | | | | Tablet/ Oral Tablet | Alogliptin benzoate | | 12.500mg | | 6010203 |
| 8959441000033110 | 23637411000001100 | Alogliptin 25mg tablets | Alogliptin 25mg tablets | | | | Tablet/ Oral Tablet | Alogliptin benzoate | | 25.000mg | | 6010203 |
| 8959541000033110 | 23634111000001100 | Vipidia 6.25mg tablets (Takeda UK Ltd) | Vipidia 6.25mg tablets | | | | Tablet/ Oral Tablet | Alogliptin benzoate | | 6.250mg | |  |
| 8959641000033110 | 23636011000001100 | Vipidia 12.5mg tablets (Takeda UK Ltd) | Vipidia 12.5mg tablets | | | | Tablet/ Oral Tablet | Alogliptin benzoate | | 12.500mg | |  |
| 8959741000033110 | 23636311000001100 | Vipidia 25mg tablets (Takeda UK Ltd) | Vipidia 25mg tablets | | | | Tablet/ Oral Tablet | Alogliptin benzoate | | 25.000mg | |  |
| 8959841000033110 | 23637211000001100 | Alogliptin 12.5mg / Metformin 1g tablets | Alogliptin 12.5mg / Metformin 1g tablets | | | | Tablet/ Oral Tablet | Alogliptin benzoate/ Metformin hydrochloride | | 12.500mg + 1.000gram | | 6010203 |
| 8959941000033110 | 23632611000001100 | Vipdomet 12.5mg/1000mg tablets (Takeda UK Ltd) | Vipdomet 12.5mg/1000mg tablets | | | | Tablet/ Oral Tablet | Alogliptin benzoate/ Metformin hydrochloride | | 12.500mg + 1.000gram | |  |
| 9106041000033110 | 24054611000001100 | Dapagliflozin 5mg / Metformin 1g tablets | Dapagliflozin 5mg / Metformin 1g tablets | | | | Tablet/ Oral Tablet | Dapagliflozin propanediol monohydrate/ Metformin hydrochloride | | 5.000mg + 1.000gram | | 6010203 |
| 9106141000033110 | 24054711000001100 | Dapagliflozin 5mg / Metformin 850mg tablets | Dapagliflozin 5mg / Metformin 850mg tablets | | | | Tablet/ Oral Tablet | Dapagliflozin propanediol monohydrate/ Metformin hydrochloride | | 5.000mg + 850.000mg | | 6010203 |
| 9106241000033110 | 24018511000001100 | Xigduo 5mg/1000mg tablets (AstraZeneca UK Ltd) | Xigduo 5mg/1000mg tablets | | | | Tablet/ Oral Tablet | Dapagliflozin propanediol monohydrate/ Metformin hydrochloride | | 5.000mg + 1.000gram | |  |
| 9106341000033110 | 24018111000001100 | Xigduo 5mg/850mg tablets (AstraZeneca UK Ltd) | Xigduo 5mg/850mg tablets | | | | Tablet/ Oral Tablet | Dapagliflozin propanediol monohydrate/ Metformin hydrochloride | | 5.000mg + 850.000mg | |  |
| 9110341000033110 | 39734511000001104 | Canagliflozin 100mg tablets | Canagliflozin 100mg tablets | | | | Tablet/ Oral Tablet | Canagliflozin hemihydrate | | 100.000mg | | 6010203 |
| 9110441000033110 | 24104511000001100 | Canagliflozin 300mg tablets | Canagliflozin 300mg tablets | | | | Tablet/ Oral Tablet | Canagliflozin hemihydrate | | 300.000mg | | 6010203 |
| 9110541000033110 | 24088611000001100 | Invokana 100mg tablets (Napp Pharmaceuticals Ltd) | Invokana 100mg tablets | | | | Tablet/ Oral Tablet | Canagliflozin hemihydrate | | 100.000mg | |  |
| 9110641000033110 | 24088311000001100 | Invokana 300mg tablets (Napp Pharmaceuticals Ltd) | Invokana 300mg tablets | | | | Tablet/ Oral Tablet | Canagliflozin hemihydrate | | 300.000mg | |  |
| 9230641000033110 | 24568411000001100 | Sukkarto SR 500mg tablets (Morningside Healthcare Ltd) | Sukkarto SR 500mg tablets | | | | Modified-release tablet | Metformin hydrochloride | | 500.000mg | |  |
| 9230841000033110 | 24568211000001100 | Sukkarto SR 1000mg tablets (Morningside Healthcare Ltd) | Sukkarto SR 1000mg tablets | | | | Modified-release tablet | Metformin hydrochloride | | 1.000gram | |  |
| 9336641000033110 | 25290511000001100 | Empagliflozin 10mg tablets | Empagliflozin 10mg tablets | | | | Tablet/ Oral Tablet | Empagliflozin | | 10.000mg | | 6010203 |
| 9336841000033110 | 25290611000001100 | Empagliflozin 25mg tablets | Empagliflozin 25mg tablets | | | | Tablet/ Oral Tablet | Empagliflozin | | 25.000mg | | 6010203 |
| 9337141000033110 | 25238811000001100 | Jardiance 10mg tablets (Boehringer Ingelheim Ltd) | Jardiance 10mg tablets | | | | Tablet/ Oral Tablet | Empagliflozin | | 10.000mg | |  |
| 9337241000033110 | 25239711000001100 | Jardiance 25mg tablets (Boehringer Ingelheim Ltd) | Jardiance 25mg tablets | | | | Tablet/ Oral Tablet | Empagliflozin | | 25.000mg | |  |
| 9851541000033110 | 28049211000001100 | Canagliflozin 50mg / Metformin 1g tablets | Canagliflozin 50mg / Metformin 1g tablets | | | | Tablet/ Oral Tablet | Canagliflozin hemihydrate/ Metformin hydrochloride | | 50.000mg + 1.000gram | | 6010203 |
| 9851641000033110 | 28049311000001100 | Canagliflozin 50mg / Metformin 850mg tablets | Canagliflozin 50mg / Metformin 850mg tablets | | | | Tablet/ Oral Tablet | Canagliflozin hemihydrate/ Metformin hydrochloride | | 50.000mg + 850.000mg | | 6010203 |
| 9851741000033110 | 28022511000001100 | Vokanamet 50mg/850mg tablets (Napp Pharmaceuticals Ltd) | Vokanamet 50mg/850mg tablets | | | | Tablet/ Oral Tablet | Canagliflozin hemihydrate/ Metformin hydrochloride | | 50.000mg + 850.000mg | |  |
| 9851841000033110 | 28024411000001100 | Vokanamet 50mg/1000mg tablets (Napp Pharmaceuticals Ltd) | Vokanamet 50mg/1000mg tablets | | | | Tablet/ Oral Tablet | Canagliflozin hemihydrate/ Metformin hydrochloride | | 50.000mg + 1.000gram | |  |
| 10043341000033100 | 28420711000001100 | Vamju 30mg modified-release tablets (Advanz Pharma) | Vamju 30mg modified-release tablets | | | | Modified-release tablet | Gliclazide | | 30.000mg | |  |
| 10043441000033100 | 28420911000001100 | Vamju 60mg modified-release tablets (Advanz Pharma) | Vamju 60mg modified-release tablets | | | | Modified-release tablet | Gliclazide | | 60.000mg | |  |
| 10044541000033100 | 28279611000001100 | Insulin degludec 100units/ml / Liraglutide 3.6mg/ml solution for injection 3ml pre-filled disposable devices | Insulin degludec 100units/ml / Liraglutide 3.6mg/ml solution for injection 3ml pre-filled disposable devices | | | | Solution for injection | Insulin degludec/ Liraglutide | | 100.000unit/1.000ml + 100.000unit/1.000ml + 3.600mg/1.000ml | | 6010203 |
| 10044641000033100 | 28054311000001100 | Xultophy 100units/ml / 3.6mg/ml solution for injection 3ml pre-filled pens (Novo Nordisk Ltd) | Xultophy 100units/ml / 3.6mg/ml solution for injection 3ml pre-filled pens | | | | Solution for injection | Insulin degludec/ Liraglutide | | 100.000unit/1.000ml + 100.000unit/1.000ml + 3.600mg/1.000ml | | |
| 10207841000033100 | 28789611000001100 | Dulaglutide 0.75mg/0.5ml solution for injection pre-filled disposable devices | Dulaglutide 0.75mg/0.5ml solution for injection pre-filled disposable devices | | | | Solution for injection | Dulaglutide | | 1.500mg/1.000ml | | 6010203 |
| 10207941000033100 | 28789711000001100 | Dulaglutide 1.5mg/0.5ml solution for injection pre-filled disposable devices | Dulaglutide 1.5mg/0.5ml solution for injection pre-filled disposable devices | | | | Solution for injection | Dulaglutide | | 3.000mg/1.000ml | | 6010203 |
| 10208041000033100 | 28461011000001100 | Trulicity 0.75mg/0.5ml solution for injection pre-filled pens (Eli Lilly and Company Ltd) | Trulicity 0.75mg/0.5ml solution for injection pre-filled pens | | | | Solution for injection | Dulaglutide | | 1.500mg/1.000ml | |  |
| 10208141000033100 | 28462311000001100 | Trulicity 1.5mg/0.5ml solution for injection pre-filled pens (Eli Lilly and Company Ltd) | Trulicity 1.5mg/0.5ml solution for injection pre-filled pens | | | | Solution for injection | Dulaglutide | | 3.000mg/1.000ml | |  |
| 10251341000033100 | 28940811000001100 | Glidipion 15mg tablets (Actavis UK Ltd) | Glidipion 15mg tablets | | | | Tablet/ Oral Tablet | Pioglitazone hydrochloride | | 15.000mg | |  |
| 10336941000033100 | 29742811000001100 | Diabiom 15mg tablets (Tillomed Laboratories Ltd) | Diabiom 15mg tablets | | | | Tablet/ Oral Tablet | Pioglitazone hydrochloride | | 15.000mg | |  |
| 10337041000033100 | 29743011000001100 | Diabiom 30mg tablets (Tillomed Laboratories Ltd) | Diabiom 30mg tablets | | | | Tablet/ Oral Tablet | Pioglitazone hydrochloride | | 30.000mg | |  |
| 10598841000033100 | 30012111000001100 | Glucient SR 750mg tablets (Consilient Health Ltd) | Glucient SR 750mg tablets | | | | Modified-release tablet | Metformin hydrochloride | | 750.000mg | |  |
| 10598941000033100 | 30012311000001100 | Glucient SR 1000mg tablets (Consilient Health Ltd) | Glucient SR 1000mg tablets | | | | Modified-release tablet | Metformin hydrochloride | | 1.000gram | |  |
| 10614141000033100 | 30318411000001100 | Empagliflozin 5mg / Metformin 850mg tablets | Empagliflozin 5mg / Metformin 850mg tablets | | | | Tablet/ Oral Tablet | Empagliflozin/ Metformin hydrochloride | | 5.000mg + 850.000mg | | 6010203 |
| 10614241000033100 | 30318311000001100 | Empagliflozin 5mg / Metformin 1g tablets | Empagliflozin 5mg / Metformin 1g tablets | | | | Tablet/ Oral Tablet | Empagliflozin/ Metformin hydrochloride | | 5.000mg + 1.000gram | | 6010203 |
| 10614341000033100 | 30318211000001100 | Empagliflozin 12.5mg / Metformin 850mg tablets | Empagliflozin 12.5mg / Metformin 850mg tablets | | | | Tablet/ Oral Tablet | Empagliflozin/ Metformin hydrochloride | | 12.500mg + 850.000mg | | 6010203 |
| 10614441000033100 | 30318111000001100 | Empagliflozin 12.5mg / Metformin 1g tablets | Empagliflozin 12.5mg / Metformin 1g tablets | | | | Tablet/ Oral Tablet | Empagliflozin/ Metformin hydrochloride | | 12.500mg + 1.000gram | | 6010203 |
| 10614541000033100 | 30173411000001100 | Synjardy 5mg/850mg tablets (Boehringer Ingelheim Ltd) | Synjardy 5mg/850mg tablets | | | | Tablet/ Oral Tablet | Empagliflozin/ Metformin hydrochloride | | 5.000mg + 850.000mg | |  |
| 10614641000033100 | 30174111000001100 | Synjardy 5mg/1000mg tablets (Boehringer Ingelheim Ltd) | Synjardy 5mg/1000mg tablets | | | | Tablet/ Oral Tablet | Empagliflozin/ Metformin hydrochloride | | 5.000mg + 1.000gram | |  |
| 10614741000033100 | 30175011000001100 | Synjardy 12.5mg/850mg tablets (Boehringer Ingelheim Ltd) | Synjardy 12.5mg/850mg tablets | | | | Tablet/ Oral Tablet | Empagliflozin/ Metformin hydrochloride | | 12.500mg + 850.000mg | |  |
| 10614841000033100 | 30175711000001100 | Synjardy 12.5mg/1000mg tablets (Boehringer Ingelheim Ltd) | Synjardy 12.5mg/1000mg tablets | | | | Tablet/ Oral Tablet | Empagliflozin/ Metformin hydrochloride | | 12.500mg + 1.000gram | |  |
| 10701641000033100 | 30982411000001100 | Bilxona 30mg modified-release tablets (Accord-UK Ltd) | Bilxona 30mg modified-release tablets | | | | Modified-release tablet | Gliclazide | | 30.000mg | |  |
| 10701741000033100 | 37618711000001104 | Bilxona 60mg modified-release tablets (Accord-UK Ltd) | Bilxona 60mg modified-release tablets | | | | Modified-release tablet | Gliclazide | | 60.000mg | |  |
| 10951041000033100 | 31015711000001100 | Albiglutide 30mg powder and solvent for solution for injection pre-filled disposable devices | Albiglutide 30mg powder and solvent for solution for injection pre-filled disposable devices | | | | Powder and solvent for solution for injection | Albiglutide | | 30.000mg | | 6010203 |
| 10951141000033100 | 31015811000001100 | Albiglutide 50mg powder and solvent for solution for injection pre-filled disposable devices | Albiglutide 50mg powder and solvent for solution for injection pre-filled disposable devices | | | | Powder and solvent for solution for injection | Albiglutide | | 50.000mg | | 6010203 |
| 11781341000033100 | 33550811000001100 | Metformin 1g/5ml oral solution sugar free | Metformin 1g/5ml oral solution sugar free | | | | Oral solution | Metformin hydrochloride | | 200.000mg/1.000ml | | 6010202 |
| 11781441000033100 | 33550911000001100 | Metformin 850mg/5ml oral solution sugar free | Metformin 850mg/5ml oral solution sugar free | | | | Oral solution | Metformin hydrochloride | | 170.000mg/1.000ml | | 6010202 |
| 11898041000033100 | 33745311000001100 | Saxagliptin 5mg / Dapagliflozin 10mg tablets | Saxagliptin 5mg / Dapagliflozin 10mg tablets | | | | Tablet/ Oral Tablet | Dapagliflozin propanediol monohydrate/ Saxagliptin hydrochloride | | 10.000mg + 5.000mg | | 6010203 |
| 11898141000033100 | 33682311000001100 | Qtern 5mg/10mg tablets (AstraZeneca UK Ltd) | Qtern 5mg/10mg tablets | | | | Tablet/ Oral Tablet | Dapagliflozin propanediol monohydrate/ Saxagliptin hydrochloride | | 10.000mg + 5.000mg | |  |
| 11918941000033100 | 33766211000001100 | Zicron PR 30mg tablets (Bristol Laboratories Ltd) | Zicron PR 30mg tablets | | | | Modified-release tablet | Gliclazide | | 30.000mg | |  |
| 11919041000033100 | 33747711000001100 | Saxenda 6mg/ml solution for injection 3ml pre-filled pens (Novo Nordisk Ltd) | Saxenda 6mg/ml solution for injection 3ml pre-filled pens | | | | Solution for injection | Liraglutide | | 6.000mg/1.000ml | |  |
| 12326541000033100 | 34552411000001100 | Meijumet 500mg modified-release tablets (Medreich Plc) | Meijumet 500mg modified-release tablets | | | | Modified-release tablet | Metformin hydrochloride | | 500.000mg | |  |
| 12326641000033100 | 34552811000001100 | Meijumet 750mg modified-release tablets (Medreich Plc) | Meijumet 750mg modified-release tablets | | | | Modified-release tablet | Metformin hydrochloride | | 750.000mg | |  |
| 12326741000033100 | 34553311000001100 | Meijumet 1000mg modified-release tablets (Medreich Plc) | Meijumet 1000mg modified-release tablets | | | | Modified-release tablet | Metformin hydrochloride | | 1.000gram | |  |
| 12593141000033100 | 35547511000001100 | Yaltormin SR 500mg tablets (Wockhardt UK Ltd) | Yaltormin SR 500mg tablets | | | | Modified-release tablet | Metformin hydrochloride | | 500.000mg | |  |
| 12593241000033100 | 35548011000001100 | Yaltormin SR 750mg tablets (Wockhardt UK Ltd) | Yaltormin SR 750mg tablets | | | | Modified-release tablet | Metformin hydrochloride | | 750.000mg | |  |
| 12593341000033100 | 35548311000001100 | Yaltormin SR 1000mg tablets (Wockhardt UK Ltd) | Yaltormin SR 1000mg tablets | | | | Modified-release tablet | Metformin hydrochloride | | 1.000gram | |  |
| 12664441000033100 | 35849011000001100 | Metuxtan SR 500mg tablets (Accord-UK Ltd) | Metuxtan SR 500mg tablets | | | | Modified-release tablet | Metformin hydrochloride | | 500.000mg | |  |
| 12880941000033100 | 36529511000001104 | Ertugliflozin 5mg tablets | Ertugliflozin 5mg tablets | | | | Tablet/ Oral Tablet | Ertugliflozin L-pyroglutamic acid | | 5.000mg | | 6010203 |
| 12881041000033100 | 36529411000001104 | Ertugliflozin 15mg tablets | Ertugliflozin 15mg tablets | | | | Tablet/ Oral Tablet | Ertugliflozin L-pyroglutamic acid | | 15.000mg | | 6010203 |
| 12881141000033100 | 36514811000001104 | Steglatro 5mg tablets (Merck Sharp & Dohme (UK) Ltd) | Steglatro 5mg tablets | | | | Tablet/ Oral Tablet | Ertugliflozin L-pyroglutamic acid | | 5.000mg | |  |
| 12881241000033100 | 36515111000001104 | Steglatro 15mg tablets (Merck Sharp & Dohme (UK) Ltd) | Steglatro 15mg tablets | | | | Tablet/ Oral Tablet | Ertugliflozin L-pyroglutamic acid | | 15.000mg | |  |
| 12902541000033100 | 36620611000001104 | Suliqua 100units/ml / 33micrograms/ml solution for injection 3ml pre-filled SoloStar pens (Sanofi) | Suliqua 100units/ml / 33micrograms/ml solution for injection 3ml pre-filled SoloStar pens | | | | Solution for injection | Insulin glargine/ Lixisenatide | | 100.000unit/1.000ml + 100.000unit/1.000ml + 33.000microgram/1.000ml | | |
| 12902641000033100 | 36618311000001104 | Suliqua 100units/ml / 50micrograms/ml solution for injection 3ml pre-filled SoloStar pens (Sanofi) | Suliqua 100units/ml / 50micrograms/ml solution for injection 3ml pre-filled SoloStar pens | | | | Solution for injection | Insulin glargine/ Lixisenatide | | 100.000unit/1.000ml + 100.000unit/1.000ml + 50.000microgram/1.000ml | | |
| 12902741000033100 | 36630511000001104 | Insulin glargine 100units/ml / Lixisenatide 33micrograms/ml solution for injection 3ml pre-filled disposable devices | Insulin glargine 100units/ml / Lixisenatide 33micrograms/ml solution for injection 3ml pre-filled disposable devices | | | | Solution for injection | Insulin glargine/ Lixisenatide | | 100.000unit/1.000ml + 100.000unit/1.000ml + 33.000microgram/1.000ml | | 6010102 |
| 12902841000033100 | 36630611000001104 | Insulin glargine 100units/ml / Lixisenatide 50micrograms/ml solution for injection 3ml pre-filled disposable devices | Insulin glargine 100units/ml / Lixisenatide 50micrograms/ml solution for injection 3ml pre-filled disposable devices | | | | Solution for injection | Insulin glargine/ Lixisenatide | | 100.000unit/1.000ml + 100.000unit/1.000ml + 50.000microgram/1.000ml | | 6010102 |
| 12998941000033100 | 36914911000001104 | Gliclazide 160mg tablets | Gliclazide 160mg tablets | | | | Tablet/ Oral Tablet | Gliclazide | | 160.000mg | | 6010201 |
| 12999041000033100 | 36910511000001104 | Glydex 160mg tablets (Medreich Plc) | Glydex 160mg tablets | | | | Tablet/ Oral Tablet | Gliclazide | | 160.000mg | |  |
| 13116741000033100 | 37280311000001104 | Empagliflozin 10mg / Linagliptin 5mg tablets | Empagliflozin 10mg / Linagliptin 5mg tablets | | | | Tablet/ Oral Tablet | Empagliflozin/ Linagliptin | | 10.000mg + 5.000mg | | 6010203 |
| 13116841000033100 | 37280511000001104 | Empagliflozin 25mg / Linagliptin 5mg tablets | Empagliflozin 25mg / Linagliptin 5mg tablets | | | | Tablet/ Oral Tablet | Empagliflozin/ Linagliptin | | 25.000mg + 5.000mg | | 6010203 |
| 13116941000033100 | 37225211000001104 | Glyxambi 10mg/5mg tablets (Boehringer Ingelheim Ltd) | Glyxambi 10mg/5mg tablets | | | | Tablet/ Oral Tablet | Empagliflozin/ Linagliptin | | 10.000mg + 5.000mg | |  |
| 13117041000033100 | 37225511000001104 | Glyxambi 25mg/5mg tablets (Boehringer Ingelheim Ltd) | Glyxambi 25mg/5mg tablets | | | | Tablet/ Oral Tablet | Empagliflozin/ Linagliptin | | 25.000mg + 5.000mg | |  |
| 13122541000033100 | 37405911000001104 | Glibenclamide 600micrograms/ml oral suspension sugar free | Glibenclamide 600micrograms/ml oral suspension sugar free | | | | Oral suspension | Glibenclamide | | 600.000microgram/1.000ml | | 6010201 |
| 13122741000033100 | 37337011000001104 | Amglidia 0.6mg/ml oral suspension with 1ml oral syringe (Bioprojet UK Ltd) | Amglidia 0.6mg/ml oral suspension with 1ml oral syringe | | | | Oral suspension | Glibenclamide | | 600.000microgram/1.000ml | |  |
| 13345341000033100 | 38018511000001104 | Lamzarin 60mg modified-release tablets (Key Pharmaceuticals Ltd) | Lamzarin 60mg modified-release tablets | | | | Modified-release tablet | Gliclazide | | 60.000mg | |  |
| 13429041000033100 | 38238111000001104 | Sukkarto SR 750mg tablets (Morningside Healthcare Ltd) | Sukkarto SR 750mg tablets | | | | Modified-release tablet | Metformin hydrochloride | | 750.000mg | |  |
| 13606641000033100 | 38749111000001104 | Glucorex SR 500mg tablets (GlucoRx Ltd) | Glucorex SR 500mg tablets | | | | Modified-release tablet | Metformin hydrochloride | | 500.000mg | |  |
| 13753141000033100 | 39233411000001104 | Dulaglutide 3mg/0.5ml solution for injection pre-filled disposable devices | Dulaglutide 3mg/0.5ml solution for injection pre-filled disposable devices | | | | Solution for injection | Dulaglutide | | 6.000mg/1.000ml | | 6010203 |
| 13753241000033100 | 39233511000001104 | Dulaglutide 4.5mg/0.5ml solution for injection pre-filled disposable devices | Dulaglutide 4.5mg/0.5ml solution for injection pre-filled disposable devices | | | | Solution for injection | Dulaglutide | | 9.000mg/1.000ml | | 6010203 |
| 13753341000033100 | 39232411000001104 | Trulicity 3mg/0.5ml solution for injection pre-filled pens (Eli Lilly and Company Ltd) | Trulicity 3mg/0.5ml solution for injection pre-filled pens | | | | Solution for injection | Dulaglutide | | 6.000mg/1.000ml | |  |
| 13753441000033100 | 39232611000001104 | Trulicity 4.5mg/0.5ml solution for injection pre-filled pens (Eli Lilly and Company Ltd) | Trulicity 4.5mg/0.5ml solution for injection pre-filled pens | | | | Solution for injection | Dulaglutide | | 9.000mg/1.000ml | |  |
| 13803041000033100 | 39461411000001104 | Metformin 1g tablets | Metformin 1g tablets | | | | Tablet/ Oral Tablet | Metformin hydrochloride | | 1.000gram | | 6010202 |
| 13828541000033100 | 39573711000001104 | Axpinet 500mg tablets (GlucoRx Ltd) | Axpinet 500mg tablets | | | | Tablet/ Oral Tablet | Metformin hydrochloride | | 500.000mg | |  |
| 13828641000033100 | 39573911000001104 | Axpinet 850mg tablets (GlucoRx Ltd) | Axpinet 850mg tablets | | | | Tablet/ Oral Tablet | Metformin hydrochloride | | 850.000mg | |  |
| 14018041000033100 | 40564111000001104 | Jesacrin 1000mg modified-release tablets (Key Pharmaceuticals Ltd) | Jesacrin 1000mg modified-release tablets | | | | Modified-release tablet | Metformin hydrochloride | | 1.000gram | |  |
| 14116341000033100 | 41152511000001104 | Tirzepatide 2.5mg/0.5ml solution for injection pre-filled disposable devices | Tirzepatide 2.5mg/0.5ml solution for injection pre-filled disposable devices | | | | Solution for injection | Tirzepatide | | 5.000mg/1.000ml | | 6010203 |
| 14116441000033100 | 41152611000001104 | Tirzepatide 5mg/0.5ml solution for injection pre-filled disposable devices | Tirzepatide 5mg/0.5ml solution for injection pre-filled disposable devices | | | | Solution for injection | Tirzepatide | | 10.000mg/1.000ml | | 6010203 |
| 14116541000033100 | 41126111000001104 | Mounjaro 2.5mg/0.5ml solution for injection pre-filled pens (Eli Lilly and Company Ltd) | Mounjaro 2.5mg/0.5ml solution for injection pre-filled pens | | | | Solution for injection | Tirzepatide | | 5.000mg/1.000ml | |  |
| 14116641000033100 | 41126511000001104 | Mounjaro 5mg/0.5ml solution for injection pre-filled pens (Eli Lilly and Company Ltd) | Mounjaro 5mg/0.5ml solution for injection pre-filled pens | | | | Solution for injection | Tirzepatide | | 10.000mg/1.000ml | |  |
| 14121441000033100 | 41213411000001104 | Sitagliptin 100mg/5ml oral solution sugar free | Sitagliptin 100mg/5ml oral solution sugar free | | | | Oral solution | Sitagliptin hydrochloride | | 20.000mg/1.000ml | | 6010203 |
| 14219541000033100 | 41535511000001104 | Sitagliptin 125mg/5ml oral solution sugar free | Sitagliptin 125mg/5ml oral solution sugar free | | | | Oral solution | Sitagliptin hydrochloride | | 25.000mg/1.000ml | | 6010203 |
| 14256541000033100 | 42385711000001104 | Jesacrin 500mg modified-release tablets (Key Pharmaceuticals Ltd) | Jesacrin 500mg modified-release tablets | | | | Modified-release tablet | Metformin hydrochloride | | 500.000mg | |  |
| 8268441000033110 | 21994711000001100 | Lixisenatide 10micrograms/0.2ml solution for injection 3ml pre-filled disposable devices and Lixisenatide 20micrograms/0.2ml solution for injection 3ml pre-filled disposable devices | Lixisenatide 10micrograms/0.2ml solution for injection 3ml pre-filled disposable devices and Lixisenatide 20micrograms/0.2ml solution for injection 3ml pre-filled disposable devices | | | | Not applicable |  | 6010203 | |  |  |
| 8268641000033110 | 21953711000001100 | Lyxumia 10micrograms/20micrograms treatment initiation pack (Sanofi) | Lyxumia 10micrograms/20micrograms treatment initiation pack | | | | Not applicable |  | |  | |  |
| 4149341000033110 | 11494211000001100 | Exenatide 5micrograms/0.02ml solution for injection 1.2ml pre-filled disposable devices | Exenatide 5micrograms/0.02ml solution for injection 1.2ml pre-filled disposable devices | | | | Solution for injection | Exenatide | | 250.000microgram/1.000ml | | 6010203 |
| 4149441000033110 | 11494111000001100 | Exenatide 10micrograms/0.04ml solution for injection 2.4ml pre-filled disposable devices | Exenatide 10micrograms/0.04ml solution for injection 2.4ml pre-filled disposable devices | | | | Solution for injection | Exenatide | | 250.000microgram/1.000ml | | 6010203 |
| 4149541000033110 | 11494811000001100 | Byetta 5micrograms/0.02ml solution for injection 1.2ml pre-filled pens (AstraZeneca UK Ltd) | Byetta 5micrograms/0.02ml solution for injection 1.2ml pre-filled pens | | | | Solution for injection | Exenatide | | 250.000microgram/1.000ml | |  |
| 4149641000033110 | 11494611000001100 | Byetta 10micrograms/0.04ml solution for injection 2.4ml pre-filled pens (AstraZeneca UK Ltd) | Byetta 10micrograms/0.04ml solution for injection 2.4ml pre-filled pens | | | | Solution for injection | Exenatide | | 250.000microgram/1.000ml | |  |
| 6388241000033110 | 19275411000001100 | Exenatide 2mg powder and solvent for prolonged-release suspension for injection vials | Exenatide 2mg powder and solvent for prolonged-release suspension for injection vials | | | | Powder and solvent for prolonged-release suspension for inj | Exenatide | | 2.000mg | | 6010203 |
| 6388341000033110 | 19274811000001100 | Bydureon 2mg powder and solvent for prolonged-release suspension for injection vials (AstraZeneca UK Ltd) | Bydureon 2mg powder and solvent for prolonged-release suspension for injection vials | | | | Powder and solvent for prolonged-release suspension for inj | Exenatide | | 2.000mg | |  |
| 13427341000033100 | 28440211000001100 | Exenatide 2mg powder and solvent for prolonged-release suspension for injection pre-filled disposable devices | Exenatide 2mg powder and solvent for prolonged-release suspension for injection pre-filled disposable devices | | | | Powder and solvent for prolonged-release suspension for inj | Exenatide | | 2.000mg | | 6010203 |
| 13427441000033100 | 28426011000001100 | Bydureon 2mg powder and solvent for prolonged-release suspension for injection pre-filled pens (AstraZeneca UK Ltd) | Bydureon 2mg powder and solvent for prolonged-release suspension for injection pre-filled pens | | | | Powder and solvent for prolonged-release suspension for inj | Exenatide | | 2.000mg | |  |
| 13427541000033100 | 38082811000001104 | Exenatide 2mg/0.85ml prolonged-release suspension for injection pre-filled disposable devices | Exenatide 2mg/0.85ml prolonged-release suspension for injection pre-filled disposable devices | | | | Prolonged-release suspension for injection/ | Exenatide | | 2.353mg/1.000ml | | 6010203 |
| 13427641000033100 | 38060511000001104 | Bydureon BCise 2mg/0.85ml prolonged-release suspension for injection pre-filled pens (AstraZeneca UK Ltd) | Bydureon BCise 2mg/0.85ml prolonged-release suspension for injection pre-filled pens | | | | Prolonged-release suspension for injection/ | Exenatide | | 2.353mg/1.000ml | |  |
| 12876141000033100 | 36491111000001104 | Semaglutide 1mg/0.74ml solution for injection 3ml pre-filled disposable devices | Semaglutide 1mg/0.74ml solution for injection 3ml pre-filled disposable devices | | | | Solution for injection | Semaglutide | | 1.340mg/1.000ml | | 6010203 |
| 12876241000033100 | 36490911000001104 | Semaglutide 0.25mg/0.19ml solution for injection 1.5ml pre-filled disposable devices | Semaglutide 0.25mg/0.19ml solution for injection 1.5ml pre-filled disposable devices | | | | Solution for injection | Semaglutide | | 1.340mg/1.000ml | | 6010203 |
| 12876341000033100 | 36491011000001104 | Semaglutide 0.5mg/0.37ml solution for injection 1.5ml pre-filled disposable devices | Semaglutide 0.5mg/0.37ml solution for injection 1.5ml pre-filled disposable devices | | | | Solution for injection | Semaglutide | | 1.340mg/1.000ml | | 6010203 |
| 12876441000033100 | 36471411000001104 | Ozempic 1mg/0.74ml solution for injection 3ml pre-filled pens (Novo Nordisk Ltd) | Ozempic 1mg/0.74ml solution for injection 3ml pre-filled pens | | | | Solution for injection | Semaglutide | | 1.340mg/1.000ml | |  |
| 12876541000033100 | 36470811000001104 | Ozempic 0.25mg/0.19ml solution for injection 1.5ml pre-filled pens (Novo Nordisk Ltd) | Ozempic 0.25mg/0.19ml solution for injection 1.5ml pre-filled pens | | | | Solution for injection | Semaglutide | | 1.340mg/1.000ml | |  |
| 12876641000033100 | 36471111000001104 | Ozempic 0.5mg/0.37ml solution for injection 1.5ml pre-filled pens (Novo Nordisk Ltd) | Ozempic 0.5mg/0.37ml solution for injection 1.5ml pre-filled pens | | | | Solution for injection | Semaglutide | | 1.340mg/1.000ml | |  |
| 13712241000033100 | 38840211000001104 | Semaglutide 3mg tablets | Semaglutide 3mg tablets | | | | Tablet/ Oral Tablet | Semaglutide | | 3.000mg | | 6010203 |
| 13712341000033100 | 38840311000001104 | Semaglutide 7mg tablets | Semaglutide 7mg tablets | | | | Tablet/ Oral Tablet | Semaglutide | | 7.000mg | | 6010203 |
| 13712441000033100 | 38840111000001104 | Semaglutide 14mg tablets | Semaglutide 14mg tablets | | | | Tablet/ Oral Tablet | Semaglutide | | 14.000mg | | 6010203 |
| 13712541000033100 | 38731811000001104 | Rybelsus 3mg tablets (Novo Nordisk Ltd) | Rybelsus 3mg tablets | | | | Tablet/ Oral Tablet | Semaglutide | | 3.000mg | |  |
| 13712641000033100 | 38732111000001104 | Rybelsus 7mg tablets (Novo Nordisk Ltd) | Rybelsus 7mg tablets | | | | Tablet/ Oral Tablet | Semaglutide | | 7.000mg | |  |
| 13712741000033100 | 38732411000001104 | Rybelsus 14mg tablets (Novo Nordisk Ltd) | Rybelsus 14mg tablets | | | | Tablet/ Oral Tablet | Semaglutide | | 14.000mg | |  |
| 14237541000033100 | 42237311000001104 | Semaglutide 0.25mg/0.37ml solution for injection 1.5ml pre-filled disposable devices | Semaglutide 0.25mg/0.37ml solution for injection 1.5ml pre-filled disposable devices | | | | Solution for injection | Semaglutide | | 0.680mg/1.000ml | | 6010203 |
| 14237641000033100 | 42237411000001104 | Semaglutide 1.7mg/0.75ml solution for injection 3ml pre-filled disposable devices | Semaglutide 1.7mg/0.75ml solution for injection 3ml pre-filled disposable devices | | | | Solution for injection | Semaglutide | | 2.270mg/1.000ml | | 6010203 |
| 14237741000033100 | 42237611000001104 | Semaglutide 2.4mg/0.75ml solution for injection 3ml pre-filled disposable devices | Semaglutide 2.4mg/0.75ml solution for injection 3ml pre-filled disposable devices | | | | Solution for injection | Semaglutide | | 3.200mg/1.000ml | | 6010203 |
| 14237841000033100 | 42220711000001104 | Wegovy FlexTouch 0.25mg/0.37ml solution for injection 1.5ml pre-filled pens (Novo Nordisk Ltd) | Wegovy FlexTouch 0.25mg/0.37ml solution for injection 1.5ml pre-filled pens | | | | Solution for injection | Semaglutide | | 0.680mg/1.000ml | |  |
| 14237941000033100 | 42220411000001104 | Wegovy FlexTouch 0.5mg/0.37ml solution for injection 1.5ml pre-filled pens (Novo Nordisk Ltd) | Wegovy FlexTouch 0.5mg/0.37ml solution for injection 1.5ml pre-filled pens | | | | Solution for injection | Semaglutide | | 1.340mg/1.000ml | |  |
| 14238041000033100 | 42219111000001104 | Wegovy FlexTouch 1.7mg/0.75ml solution for injection 3ml pre-filled pens (Novo Nordisk Ltd) | Wegovy FlexTouch 1.7mg/0.75ml solution for injection 3ml pre-filled pens | | | | Solution for injection | Semaglutide | | 2.270mg/1.000ml | |  |
| 14238141000033100 | 42221311000001104 | Wegovy FlexTouch 1mg/0.75ml solution for injection 3ml pre-filled pens (Novo Nordisk Ltd) | Wegovy FlexTouch 1mg/0.75ml solution for injection 3ml pre-filled pens | | | | Solution for injection | Semaglutide | | 1.340mg/1.000ml | |  |
| 14238241000033100 | 42221011000001104 | Wegovy FlexTouch 2.4mg/0.75ml solution for injection 3ml pre-filled pens (Novo Nordisk Ltd) | Wegovy FlexTouch 2.4mg/0.75ml solution for injection 3ml pre-filled pens | | | | Solution for injection | Semaglutide | | 3.200mg/1.000ml | |  |
| 14238441000033100 | 42237511000001104 | Semaglutide 1mg/0.75ml solution for injection 3ml pre-filled disposable devices | Semaglutide 1mg/0.75ml solution for injection 3ml pre-filled disposable devices | | | | Solution for injection | Semaglutide | | 1.340mg/1.000ml | | 6010203 |
| 4522041000033110 | 13626011000001100 | Niddaryl 1mg tablets (Dee Pharmaceuticals Ltd) | Niddaryl 1mg tablets | | | | Tablet | Glimepiride | | 1.000mg | |  |
| 4522141000033110 | 13626511000001100 | Niddaryl 2mg tablets (Dee Pharmaceuticals Ltd) | Niddaryl 2mg tablets | | | | Tablet | Glimepiride | | 2.000mg | |  |
| 4522241000033110 | 13626911000001100 | Niddaryl 3mg tablets (Dee Pharmaceuticals Ltd) | Niddaryl 3mg tablets | | | | Tablet | Glimepiride | | 3.000mg | |  |
| 4522341000033110 | 13627211000001100 | Niddaryl 4mg tablets (Dee Pharmaceuticals Ltd) | Niddaryl 4mg tablets | | | | Tablet | Glimepiride | | 4.000mg | |  |
| 6135341000033110 | 18678711000001100 | Vitile XL 30mg tablets (Actavis UK Ltd) | Vitile XL 30mg tablets | | | | Modified-release tablet | Gliclazide | | 30.000mg | |  |
| 6391241000033110 | 19306911000001100 | Enyglid 1mg tablets (Consilient Health Ltd) | Enyglid 1mg tablets | | | | Tablet | Repaglinide | | 1.000mg | |  |
| 6525941000033110 | 20022911000001100 | Glizofar 15mg tablets (Arrow Generics Ltd) | Glizofar 15mg tablets | | | | Tablet | Pioglitazone hydrochloride | | 15.000mg | |  |
| 6526141000033110 | 20023411000001100 | Glizofar 45mg tablets (Arrow Generics Ltd) | Glizofar 45mg tablets | | | | Tablet | Pioglitazone hydrochloride | | 45.000mg | |  |
| 8839441000033110 | 23372211000001100 | Glidipion 45mg tablets (Actavis UK Ltd) | Glidipion 45mg tablets | | | | Tablet | Pioglitazone hydrochloride | | 45.000mg | |  |
| 10337141000033100 | 29743211000001100 | Diabiom 45mg tablets (Tillomed Laboratories Ltd) | Diabiom 45mg tablets | | | | Tablet | Pioglitazone hydrochloride | | 45.000mg | |  |
| 10952341000033100 | 31014311000001100 | Eperzan 30mg powder and solvent for solution for injection pre-filled pens (GlaxoSmithKline UK Ltd) | Eperzan 30mg powder and solvent for solution for injection pre-filled pens | | | | Powder and solvent for solution for injection | Albiglutide | | 30.000mg | | 6010203 |
| 10953041000033100 | 31014611000001100 | Eperzan 50mg powder and solvent for solution for injection pre-filled pens (GlaxoSmithKline UK Ltd) | Eperzan 50mg powder and solvent for solution for injection pre-filled pens | | | | Powder and solvent for solution for injection | Albiglutide | | 50.000mg | | 6010203 |
| 13122841000033100 | 37337211000001104 | Amglidia 0.6mg/ml oral suspension with 5ml oral syringe (Amring Pharmaceuticals Ltd) | Amglidia 0.6mg/ml oral suspension with 5ml oral syringe | | | | Oral suspension | Glibenclamide | | 600.000microgram/1.000ml | |  |
| 13122941000033100 | 37337511000001104 | Amglidia 6mg/ml oral suspension with 1ml oral syringe (Amring Pharmaceuticals Ltd) | Amglidia 6mg/ml oral suspension with 1ml oral syringe | | | | Oral suspension | Glibenclamide | | 6.000mg/1.000ml | |  |
| 13123041000033100 | 37337711000001104 | Amglidia 6mg/ml oral suspension with 5ml oral syringe (Amring Pharmaceuticals Ltd) | Amglidia 6mg/ml oral suspension with 5ml oral syringe | | | | Oral suspension | Glibenclamide | | 6.000mg/1.000ml | |  |
| 13122641000033100 | 37406011000001104 | Glibenclamide 6mg/ml oral suspension sugar free | Glibenclamide 6mg/ml oral suspension sugar free | | | | Oral suspension | Glibenclamide | | 6.000mg/1.000ml | |  |
| 13345241000033100 | 38018211000001104 | Lamzarin 30mg modified-release tablets (Key Pharmaceuticals Ltd) | Lamzarin 30mg modified-release tablets | | | | Modified-release tablet | Gliclazide | | 30.000mg | |  |
| 12641000033116 |  | Acetohexamide Tablets 500 mg | Acetohexamide Tablets 500 mg | | | | |  | |  | |  |
| 646741000033112 |  | Glymidine Tablets 500 mg | Glymidine Tablets 500 mg | | | |  |  | |  | |  |
| 654041000033116 |  | Grenamide Tablets 2.5 mg | Grenamide Tablets 2.5 mg | | | |  |  | |  | |  |
| 654141000033117 |  | Grenamide Tablets 5 mg | Grenamide Tablets 5 mg | | | |  |  | |  | |  |
| 1452841000033110 |  | Tolazamide Tablets 250 mg | Tolazamide Tablets 250 mg | | | |  |  | |  | |  |
| 1472141000033110 |  | Troglitazone Tablets 200 mg | Troglitazone Tablets 200 mg | | | |  |  | |  | |  |
| 1472241000033110 |  | Troglitazone Tablets 300 mg | Troglitazone Tablets 300 mg | | | |  |  | |  | |  |
| 4269841000033110 |  | Vildagliptin Tablets 100 mg | Vildagliptin Tablets 100 mg | | | |  |  | |  | |  |
| 3890941000033110 |  | Metformin Hydrochloride Sugar free suspension 500 mg/5 ml | Metformin Hydrochloride Sugar free suspension 500 mg/5 ml | | | | |  | |  | |  |
| 1472341000033110 |  | Troglitazone Tablets 400 mg | Troglitazone Tablets 400 mg | | | |  |  | |  | |  |
| 1452741000033110 |  | Tolazamide Tablets 100 mg | Tolazamide Tablets 100 mg | | | |  |  | |  | |  |
| 209341000033112 |  | Calabren Tablets 2.5 mg | Calabren Tablets 2.5 mg | | | |  |  | |  | |  |
| 218441000033114 |  | Calabren Tablets 5 mg | Calabren Tablets 5 mg | | | |  |  | |  | |  |
| 463941000033112 |  | Dimelor Tablets 500 mg | Dimelor Tablets 500 mg | | | |  |  | |  | |  |
| 465841000033115 |  | Diabinese Tablets 100 mg | Diabinese Tablets 100 mg | | | |  |  | |  | |  |
| 465941000033111 |  | Diabinese Tablets 250 mg | Diabinese Tablets 250 mg | | | |  |  | |  | |  |
| 645541000033110 |  | Glucamet Tablets 500 mg | Glucamet Tablets 500 mg | | | |  |  | |  | |  |
| 645641000033111 |  | Glucamet Tablets 850 mg | Glucamet Tablets 850 mg | | | |  |  | |  | |  |
| 646241000033118 |  | Glyconon Tablets 500 mg | Glyconon Tablets 500 mg | | | |  |  | |  | |  |
| 837141000033116 |  | Libanil Tablets 5 mg | Libanil Tablets 5 mg | | | |  |  | |  | |  |
| 869841000033114 |  | Malix Tablets 2.5 mg | Malix Tablets 2.5 mg | | | |  |  | |  | |  |
| 1184741000033110 |  | Romozin Tablets 200 mg | Romozin Tablets 200 mg | | | |  |  | |  | |  |
| 5492041000033110 |  | Amaryl M Tablets 2 mg + 500 mg | Amaryl M Tablets 2 mg + 500 mg | | | | |  | |  | |  |
| 8027541000033110 |  | Diamicron M/R tablets 60 mg | Diamicron M/R tablets 60 mg | | | |  |  | |  | |  |
| 8028441000033110 |  | Galvus Met Tablets 850 mg + 50 mg | Galvus Met Tablets 850 mg + 50 mg | | | | |  | |  | |  |
| 5132141000033110 |  | Janumet Tablets 50 mg + 500 mg | Janumet Tablets 50 mg + 500 mg | | | | |  | |  | |  |
| 1136741000033110 |  | Pramidex Tablets 500 mg | Pramidex Tablets 500 mg | | | |  |  | |  | |  |
| 1184841000033110 |  | Romozin Tablets 300 mg | Romozin Tablets 300 mg | | | |  |  | |  | |  |
| 1452341000033110 |  | Tolanase Tablets 250 mg | Tolanase Tablets 250 mg | | | |  |  | |  | |  |
| 5491941000033110 |  | Amaryl M Tablets 1 mg + 250 mg | Amaryl M Tablets 1 mg + 250 mg | | | | |  | |  | |  |
| 646341000033111 |  | Glymese Tablets 250 mg | Glymese Tablets 250 mg | | | |  |  | |  | |  |
| 8028541000033110 |  | Galvus Met Tablets 1 gram + 50 mg | Galvus Met Tablets 1 gram + 50 mg | | | | |  | |  | |  |
| 5128241000033110 |  | Glucophage Tablets 1000 mg | Glucophage Tablets 1000 mg | | | |  |  | |  | |  |
| 4270041000033110 |  | Galvus Tablets 100 mg | Galvus Tablets 100 mg | | | |  |  | |  | |  |
| 1452241000033110 |  | Tolanase Tablets 100 mg | Tolanase Tablets 100 mg | | | |  |  | |  | |  |
| 837041000033115 |  | Libanil Tablets 2.5 mg | Libanil Tablets 2.5 mg | | | |  |  | |  | |  |
| 869941000033118 |  | Malix Tablets 5 mg | Malix Tablets 5 mg | | | |  |  | |  | |  |
| 650141000033114 |  | Gondafon Tablets 500 mg | Gondafon Tablets 500 mg | | | |  |  | |  | |  |
| 1017241000033110 |  | Orabet Tablets 850 mg | Orabet Tablets 850 mg | | | |  |  | |  | |  |
| 1153441000033110 |  | Rastinon Tablets 500 mg | Rastinon Tablets 500 mg | | | |  |  | |  | |  |
| 406741000033116 |  | Daonil Tablets 2.5 mg | Daonil Tablets 2.5 mg | | | |  |  | |  | |  |
| 1017141000033110 |  | Orabet Tablets 500 mg | Orabet Tablets 500 mg | | | |  |  | |  | |  |
| 1184941000033110 |  | Romozin Tablets 400 mg | Romozin Tablets 400 mg | | | |  |  | |  | |  |
| 10042541000033100 |  | Exenatide Prolonged release suspension for injection 2 mg device |  |  |  |  |  |  |  |  |  |  |
| 10042641000033100 |  | Bydureon Prolonged release suspension for injection 2 mg pen | Bydureon Prolonged release suspension for injection 2 mg pen | | | | |  | |  | |  |

**Abbreviations**

CPRD = Clinical Practice Research Datalink.

#### **Supplementary table S4. List of statins DM+D/Product code list included in CPRD prescriptions as validation**

| **ProdCodeId** | **dmdid** | **Term from EMIS** | **ProductName** | **Formulation** | **Route Of Administration** | **DrugSubstanceName** | **SubstanceStrength** | **BNFChapter** |
| --- | --- | --- | --- | --- | --- | --- | --- | --- |
| 91941000033117 | 39695411000001104 | Atorvastatin 10mg tablets | Atorvastatin 10mg tablets | Tablet/ Oral Tablet | Oral | Atorvastatin calcium trihydrate | 10.000mg | 2120000 |
| 92041000033111 | 39733011000001104 | Atorvastatin 20mg tablets | Atorvastatin 20mg tablets | Tablet/ Oral Tablet | Oral | Atorvastatin calcium trihydrate | 20.000mg | 2120000 |
| 92141000033110 | 39733111000001104 | Atorvastatin 40mg tablets | Atorvastatin 40mg tablets | Tablet/ Oral Tablet | Oral | Atorvastatin calcium trihydrate | 40.000mg | 2120000 |
| 233741000033115 | 42373111000001104 | Cerivastatin 100microgram tablets | Cerivastatin 100microgram tablets | Tablet/ Oral Tablet | Oral | Cerivastatin sodium | 100.000microgram | 2120000 |
| 233841000033113 | 42373211000001104 | Cerivastatin 200microgram tablets | Cerivastatin 200microgram tablets | Tablet/ Oral Tablet | Oral | Cerivastatin sodium | 200.000microgram | 2120000 |
| 233941000033117 | 42373311000001104 | Cerivastatin 300microgram tablets | Cerivastatin 300microgram tablets | Tablet/ Oral Tablet | Oral | Cerivastatin sodium | 300.000microgram | 2120000 |
| 578641000033114 | 39693011000001104 | Fluvastatin 20mg capsules | Fluvastatin 20mg capsules | Capsule/ Oral capsule | Oral | Fluvastatin sodium | 20.000mg | 2120000 |
| 578741000033117 | 39693111000001104 | Fluvastatin 40mg capsules | Fluvastatin 40mg capsules | Capsule/ Oral capsule | Oral | Fluvastatin sodium | 40.000mg | 2120000 |
| 819941000033119 | 84811000001104 | Lescol 20mg capsules (Novartis Pharmaceuticals UK Ltd) | Lescol 20mg capsules | Capsule/ Oral capsule | Oral | Fluvastatin sodium | 20.000mg |  |
| 820041000033116 | 409611000001108 | Lescol 40mg capsules (Novartis Pharmaceuticals UK Ltd) | Lescol 40mg capsules | Capsule/ Oral capsule | Oral | Fluvastatin sodium | 40.000mg |  |
| 834841000033118 | 802411000001108 | Lipostat 10mg tablets (Bristol-Myers Squibb Pharmaceuticals Ltd) | Lipostat 10mg tablets | Tablet/ Oral Tablet | Oral | Pravastatin sodium | 10.000mg |  |
| 834941000033114 | 454111000001107 | Lipostat 20mg tablets (Bristol-Myers Squibb Pharmaceuticals Ltd) | Lipostat 20mg tablets | Tablet/ Oral Tablet | Oral | Pravastatin sodium | 20.000mg |  |
| 836241000033113 | 643911000001108 | Lipitor 10mg tablets (Viatris UK Healthcare Ltd) | Lipitor 10mg tablets | Tablet/ Oral Tablet | Oral | Atorvastatin calcium trihydrate | 10.000mg |  |
| 836341000033115 | 232011000001102 | Lipitor 20mg tablets (Viatris UK Healthcare Ltd) | Lipitor 20mg tablets | Tablet/ Oral Tablet | Oral | Atorvastatin calcium trihydrate | 20.000mg |  |
| 836441000033114 | 484211000001108 | Lipitor 40mg tablets (Viatris UK Healthcare Ltd) | Lipitor 40mg tablets | Tablet/ Oral Tablet | Oral | Atorvastatin calcium trihydrate | 40.000mg |  |
| 836541000033110 | 535011000001102 | Lipostat 40mg tablets (Bristol-Myers Squibb Pharmaceuticals Ltd) | Lipostat 40mg tablets | Tablet/ Oral Tablet | Oral | Pravastatin sodium | 40.000mg |  |
| 836641000033111 | 4535911000001100 | Lipobay 100microgram tablets (Bayer Plc) | Lipobay 100microgram tablets | Tablet/ Oral Tablet | Oral | Cerivastatin sodium | 100.000microgram |  |
| 836741000033119 | 4537511000001100 | Lipobay 200microgram tablets (Bayer Plc) | Lipobay 200microgram tablets | Tablet/ Oral Tablet | Oral | Cerivastatin sodium | 200.000microgram |  |
| 836841000033112 | 4566311000001100 | Lipobay 300microgram tablets (Bayer Plc) | Lipobay 300microgram tablets | Tablet/ Oral Tablet | Oral | Cerivastatin sodium | 300.000microgram |  |
| 1130141000033110 | 42379811000001104 | Pravastatin 10mg tablets | Pravastatin 10mg tablets | Tablet/ Oral Tablet | Oral | Pravastatin sodium | 10.000mg | 2120000 |
| 1130241000033110 | 42379911000001104 | Pravastatin 20mg tablets | Pravastatin 20mg tablets | Tablet/ Oral Tablet | Oral | Pravastatin sodium | 20.000mg | 2120000 |
| 1136541000033110 | 42380011000001104 | Pravastatin 40mg tablets | Pravastatin 40mg tablets | Tablet/ Oral Tablet | Oral | Pravastatin sodium | 40.000mg | 2120000 |
| 1336841000033110 | 42381911000001104 | Simvastatin 10mg tablets | Simvastatin 10mg tablets | Tablet/ Oral Tablet | Oral | Simvastatin | 10.000mg | 2120000 |
| 1336941000033110 | 42382111000001104 | Simvastatin 20mg tablets | Simvastatin 20mg tablets | Tablet/ Oral Tablet | Oral | Simvastatin | 20.000mg | 2120000 |
| 1337541000033110 | 42382311000001104 | Simvastatin 40mg tablets | Simvastatin 40mg tablets | Tablet/ Oral Tablet | Oral | Simvastatin | 40.000mg | 2120000 |
| 1562241000033110 | 859611000001107 | Zocor 40mg tablets (Organon Pharma (UK) Ltd) | Zocor 40mg tablets | Tablet/ Oral Tablet | Oral | Simvastatin | 40.000mg |  |
| 1562841000033110 | 108111000001106 | Zocor 10mg tablets (Organon Pharma (UK) Ltd) | Zocor 10mg tablets | Tablet/ Oral Tablet | Oral | Simvastatin | 10.000mg |  |
| 1562941000033110 | 776811000001104 | Zocor 20mg tablets (Organon Pharma (UK) Ltd) | Zocor 20mg tablets | Tablet/ Oral Tablet | Oral | Simvastatin | 20.000mg |  |
| 1916741000033110 | 4538111000001100 | Lipobay 400microgram tablets (Bayer Plc) | Lipobay 400microgram tablets | Tablet/ Oral Tablet | Oral | Cerivastatin sodium | 400.000microgram |  |
| 1916841000033110 | 42373411000001104 | Cerivastatin 400microgram tablets | Cerivastatin 400microgram tablets | Tablet/ Oral Tablet | Oral | Cerivastatin sodium | 400.000microgram | 2120000 |
| 2066241000033110 | 42297711000001104 | Simvastatin 80mg tablets | Simvastatin 80mg tablets | Tablet/ Oral Tablet | Oral | Simvastatin | 80.000mg | 2120000 |
| 2066341000033110 | 113211000001106 | Zocor 80mg tablets (Organon Pharma (UK) Ltd) | Zocor 80mg tablets | Tablet/ Oral Tablet | Oral | Simvastatin | 80.000mg |  |
| 2189041000033110 | 36566411000001104 | Fluvastatin 80mg modified-release tablets | Fluvastatin 80mg modified-release tablets | Modified-release tablet | Oral | Fluvastatin sodium | 80.000mg | 2120000 |
| 2189141000033110 | 378111000001106 | Lescol XL 80mg tablets (Novartis Pharmaceuticals UK Ltd) | Lescol XL 80mg tablets | Modified-release tablet | Oral | Fluvastatin sodium | 80.000mg |  |
| 2261041000033110 | 39733211000001104 | Atorvastatin 80mg tablets | Atorvastatin 80mg tablets | Tablet/ Oral Tablet | Oral | Atorvastatin calcium trihydrate | 80.000mg | 2120000 |
| 2261141000033110 | 756111000001109 | Lipitor 80mg tablets (Viatris UK Healthcare Ltd) | Lipitor 80mg tablets | Tablet/ Oral Tablet | Oral | Atorvastatin calcium trihydrate | 80.000mg |  |
| 2290041000033110 | 42373511000001104 | Cerivastatin 800microgram tablets | Cerivastatin 800microgram tablets | Tablet/ Oral Tablet | Oral | Cerivastatin sodium | 800.000microgram | 2120000 |
| 2290141000033110 | 4568511000001100 | Lipobay 800microgram tablets (Bayer Plc) | Lipobay 800microgram tablets | Tablet/ Oral Tablet | Oral | Cerivastatin sodium | 800.000microgram |  |
| 2891541000033110 | 42297211000001104 | Rosuvastatin 10mg tablets | Rosuvastatin 10mg tablets | Tablet/ Oral Tablet | Oral | Rosuvastatin calcium | 10.000mg | 2120000 |
| 2891641000033110 | 42297311000001104 | Rosuvastatin 20mg tablets | Rosuvastatin 20mg tablets | Tablet/ Oral Tablet | Oral | Rosuvastatin calcium | 20.000mg | 2120000 |
| 2891741000033110 | 42297411000001104 | Rosuvastatin 40mg tablets | Rosuvastatin 40mg tablets | Tablet/ Oral Tablet | Oral | Rosuvastatin calcium | 40.000mg | 2120000 |
| 2891841000033110 | 4171011000001100 | Crestor 10mg tablets (AstraZeneca UK Ltd) | Crestor 10mg tablets | Tablet/ Oral Tablet | Oral | Rosuvastatin calcium | 10.000mg |  |
| 2891941000033110 | 4171311000001100 | Crestor 20mg tablets (AstraZeneca UK Ltd) | Crestor 20mg tablets | Tablet/ Oral Tablet | Oral | Rosuvastatin calcium | 20.000mg |  |
| 2892041000033110 | 4172111000001100 | Crestor 40mg tablets (AstraZeneca UK Ltd) | Crestor 40mg tablets | Tablet/ Oral Tablet | Oral | Rosuvastatin calcium | 40.000mg |  |
| 2973941000033110 | 4896211000001100 | Simvador 10mg tablets (Dexcel-Pharma Ltd) | Simvador 10mg tablets | Tablet/ Oral Tablet | Oral | Simvastatin | 10.000mg |  |
| 2974041000033110 | 4896511000001100 | Simvador 20mg tablets (Dexcel-Pharma Ltd) | Simvador 20mg tablets | Tablet/ Oral Tablet | Oral | Simvastatin | 20.000mg |  |
| 2974141000033110 | 4896711000001100 | Simvador 40mg tablets (Dexcel-Pharma Ltd) | Simvador 40mg tablets | Tablet/ Oral Tablet | Oral | Simvastatin | 40.000mg |  |
| 3140841000033110 | 7630211000001100 | Ranzolont 10mg tablets (Ranbaxy (UK) Ltd) | Ranzolont 10mg tablets | Tablet/ Oral Tablet | Oral | Simvastatin | 10.000mg |  |
| 3140941000033110 | 7631911000001100 | Ranzolont 20mg tablets (Ranbaxy (UK) Ltd) | Ranzolont 20mg tablets | Tablet/ Oral Tablet | Oral | Simvastatin | 20.000mg |  |
| 3141041000033110 | 7632811000001100 | Ranzolont 40mg tablets (Ranbaxy (UK) Ltd) | Ranzolont 40mg tablets | Tablet/ Oral Tablet | Oral | Simvastatin | 40.000mg |  |
| 3292441000033110 | 42382011000001104 | Simvastatin 20mg / Ezetimibe 10mg tablets | Simvastatin 20mg / Ezetimibe 10mg tablets | Tablet/ Oral Tablet | Oral | Ezetimibe/ Simvastatin | 10.000mg + 20.000mg | 2120000 |
| 3292541000033110 | 42382211000001104 | Simvastatin 40mg / Ezetimibe 10mg tablets | Simvastatin 40mg / Ezetimibe 10mg tablets | Tablet/ Oral Tablet | Oral | Ezetimibe/ Simvastatin | 10.000mg + 40.000mg | 2120000 |
| 3292641000033110 | 42297611000001104 | Simvastatin 80mg / Ezetimibe 10mg tablets | Simvastatin 80mg / Ezetimibe 10mg tablets | Tablet/ Oral Tablet | Oral | Ezetimibe/ Simvastatin | 10.000mg + 80.000mg | 2120000 |
| 3292741000033110 | 9309911000001100 | Inegy 10mg/20mg tablets (Organon Pharma (UK) Ltd) | Inegy 10mg/20mg tablets | Tablet/ Oral Tablet | Oral | Ezetimibe/ Simvastatin | 10.000mg + 20.000mg |  |
| 3292841000033110 | 9310311000001100 | Inegy 10mg/40mg tablets (Organon Pharma (UK) Ltd) | Inegy 10mg/40mg tablets | Tablet/ Oral Tablet | Oral | Ezetimibe/ Simvastatin | 10.000mg + 40.000mg |  |
| 3292941000033110 | 9310611000001100 | Inegy 10mg/80mg tablets (Organon Pharma (UK) Ltd) | Inegy 10mg/80mg tablets | Tablet/ Oral Tablet | Oral | Ezetimibe/ Simvastatin | 10.000mg + 80.000mg |  |
| 3304141000033110 | 8722111000001100 | Simvastatin 20mg/5ml oral suspension | Simvastatin 20mg/5ml oral suspension | Oral suspension | Oral | Simvastatin | 4.000mg/1.000ml | 2120000 |
| 3836041000033110 | 42381811000001104 | Rosuvastatin 5mg tablets | Rosuvastatin 5mg tablets | Tablet/ Oral Tablet | Oral | Rosuvastatin calcium | 5.000mg | 2120000 |
| 3836141000033110 | 9747511000001100 | Crestor 5mg tablets (AstraZeneca UK Ltd) | Crestor 5mg tablets | Tablet/ Oral Tablet | Oral | Rosuvastatin calcium | 5.000mg |  |
| 3868741000033110 | 14018311000001100 | Atorvastatin 20mg/5ml oral suspension | Atorvastatin 20mg/5ml oral suspension | Oral suspension | Oral | Atorvastatin calcium trihydrate | 4.000mg/1.000ml | 2120000 |
| 4152341000033110 | 13894411000001100 | Simvastatin 40mg/5ml oral suspension | Simvastatin 40mg/5ml oral suspension | Oral suspension | Oral | Simvastatin | 8.000mg/1.000ml | 2120000 |
| 4941641000033110 | 15158611000001100 | Simvador 80mg tablets (Dexcel-Pharma Ltd) | Simvador 80mg tablets | Tablet/ Oral Tablet | Oral | Simvastatin | 80.000mg |  |
| 5007141000033110 | 15364211000001100 | Luvinsta XL 80mg tablets (Actavis UK Ltd) | Luvinsta XL 80mg tablets | Modified-release tablet | Oral | Fluvastatin sodium | 80.000mg |  |
| 5711041000033110 | 15534511000001100 | Pravastatin 40mg/5ml oral suspension | Pravastatin 40mg/5ml oral suspension | Oral suspension | Oral | Pravastatin sodium | 8.000mg/1.000ml | 2120000 |
| 5711841000033110 | 17282011000001100 | Dorisin XL 80mg tablets (Aspire Pharma Ltd) | Dorisin XL 80mg tablets | Modified-release tablet | Oral | Fluvastatin sodium | 80.000mg |  |
| 5817741000033110 | 17369311000001100 | Simvastatin 20mg/5ml oral suspension sugar free | Simvastatin 20mg/5ml oral suspension sugar free | Oral suspension | Oral | Simvastatin | 4.000mg/1.000ml | 2120000 |
| 5817841000033110 | 17429811000001100 | Simvastatin 40mg/5ml oral suspension sugar free | Simvastatin 40mg/5ml oral suspension sugar free | Oral suspension | Oral | Simvastatin | 8.000mg/1.000ml | 2120000 |
| 5888441000033110 | 13894211000001100 | Simvastatin 20mg/5ml oral solution | Simvastatin 20mg/5ml oral solution | Oral solution | Oral | Simvastatin | 4.000mg/1.000ml | 2120000 |
| 5888541000033110 | 13894311000001100 | Simvastatin 40mg/5ml oral solution | Simvastatin 40mg/5ml oral solution | Oral solution | Oral | Simvastatin | 8.000mg/1.000ml | 2120000 |
| 5897741000033110 | 14158711000001100 | Atorvastatin 10mg/5ml oral suspension | Atorvastatin 10mg/5ml oral suspension | Oral suspension | Oral | Atorvastatin calcium trihydrate | 2.000mg/1.000ml | 2120000 |
| 5897841000033110 | 14158611000001100 | Atorvastatin 10mg/5ml oral solution | Atorvastatin 10mg/5ml oral solution | Oral solution | Oral | Atorvastatin calcium trihydrate | 2.000mg/1.000ml | 2120000 |
| 5990541000033110 | 14018211000001100 | Atorvastatin 20mg/5ml oral solution | Atorvastatin 20mg/5ml oral solution | Oral solution | Oral | Atorvastatin calcium trihydrate | 4.000mg/1.000ml | 2120000 |
| 6468641000033110 | 19722411000001100 | Atorvastatin 10mg chewable tablets sugar free | Atorvastatin 10mg chewable tablets sugar free | Chewable tablet | Oral | Atorvastatin calcium trihydrate | 10.000mg | 2120000 |
| 6468841000033110 | 19722511000001100 | Atorvastatin 20mg chewable tablets sugar free | Atorvastatin 20mg chewable tablets sugar free | Chewable tablet | Oral | Atorvastatin calcium trihydrate | 20.000mg | 2120000 |
| 6469041000033110 | 19719311000001100 | Lipitor 10mg chewable tablets (Viatris UK Healthcare Ltd) | Lipitor 10mg chewable tablets | Chewable tablet | Oral | Atorvastatin calcium trihydrate | 10.000mg |  |
| 6469141000033110 | 19719611000001100 | Lipitor 20mg chewable tablets (Viatris UK Healthcare Ltd) | Lipitor 20mg chewable tablets | Chewable tablet | Oral | Atorvastatin calcium trihydrate | 20.000mg |  |
| 7861441000033110 | 20528511000001100 | Atorvastatin 30mg tablets | Atorvastatin 30mg tablets | Tablet/ Oral Tablet | Oral | Atorvastatin calcium trihydrate | 30.000mg | 2120000 |
| 7861541000033110 | 20528611000001100 | Atorvastatin 60mg tablets | Atorvastatin 60mg tablets | Tablet/ Oral Tablet | Oral | Atorvastatin calcium trihydrate | 60.000mg | 2120000 |
| 8493841000033110 | 14957711000001100 | Pravastatin 5mg/5ml oral solution | Pravastatin 5mg/5ml oral solution | Oral solution | Oral | Pravastatin sodium | 1.000mg/1.000ml | 2120000 |
| 8962441000033110 | 23864311000001100 | Nandovar XL 80mg tablets (Sandoz Ltd) | Nandovar XL 80mg tablets | Modified-release tablet | Oral | Fluvastatin sodium | 80.000mg |  |
| 10618641000033100 | 14957811000001100 | Pravastatin 5mg/5ml oral suspension | Pravastatin 5mg/5ml oral suspension | Oral suspension | Oral | Pravastatin sodium | 1.000mg/1.000ml | 2120000 |
| 11241741000033100 | 32234211000001100 | Fenofibrate 145mg / Simvastatin 20mg tablets | Fenofibrate 145mg / Simvastatin 20mg tablets | Tablet/ Oral Tablet | Oral | Fenofibrate/ Simvastatin | 145.000mg + 20.000mg | 2120000 |
| 11241841000033100 | 32234311000001100 | Fenofibrate 145mg / Simvastatin 40mg tablets | Fenofibrate 145mg / Simvastatin 40mg tablets | Tablet/ Oral Tablet | Oral | Fenofibrate/ Simvastatin | 145.000mg + 40.000mg | 2120000 |
| 11241941000033100 | 32170911000001100 | Cholib 145mg/20mg tablets (Viatris UK Healthcare Ltd) | Cholib 145mg/20mg tablets | Tablet/ Oral Tablet | Oral | Fenofibrate/ Simvastatin | 145.000mg + 20.000mg |  |
| 13860741000033100 | 39832611000001104 | Rosuvastatin 5mg capsules | Rosuvastatin 5mg capsules | Capsule/ Oral capsule | Oral | Rosuvastatin calcium | 5.000mg | 2120000 |
| 13860841000033100 | 39832311000001104 | Rosuvastatin 10mg capsules | Rosuvastatin 10mg capsules | Capsule/ Oral capsule | Oral | Rosuvastatin calcium | 10.000mg | 2120000 |
| 13860941000033100 | 39832411000001104 | Rosuvastatin 20mg capsules | Rosuvastatin 20mg capsules | Capsule/ Oral capsule | Oral | Rosuvastatin calcium | 20.000mg | 2120000 |
| 13861041000033100 | 39832511000001104 | Rosuvastatin 40mg capsules | Rosuvastatin 40mg capsules | Capsule/ Oral capsule | Oral | Rosuvastatin calcium | 40.000mg | 2120000 |
| 14021741000033100 | 40573211000001104 | Atorvastatin 20mg/5ml oral suspension sugar free | Atorvastatin 20mg/5ml oral suspension sugar free | Oral suspension | Oral | Atorvastatin calcium trihydrate | 4.000mg/1.000ml | 2120000 |
| 14079441000033100 | 40917211000001104 | Rosuvastatin 15mg tablets | Rosuvastatin 15mg tablets | Tablet/ Oral Tablet | Oral | Rosuvastatin calcium | 15.000mg | 2120000 |
| 14079541000033100 | 40917311000001104 | Rosuvastatin 30mg tablets | Rosuvastatin 30mg tablets | Tablet/ Oral Tablet | Oral | Rosuvastatin calcium | 30.000mg | 2120000 |

**Abbreviations**

CPRD = Clinical Practice Research Datalink.

#### **Supplementary table S5. Numbers of participants and prescriptions for prescriptions of oral hypoglycemic agents in CPRD**

| **Drug Class** | **Drug Name** | **Number of prescriptions** | **Number of participants (CPRD)** |
| --- | --- | --- | --- |
| **Acarbose** | **Acarbose** | 1799 | 75 |
| **DPP-4 inhibitors** | **Sitagliptin** | 413401 | 12288 |
|  | **Linagliptin** | 231159 | 7754 |
|  | **Alogliptin benzoate** | 207177 | 7597 |
|  | **Saxagliptin hydrochloride** | 54678 | 1733 |
|  | **Vildagliptin** | 14693 | 459 |
|  | **Dapagliflozin propanediol monohydrate/ Saxagliptin hydrochloride** | 1271 | 61 |
|  | **Empagliflozin/ Linagliptin** | 191 | 16 |
|  | **Sitagliptin hydrochloride** | 2 | 1 |
| **GLP-1 agonists** | **Dulaglutide** | 83726 | 3693 |
|  | **Liraglutide** | 65569 | 2624 |
|  | **Semaglutide** | 55901 | 3689 |
|  | **Exenatide** | 18577 | 913 |
|  | **Lixisenatide** | 7856 | 424 |
|  | **Insulin degludec/ Liraglutide** | 2428 | 96 |
|  | **Insulin glargine/ Lixisenatide** | 26 | 2 |
| **Meglitinides** | **Repaglinide** | 5894 | 238 |
|  | **Nateglinide** | 1087 | 45 |
| **Metformin** | **Metformin hydrochloride** | 5298667 | 95893 |
|  | **Metformin hydrochloride/ Sitagliptin** | 12419 | 388 |
|  | **Metformin hydrochloride/ Rosiglitazone maleate** | 10706 | 518 |
|  | **Metformin hydrochloride/ Pioglitazone hydrochloride** | 10414 | 315 |
|  | **Metformin hydrochloride/ Vildagliptin** | 9204 | 225 |
|  | **Alogliptin benzoate/ Metformin hydrochloride** | 8486 | 261 |
|  | **Dapagliflozin propanediol monohydrate/ Metformin hydrochloride** | 7033 | 323 |
|  | **Empagliflozin/ Metformin hydrochloride** | 5340 | 249 |
|  | **Linagliptin/ Metformin hydrochloride** | 5095 | 185 |
|  | **Metformin hydrochloride/ Saxagliptin hydrochloride** | 1209 | 45 |
|  | **Canagliflozin hemihydrate/ Metformin hydrochloride** | 651 | 30 |
| **SGLT-2 inhibitors** | **Dapagliflozin propanediol monohydrate** | 232738 | 12933 |
|  | **Empagliflozin** | 181890 | 8890 |
|  | **Canagliflozin hemihydrate** | 74498 | 2709 |
|  | **Ertugliflozin L-pyroglutamic acid** | 928 | 74 |
| **Sulfonylureas** | **Gliclazide** | 1464808 | 32001 |
|  | **Glimepiride** | 54545 | 1246 |
|  | **Glipizide** | 16286 | 497 |
|  | **Tolbutamide** | 3941 | 178 |
|  | **Glibenclamide** | 3615 | 194 |
| **Thiazolidinedione** | **Pioglitazone hydrochloride** | 128496 | 3726 |
|  | **Rosiglitazone maleate** | 11142 | 694 |

**Abbreviations**

CPRD = Clinical Practice Research Datalink; DPP-4 = dipeptidyl peptidase-4; GLP-1 = glucagon-like peptide-1; SGLT-2 = sodium-glucose cotransporter-2.

#### **Supplementary table S6. Numbers of participants and prescriptions for prescriptions of statins in the CPRD validation sample**

| **Drug Name** | **Number of prescriptions** | **Number of participants (CPRD)** |
| --- | --- | --- |
| **Atorvastatin calcium trihydrate** | 387175 | 7100 |
| **Simvastatin** | 385750 | 6406 |
| **Pravastatin sodium** | 34547 | 840 |
| **Rosuvastatin calcium** | 33093 | 766 |
| **Fluvastatin sodium** | 4264 | 152 |
| **Ezetimibe/ Simvastatin** | 1251 | 55 |
| **Cerivastatin sodium** | 916 | 92 |
| **Fenofibrate/ Simvastatin** | 11 | 2 |

**Abbreviations**

CPRD = Clinical Practice Research Datalink.

#### **Supplementary table S7. Numbers of participants and prescriptions for antidepressants and antipsychotics in UKB primary care records**

| **Drug Class** | **Drug Name** | | **Number of prescriptions** | **Number of UKB participants** |
| --- | --- | --- | --- | --- |
| **FGA** | **flupentixol** | 10101 | | 1102 |
|  | **prochlorperazine** | 87360 | | 32082 |
|  | **trifluoperazine** | 9207 | | 521 |
|  | **thioridazine** | 5899 | | 659 |
|  | **chlorpromazine** | 4531 | | 318 |
|  | **promethazine** | 3357 | | 527 |
|  | **levomepromazine** | 2326 | | 833 |
|  | **perphenazine** | 1252 | | 57 |
|  | **promazine** | 1215 | | 92 |
|  | **pericyazine** | 417 | | 27 |
|  | **pimozide** | 236 | | 11 |
|  | **sulpiride** | 4867 | | 135 |
|  | **haloperidol** | 3842 | | 742 |
|  | **zuclopenthixol** | 1637 | | 44 |
|  | **benperidol** | 12 | | 1 |
| **MAOI** | **phenelzine** | 3381 | | 71 |
|  | **moclobemide** | 3128 | | 141 |
|  | **tranylcypromine** | 1944 | | 36 |
|  | **isocarboxazid** | 19 | | 4 |
| **MAOI /typical antipsychotic** | **tranylcypromine/trifluoperazine** | 56 | | 2 |
| **NDRI** | **bupropion** | 894 | | 420 |
| **NRI** | **reboxetine** | 5291 | | 325 |
|  | **viloxazine** | 3 | | 1 |
| **SARI** | **trazodone** | 60428 | | 3235 |
|  | **nefazodone** | 3209 | | 313 |
| **SGA** | **flupentixol** | 2694 | | 180 |
|  | **olanzapine** | 41399 | | 923 |
|  | **quetiapine** | 33117 | | 850 |
|  | **risperidone** | 23877 | | 722 |
|  | **aripiprazole** | 8096 | | 275 |
|  | **amisulpride** | 7369 | | 147 |
|  | **fluphenazine** | 499 | | 33 |
|  | **pipotiazine** | 137 | | 5 |
|  | **paliperidone** | 81 | | 5 |
|  | **clozapine** | 29 | | 16 |
|  | **lurasidone** | 23 | | 2 |
|  | **asenapine** | 7 | | 1 |
|  | **zotepine** | 2 | | 1 |
|  | **remoxipride** | 1 | | 1 |
|  | **sertindole** | 1 | | 1 |
| **SNRI** | **venlafaxine** | 170599 | | 5832 |
|  | **duloxetine** | 29206 | | 2312 |
| **SSRI** | **citalopram** | 551339 | | 25516 |
|  | **fluoxetine** | 391439 | | 21705 |
|  | **sertraline** | 238737 | | 12912 |
|  | **paroxetine** | 165293 | | 8410 |
|  | **escitalopram** | 56123 | | 3106 |
|  | **fluvoxamine** | 2227 | | 178 |
|  | **dapoxetine** | 8 | | 3 |
| **TCA** | **amitriptyline** | 600657 | | 44649 |
|  | **dosulepin** | 171937 | | 9751 |
|  | **lofepramine** | 41343 | | 3896 |
|  | **clomipramine** | 26502 | | 950 |
|  | **nortriptyline** | 16761 | | 2030 |
|  | **imipramine** | 16040 | | 950 |
|  | **trimipramine** | 7602 | | 390 |
|  | **doxepin** | 5066 | | 486 |
|  | **protriptyline** | 26 | | 1 |
|  | **amoxapine** | 15 | | 8 |
| **TCA/typical antipsychotic** | **amitriptyline/perphenazine** | 2379 | | 146 |
|  | **nortriptyline/fluphenazine** | 1007 | | 133 |
| **TeCA** | **mianserin** | 827 | | 72 |
|  | **maprotiline** | 343 | | 24 |
| **Amino acid** | **tryptophan** | 471 | | 18 |
| **Antiepileptic** | **carbamazepine** | 71357 | | 2851 |
|  | **valproate** | 52411 | | 1087 |
| **Lithium** | **lithium** | 63285 | | 780 |
| **Other antidepressants** | **mirtazapine** | 133763 | | 7702 |
|  | **agomelatine** | 365 | | 36 |
|  | **vortioxetine** | 16 | | 4 |
| **Tricyclic antipsychotic** | **loxapine** | 158 | | 2 |

**Abbreviations**

FGA = first-generation antipsychotics; SNRI = serotonin-norepinephrine reuptake inhibitors; SSRI = selective serotonin reuptake inhibitors; TCA = tricyclic antidepressants; TeCA = tetracyclic antidepressants; UKB = UK Biobank.

**Supplementary table S8. Number of antidepressant and antipsychotic prescriptions in UKB primary care records, with strength information extracted and imputed by T-Rx**

| **Drug Class** | **Details** | **N** |
| --- | --- | --- |
| **Antidepressants** | Total number of prescriptions | 2,718,545 |
|  | Number of prescriptions with strengths extracted by *`strength_extract()`* function in T-Rx. | 2,320,226 |
|  | Number of prescriptions with multi-ingredient products | 3,442 |
|  | Number of prescriptions with product strengths imputed under *`strength_impute()`* function in T-Rx (multi-ingredient products included) | 398,263 |
| **Antipsychotics / lithium** | Total number of prescriptions | 430,705 |
|  | Number of prescriptions with strengths extracted by *`strength_extract()`* function in T-Rx. | 349,705 |
|  | Number of prescriptions with multi-ingredient products | 0 |
|  | Number of prescriptions with product strengths imputed under *`strength_impute()`* function in T-Rx (multi-ingredient products included) | 81,000 |

**Abbreviations**

UKB = UK Biobank.

**Supplementary table S9. Distribution of strengths of antidepressant prescriptions in UKB primary care records after extraction and imputation by T-Rx**

| Drug Class | Drug Name | Total number of prescriptions | Strength | Number of  prescriptions |
| --- | --- | --- | --- | --- |
| FGA^a^ | **flupentixol** | 10101 | 0.5mg | 35 (0.347%) |
|  |  |  | 1mg | 3361 (33.274%) |
|  |  |  | 500mg | 4 (0.04%) |
|  |  |  | 500mcg | 6701 (66.34%) |
|  | **fluphenazine** | 1007 | 500mcg | 1007 (100%) |
|  | **perphenazine** | 2379 | 2mg | 2379 (100%) |
| MAOI | **isocarboxazid** | 19 | 10mg | 19 (100%) |
|  | **moclobemide** | 3128 | 150mg | 2297 (73.434%) |
|  |  |  | 300mg | 831 (26.566%) |
|  | **phenelzine** | 3381 | 15mg | 3381 (100%) |
|  | **tranylcypromine** | 2000 | 10mg | 2000 (100%) |
| NDRI | **bupropion** | 894 | 150mg | 894 (100%) |
| NRI | **reboxetine** | 5291 | 4mg | 5291 (100%) |
|  | **viloxazine** | 3 | 50mg | 3 (100%) |
| SARI | **nefazodone** | 3209 | 50mg | 6 (0.187%) |
|  |  |  | 100mg | 1376 (42.879%) |
|  |  |  | 200mg | 1827 (56.934%) |
|  | **trazodone** | 60428 | 50mg/5ml | 701 (1.16%) |
|  |  |  | 50mg | 29601 (48.986%) |
|  |  |  | 100mg | 14491 (23.981%) |
|  |  |  | 150mg | 15635 (25.874%) |
| SNRI | **duloxetine** | 29206 | 20mg | 3856 (13.203%) |
|  |  |  | 30mg | 19958 (68.335%) |
|  |  |  | 40mg | 415 (1.421%) |
|  |  |  | 60mg | 4977 (17.041%) |
|  | **venlafaxine** | 170599 | 37.5mg/5ml | 10 (0.006%) |
|  |  |  | 37.5mg | 26680 (15.639%) |
|  |  |  | 50mg | 254 (0.149%) |
|  |  |  | 75mg/5ml | 1 (0.001%) |
|  |  |  | 75mg | 102523 (60.096%) |
|  |  |  | 150mg | 36105 (21.164%) |
|  |  |  | 225mg | 5026 (2.946%) |
| SSRI | **citalopram** | 551339 | 10mg | 117016 (21.224%) |
|  |  |  | 20mg | 383157 (69.496%) |
|  |  |  | 40mg/ml | 725 (0.131%) |
|  |  |  | 40mg | 50441 (9.149%) |
|  | **dapoxetine** | 8 | 30mg | 2 (25%) |
|  |  |  | 60mg | 6 (75%) |
|  | **escitalopram** | 56123 | 5mg | 7111 (12.67%) |
|  |  |  | 10mg/ml | 10 (0.018%) |
|  |  |  | 10mg | 34759 (61.934%) |
|  |  |  | 20mg/ml | 30 (0.053%) |
|  |  |  | 20mg | 14213 (25.325%) |
|  | **fluoxetine** | 391439 | 10mg | 112 (0.029%) |
|  |  |  | 20mg/5ml | 2423 (0.619%) |
|  |  |  | 20mg | 386168 (98.653%) |
|  |  |  | 30mg | 15 (0.004%) |
|  |  |  | 40mg | 35 (0.009%) |
|  |  |  | 60mg | 2686 (0.686%) |
|  | **fluvoxamine** | 2227 | 50mg | 1617 (72.609%) |
|  |  |  | 100mg | 610 (27.391%) |
|  | **paroxetine** | 165293 | 10mg/5ml | 1138 (0.688%) |
|  |  |  | 10mg | 5724 (3.463%) |
|  |  |  | 20mg | 134188 (81.182%) |
|  |  |  | 30mg | 24243 (14.667%) |
|  | **sertraline** | 238737 | 50mg/5ml | 25 (0.01%) |
|  |  |  | 50mg | 155666 (65.204%) |
|  |  |  | 100mg/5ml | 7 (0.003%) |
|  |  |  | 100mg | 83039 (34.783%) |
| TCA | **amitriptyline** | 603036 ^b^ | 10mg/ml | 1 (0%) |
|  |  |  | 10mg/5ml | 306 (0.051%) |
|  |  |  | 10mg | 393241 (65.21%) |
|  |  |  | 25mg/5ml | 52 (0.009%) |
|  |  |  | 25mg | 192568 (31.933%) |
|  |  |  | 50mg/5ml | 9 (0.001%) |
|  |  |  | 50mg | 16805 (2.787%) |
|  |  |  | 75mg | 45 (0.007%) |
|  |  |  | 100mg | 9 (0.001%) |
|  | **amoxapine** | 15 | 25mg | 2 (13.333%) |
|  |  |  | 50mg | 9 (60%) |
|  |  |  | 100mg | 1 (6.667%) |
|  |  |  | 150mg | 3 (20%) |
|  | **clomipramine** | 26502 | 10mg | 6159 (23.24%) |
|  |  |  | 25mg/5ml | 44 (0.166%) |
|  |  |  | 25mg | 12224 (46.125%) |
|  |  |  | 50mg | 6657 (25.119%) |
|  |  |  | 75mg | 1418 (5.351%) |
|  | **dosulepin** | 171937 | 25mg/5ml | 233 (0.136%) |
|  |  |  | 25mg | 99562 (57.906%) |
|  |  |  | 75mg/5ml | 5 (0.003%) |
|  |  |  | 75mg | 72134 (41.954%) |
|  |  |  | 100mg | 2 (0.001%) |
|  |  |  | 125mg | 1 (0.001%) |
|  | **doxepin** | 5066 | 10mg | 628 (12.396%) |
|  |  |  | 25mg | 3105 (61.291%) |
|  |  |  | 50mg | 1153 (22.76%) |
|  |  |  | 75mg | 180 (3.553%) |
|  | **imipramine** | 16040 | 10mg | 651 (4.059%) |
|  |  |  | 25mg/5ml | 1 (0.006%) |
|  |  |  | 25mg | 15387 (95.929%) |
|  |  |  | 100mg | 1 (0.006%) |
|  | **lofepramine** | 41343 | 70mg/5ml | 99 (0.239%) |
|  |  |  | 70mg | 41244 (99.761%) |
|  | **nortriptyline** | 17768 ^a^ | 10mg | 16439 (92.52%) |
|  |  |  | 25mg | 1329 (7.48%) |
|  | **protriptyline** | 26 | 10mg | 26 (100%) |
|  | **trimipramine** | 7602 | 10mg | 774 (10.182%) |
|  |  |  | 25mg | 2509 (33.004%) |
|  |  |  | 50mg | 4319 (56.814%) |
| TeCA | **maprotiline** | 343 | 10mg | 32 (9.329%) |
|  |  |  | 25mg | 76 (22.157%) |
|  |  |  | 50mg | 207 (60.35%) |
|  |  |  | 75mg | 28 (8.163%) |
|  | **mianserin** | 827 | 10mg | 109 (13.18%) |
|  |  |  | 20mg | 74 (8.948%) |
|  |  |  | 30mg | 644 (77.872%) |
| Amino acid | **tryptophan** | 471 | 500mg | 471 (100%) |
| other | **agomelatine** | 365 | 25mg | 365 (100%) |
|  | **mirtazapine** | 133763 | 15mg/ml | 57 (0.043%) |
|  |  |  | 15mg | 36840 (27.541%) |
|  |  |  | 30mg | 68986 (51.573%) |
|  |  |  | 45mg | 27880 (20.843%) |
|  | **vortioxetine** | 16 | 5mg | 2 (12.5%) |
|  |  |  | 10mg | 3 (18.75%) |
|  |  |  | 20mg | 11 (68.75%) |

**Legends**

^a^ As part of multi-strength products.

^b^ 2379 amitriptyline and 1007 nortriptyline prescriptions were expanded from prescriptions with multi-strength products.

**Abbreviations**

FGA = first-generation antipsychotics; SNRI = serotonin-norepinephrine reuptake inhibitors; SSRI = selective serotonin reuptake inhibitors; TCA = tricyclic antidepressants; TeCA = tetracyclic antidepressants; UKB = UK Biobank.

**Supplementary table S10. Distribution of strengths of antipsychotic prescriptions in UKB primary care records after extraction and imputation by T-Rx**

| **Drug Class** | **Drug Name** | **Total number of prescriptions** | **Strength** | **Number of  prescriptions** |
| --- | --- | --- | --- | --- |
| **FGA** | **benperidol** | 12 | 250mcg | 12 (100%) |
|  | **chlorpromazine** | 4531 | 10mg | 2667 (58.861%) |
|  |  |  | 25mg/5ml | 80 (1.766%) |
|  |  |  | 25mg | 1199 (26.462%) |
|  |  |  | 50mg | 404 (8.916%) |
|  |  |  | 100mg/5ml | 2 (0.044%) |
|  |  |  | 100mg | 179 (3.951%) |
|  | **haloperidol** | 3842 | 1mg/ml | 6 (0.156%) |
|  |  |  | 1.5mg | 82 (2.134%) |
|  |  |  | 5mg/5ml | 52 (1.353%) |
|  |  |  | 5mg | 83 (2.16%) |
|  |  |  | 5mg/ml | 458 (11.921%) |
|  |  |  | 10mg/5ml | 1 (0.026%) |
|  |  |  | 10mg | 129 (3.358%) |
|  |  |  | 50mg/ml* | 26 (0.677%) |
|  |  |  | 100mg/ml* | 191 (4.971%) |
|  |  |  | 500mcg | 2814 (73.243%) |
|  | **levomepromazine** | 2326 | 2.5mg/5ml | 4 (0.172%) |
|  |  |  | 6mg | 485 (20.851%) |
|  |  |  | 25mg | 517 (22.227%) |
|  |  |  | 25mg/ml | 1320 (56.75%) |
|  | **pericyazine** | 417 | 2.5mg | 112 (26.859%) |
|  |  |  | 10mg | 305 (73.141%) |
|  | **perphenazine** | 1252 | 2mg | 1219 (97.364%) |
|  |  |  | 4mg | 33 (2.636%) |
|  | **pimozide** | 236 | 2mg | 96 (40.678%) |
|  |  |  | 4mg | 140 (59.322%) |
|  | **prochlorperazine** | 87360 | 3mg | 10194 (11.669%) |
|  |  |  | 5mg/5ml | 100 (0.114%) |
|  |  |  | 5mg | 76277 (87.313%) |
|  |  |  | 12.5mg/ml | 431 (0.493%) |
|  |  |  | 25mg | 358 (0.41%) |
|  | **promazine** | 1215 | 25mg/ml | 70 (5.761%) |
|  |  |  | 25mg/5ml | 301 (24.774%) |
|  |  |  | 25mg | 451 (37.119%) |
|  |  |  | 50mg/5ml | 135 (11.111%) |
|  |  |  | 50mg | 258 (21.235%) |
|  | **promethazine** | 3357 | 10mg | 1 (0.03%) |
|  |  |  | 15mg | 1 (0.03%) |
|  |  |  | 25mg | 3355 (99.94%) |
|  | **sulpiride** | 4867 | 200mg/5ml | 43 (0.884%) |
|  |  |  | 200mg | 4772 (98.048%) |
|  |  |  | 400mg | 52 (1.068%) |
|  | **thioridazine** | 5899 | 10mg | 1795 (30.429%) |
|  |  |  | 25mg/5ml | 48 (0.814%) |
|  |  |  | 25mg | 2652 (44.957%) |
|  |  |  | 50mg | 1101 (18.664%) |
|  |  |  | 100mg | 303 (5.136%) |
|  | **trifluoperazine** | 9207 | 1mg/5ml | 36 (0.391%) |
|  |  |  | 1mg | 4046 (43.945%) |
|  |  |  | 2mg | 916 (9.949%) |
|  |  |  | 5mg/5ml | 404 (4.388%) |
|  |  |  | 5mg | 3525 (38.286%) |
|  |  |  | 10mg | 205 (2.227%) |
|  |  |  | 15mg | 75 (0.815%) |
|  | **zuclopenthixol** | 1637 | 2mg | 718 (43.861%) |
|  |  |  | 10mg | 562 (34.331%) |
|  |  |  | 25mg | 139 (8.491%) |
|  |  |  | 50mg/1ml | 4 (0.244%) |
|  |  |  | 200mg/ml* | 167 (10.202%) |
|  |  |  | 500mg/ml* | 47 (2.871%) |
| **SGA** | **amisulpride** | 7369 | 25mg/5ml | 4 (0.054%) |
|  |  |  | 50mg | 1520 (20.627%) |
|  |  |  | 100mg/ml | 78 (1.058%) |
|  |  |  | 100mg | 618 (8.386%) |
|  |  |  | 200mg | 4554 (61.799%) |
|  |  |  | 400mg | 595 (8.074%) |
|  | **aripiprazole** | 8096 | 1mg/ml | 4 (0.049%) |
|  |  |  | 5mg | 1704 (21.047%) |
|  |  |  | 10mg | 3848 (47.53%) |
|  |  |  | 15mg | 2108 (26.038%) |
|  |  |  | 30mg | 431 (5.324%) |
|  |  |  | 400mg* | 1 (0.012%) |
|  | **asenapine** | 7 | 5mg | 7 (100%) |
|  | **clozapine** | 29 | 25mg | 14 (48.276%) |
|  |  |  | 100mg | 15 (51.724%) |
|  | **flupentixol** | 2694 | 3mg | 1743 (64.699%) |
|  |  |  | 20mg/ml* | 483 (17.929%) |
|  |  |  | 40mg/2ml* | 255 (9.465%) |
|  |  |  | 50mg/0.5ml* | 18 (0.668%) |
|  |  |  | 100mg/ml* | 186 (6.904%) |
|  |  |  | 200mg/ml* | 9 (0.334%) |
|  | **fluphenazine** | 499 | 1mg | 237 (47.495%) |
|  |  |  | 12.5mg/0.5ml* | 12 (2.405%) |
|  |  |  | 25mg/ml* | 232 (46.493%) |
|  |  |  | 50mg/0.5ml* | 2 (0.401%) |
|  |  |  | 50mg/2ml* | 8 (1.603%) |
|  |  |  | 100mg/ml* | 8 (1.603%) |
|  | **lurasidone** | 23 | 18.5mg | 21 (91.304%) |
|  |  |  | 37mg | 1 (4.348%) |
|  |  |  | 74mg | 1 (4.348%) |
|  | **olanzapine** | 41399 | 2.5mg/5ml | 18 (0.043%) |
|  |  |  | 2.5mg | 7298 (17.628%) |
|  |  |  | 5mg | 10388 (25.092%) |
|  |  |  | 7.5mg | 2976 (7.189%) |
|  |  |  | 10mg | 17430 (42.102%) |
|  |  |  | 15mg | 1408 (3.401%) |
|  |  |  | 20mg | 1880 (4.541%) |
|  |  |  | 210mg* | 1 (0.002%) |
|  | **paliperidone** | 81 | 3mg | 2 (2.469%) |
|  |  |  | 75mg* | 54 (66.667%) |
|  |  |  | 100mg/1ml* | 10 (12.346%) |
|  |  |  | 150mg/1.5ml* | 15 (18.519%) |
|  | **pipotiazine** | 137 | 50mg/ml* | 132 (96.35%) |
|  |  |  | 100mg/2ml* | 5 (3.65%) |
|  | **quetiapine** | 33117 | 25mg/5ml | 2 (0.006%) |
|  |  |  | 25mg | 14334 (43.283%) |
|  |  |  | 50mg | 1628 (4.916%) |
|  |  |  | 100mg | 4840 (14.615%) |
|  |  |  | 150mg | 2838 (8.57%) |
|  |  |  | 200mg | 4587 (13.851%) |
|  |  |  | 300mg | 4236 (12.791%) |
|  |  |  | 400mg | 652 (1.969%) |
|  | **remoxipride** | 1 | 150mg | 1 (100%) |
|  | **risperidone** | 23877 | 0.5mg | 9 (0.038%) |
|  |  |  | 1mg/1ml | 24 (0.101%) |
|  |  |  | 1mg/ml | 100 (0.419%) |
|  |  |  | 1mg | 9191 (38.493%) |
|  |  |  | 2mg | 4862 (20.363%) |
|  |  |  | 3mg | 1807 (7.568%) |
|  |  |  | 4mg | 4017 (16.824%) |
|  |  |  | 6mg | 347 (1.453%) |
|  |  |  | 25mg* | 48 (0.201%) |
|  |  |  | 37.5mg* | 68 (0.285%) |
|  |  |  | 50mg* | 100 (0.419%) |
|  |  |  | 500mcg | 3304 (13.838%) |
|  | **sertindole** | 1 | 16mg | 1 (100%) |
|  | **zotepine** | 2 | 25mg | 2 (100%) |
| **antiepileptic** | **carbamazepine** | 71357 | 100mg/5ml | 318 (0.446%) |
|  |  |  | 100mg | 16523 (23.155%) |
|  |  |  | 200mg | 41781 (58.552%) |
|  |  |  | 400mg | 12735 (17.847%) |
|  | **valproate** | 52411 | 100mg | 1416 (2.702%) |
|  |  |  | 150mg | 91 (0.174%) |
|  |  |  | 200mg/5ml | 364 (0.695%) |
|  |  |  | 200mg | 14729 (28.103%) |
|  |  |  | 250mg | 3828 (7.304%) |
|  |  |  | 300mg | 143 (0.273%) |
|  |  |  | 500mg | 31839 (60.749%) |
|  |  |  | 1000mg | 1 (0.002%) |
| **lithium** | **lithium** | 63285 | 1.018mg | 1 (0.002%) |
|  |  |  | 200mg | 18426 (29.116%) |
|  |  |  | 250mg | 2442 (3.859%) |
|  |  |  | 300mg | 43 (0.068%) |
|  |  |  | 400mg | 42007 (66.377%) |
|  |  |  | 450mg | 97 (0.153%) |
|  |  |  | 509mg | 1 (0.002%) |
|  |  |  | 509mg/5ml | 11 (0.017%) |
|  |  |  | 520mg/5ml | 241 (0.381%) |
|  |  |  | 564mg | 13 (0.021%) |
|  |  |  | 800mg | 3 (0.005%) |
| **tricyclic_antipsychotic** | **loxapine** | 158 | 10mg | 153 (96.835%) |
|  |  |  | 25mg | 5 (3.165%) |

* Long-acting formulations for antipsychotics

**Abbreviations**

FGA = first-generation antipsychotics; SGA = second-generation antipsychotics; UKB = UK Biobank.

#### **Supplementary table S11. Comparison table of strength extraction outputs for multi-strength products in CPRD and T-Rx**

| **Name of product (productname)** | **CPRD** | | **T-Rx** | |
| --- | --- | --- | --- | --- |
|  | **Active ingredients listed in CPRD (drugsubstancename)** | **Strengths of active ingredients (substancestrength)** | **Strengths** | **Strength units** |
| Rosiglitazone 1mg / Metformin 500mg tablets | Metformin hydrochloride/ Rosiglitazone maleate | 500.000mg + 1.000mg | 1,500 | mg,mg |
| Rosiglitazone 2mg / Metformin 1g tablets | Metformin hydrochloride/ Rosiglitazone maleate | 1.000gram + 2.000mg | 2,1 | mg,gram |
| Vildagliptin 50mg / Metformin 1g tablets | Metformin hydrochloride/ Vildagliptin | 1.000gram + 50.000mg | 50,1 | mg,gram |
| Alogliptin 12.5mg / Metformin 1g tablets | Alogliptin benzoate/ Metformin hydrochloride | 12.500mg + 1.000gram | 12.5,1 | mg,gram |
| Dapagliflozin 5mg / Metformin 1g tablets | Dapagliflozin propanediol monohydrate/ Metformin hydrochloride | 5.000mg + 1.000gram | 5,1 | mg,gram |
| Pioglitazone 15mg / Metformin 850mg tablets | Metformin hydrochloride/ Pioglitazone hydrochloride | 850.000mg + 15.000mg | 15,850 | mg,mg |
| Canagliflozin 50mg / Metformin 1g tablets | Canagliflozin hemihydrate/ Metformin hydrochloride | 50.000mg + 1.000gram | 50,1 | mg,gram |
| Empagliflozin 12.5mg / Metformin 1g tablets | Empagliflozin/ Metformin hydrochloride | 12.500mg + 1.000gram | 12.5,1 | mg,gram |
| Rosiglitazone 4mg / Metformin 1g tablets | Metformin hydrochloride/ Rosiglitazone maleate | 1.000gram + 4.000mg | 4,1 | mg,gram |
| Saxagliptin 2.5mg / Metformin 1g tablets | Metformin hydrochloride/ Saxagliptin hydrochloride | 1.000gram + 2.500mg | 2.5,1 | mg,gram |
| Rosiglitazone 2mg / Metformin 500mg tablets | Metformin hydrochloride/ Rosiglitazone maleate | 500.000mg + 2.000mg | 2,500 | mg,mg |
| Avandamet 2mg/1000mg tablets | Metformin hydrochloride/ Rosiglitazone maleate | 1.000gram + 2.000mg | 2,1000 | mg,mg |
| Vildagliptin 50mg / Metformin 850mg tablets | Metformin hydrochloride/ Vildagliptin | 850.000mg + 50.000mg | 50,850 | mg,mg |
| Avandamet 2mg/500mg tablets | Metformin hydrochloride/ Rosiglitazone maleate | 500.000mg + 2.000mg | 2,500 | mg,mg |
| Dapagliflozin 5mg / Metformin 850mg tablets | Dapagliflozin propanediol monohydrate/ Metformin hydrochloride | 5.000mg + 850.000mg | 5,850 | mg,mg |
| Saxagliptin 2.5mg / Metformin 850mg tablets | Metformin hydrochloride/ Saxagliptin hydrochloride | 850.000mg + 2.500mg | 2.5,850 | mg,mg |
| Empagliflozin 5mg / Metformin 1g tablets | Empagliflozin/ Metformin hydrochloride | 5.000mg + 1.000gram | 5,1 | mg,gram |
| Linagliptin 2.5mg / Metformin 1g tablets | Linagliptin/ Metformin hydrochloride | 2.500mg + 1000.000mg | 2.5,1 | mg,gram |
| Metformin 1g / Sitagliptin 50mg tablets | Metformin hydrochloride/ Sitagliptin | 1.000gram + 50.000mg | 1,50 | gram,mg |
| Qtern 5mg/10mg tablets | Dapagliflozin propanediol monohydrate/ Saxagliptin hydrochloride | 10.000mg + 5.000mg | 5,10 | mg,mg |
| Xultophy 100units/ml / 3.6mg/ml solution for injection 3ml pre-filled pens | Insulin degludec/ Liraglutide | 100.000unit/1.000ml + 100.000unit/1.000ml + 3.600mg/1.000ml | 100,3.6 | units/ml,mg/1.000ml |
| Xigduo 5mg/1000mg tablets | Dapagliflozin propanediol monohydrate/ Metformin hydrochloride | 5.000mg + 1.000gram | 5,1000 | mg,mg |
| Eucreas 50mg/850mg tablets | Metformin hydrochloride/ Vildagliptin | 850.000mg + 50.000mg | 50,850 | mg,mg |
| Janumet 50mg/1000mg tablets | Metformin hydrochloride/ Sitagliptin | 1.000gram + 50.000mg | 50,1000 | mg,mg |
| Avandamet 4mg/1000mg tablets | Metformin hydrochloride/ Rosiglitazone maleate | 1.000gram + 4.000mg | 4,1000 | mg,mg |
| Suliqua 100units/ml / 33micrograms/ml solution for injection 3ml pre-filled SoloStar pens | Insulin glargine/ Lixisenatide | 100.000unit/1.000ml + 100.000unit/1.000ml + 33.000microgram/1.000ml | 100,33 | units/ml,micrograms/1.000ml |
| Competact 15mg/850mg tablets | Metformin hydrochloride/ Pioglitazone hydrochloride | 850.000mg + 15.000mg | 15,850 | mg,mg |
| Empagliflozin 10mg / Linagliptin 5mg tablets | Empagliflozin/ Linagliptin | 10.000mg + 5.000mg | 10,5 | mg,mg |
| Insulin degludec 100units/ml / Liraglutide 3.6mg/ml solution for injection 3ml pre-filled disposable devices | Insulin degludec/ Liraglutide | 100.000unit/1.000ml + 100.000unit/1.000ml + 3.600mg/1.000ml | 100,3.6 | units/ml,mg/1.000ml |
| Eucreas 50mg/1000mg tablets | Metformin hydrochloride/ Vildagliptin | 1.000gram + 50.000mg | 50,1000 | mg,mg |
| Xigduo 5mg/850mg tablets | Dapagliflozin propanediol monohydrate/ Metformin hydrochloride | 5.000mg + 850.000mg | 5,850 | mg,mg |
| Linagliptin 2.5mg / Metformin 850mg tablets | Linagliptin/ Metformin hydrochloride | 2.500mg + 850.000mg | 2.5,850 | mg,mg |
| Canagliflozin 50mg / Metformin 850mg tablets | Canagliflozin hemihydrate/ Metformin hydrochloride | 50.000mg + 850.000mg | 50,850 | mg,mg |
| Empagliflozin 5mg / Metformin 850mg tablets | Empagliflozin/ Metformin hydrochloride | 5.000mg + 850.000mg | 5,850 | mg,mg |
| Jentadueto 2.5mg/850mg tablets | Linagliptin/ Metformin hydrochloride | 2.500mg + 850.000mg | 2.5,850 | mg,mg |
| Empagliflozin 12.5mg / Metformin 850mg tablets | Empagliflozin/ Metformin hydrochloride | 12.500mg + 850.000mg | 12.5,850 | mg,mg |
| Saxagliptin 5mg / Dapagliflozin 10mg tablets | Dapagliflozin propanediol monohydrate/ Saxagliptin hydrochloride | 10.000mg + 5.000mg | 5,10 | mg,mg |
| Empagliflozin 25mg / Linagliptin 5mg tablets | Empagliflozin/ Linagliptin | 25.000mg + 5.000mg | 25,5 | mg,mg |
| Synjardy 5mg/1000mg tablets | Empagliflozin/ Metformin hydrochloride | 5.000mg + 1.000gram | 5,1000 | mg,mg |
| Jentadueto 2.5mg/1000mg tablets | Linagliptin/ Metformin hydrochloride | 2.500mg + 1000.000mg | 2.5,1000 | mg,mg |
| Komboglyze 2.5mg/1000mg tablets | Metformin hydrochloride/ Saxagliptin hydrochloride | 1.000gram + 2.500mg | 2.5,1000 | mg,mg |
| Synjardy 12.5mg/1000mg tablets | Empagliflozin/ Metformin hydrochloride | 12.500mg + 1.000gram | 12.5,1000 | mg,mg |
| Vipdomet 12.5mg/1000mg tablets | Alogliptin benzoate/ Metformin hydrochloride | 12.500mg + 1.000gram | 12.5,1000 | mg,mg |
| Avandamet 1mg/500mg tablets | Metformin hydrochloride/ Rosiglitazone maleate | 500.000mg + 1.000mg | 1,500 | mg,mg |

**Legends**

For details of prescription details in CPRD Aurum, please refer to CPRD Aurum Data Specification documentation by <https://www.cprd.com/sites/default/files/2024-08/CPRD%20Aurum%20Data%20Specification%20v3.5.pdf>. Sample prescriptions in CPRD Aurum is provided in **Supplementary table 3.**

**Abbreviations**

CPRD = Clinical Practice Research Datalink; mg = millsigrams; ml = millilitres.

#### **Supplementary table S12. Performance metrics for `*strength_impute()*` evaluation in antidepressant prescriptions in UKB**

| Metric | Number of iterations | Proportion of missingness [95% CI] | | |
| --- | --- | --- | --- | --- |
|  |  | **5%** | **10%** | **30%** |
| Accuracy (product strength) | **100** | 0.693 [0.69-0.696] | 0.69 [0.688-0.692] | 0.677 [0.676-0.678] |
|  | **500** | 0.693 [0.691-0.696] | 0.69 [0.688-0.692] | 0.677 [0.677-0.678] |
|  | **750** | 0.693 [0.691-0.696] | 0.69 [0.688-0.692] | 0.677 [0.677-0.678] |
| Accuracy (product strength and units) | **100** | 0.695 [0.692-0.698] | 0.691 [0.689-0.693] | 0.679 [0.678-0.68] |
|  | **500** | 0.694 [0.692-0.697] | 0.691 [0.689-0.693] | 0.679 [0.678-0.68] |
|  | **750** | 0.694 [0.692-0.697] | 0.691 [0.689-0.693] | 0.679 [0.678-0.68] |

**Note**

Accuracy is assessed by the proportion of prescriptions with imputed product strengths and/or units matching that of those extracted by `*strength_extract()*`.

**Abbreviations**

CI = confidence intervals; UKB = UK Biobank.

**Supplementary Figures**

**Supplementary figure S1. Prescriptions of (A) antidepressants; (B) antipsychotics and lithium in UKB primary care records, extracted using READ v2, BNF or dm+d codes**

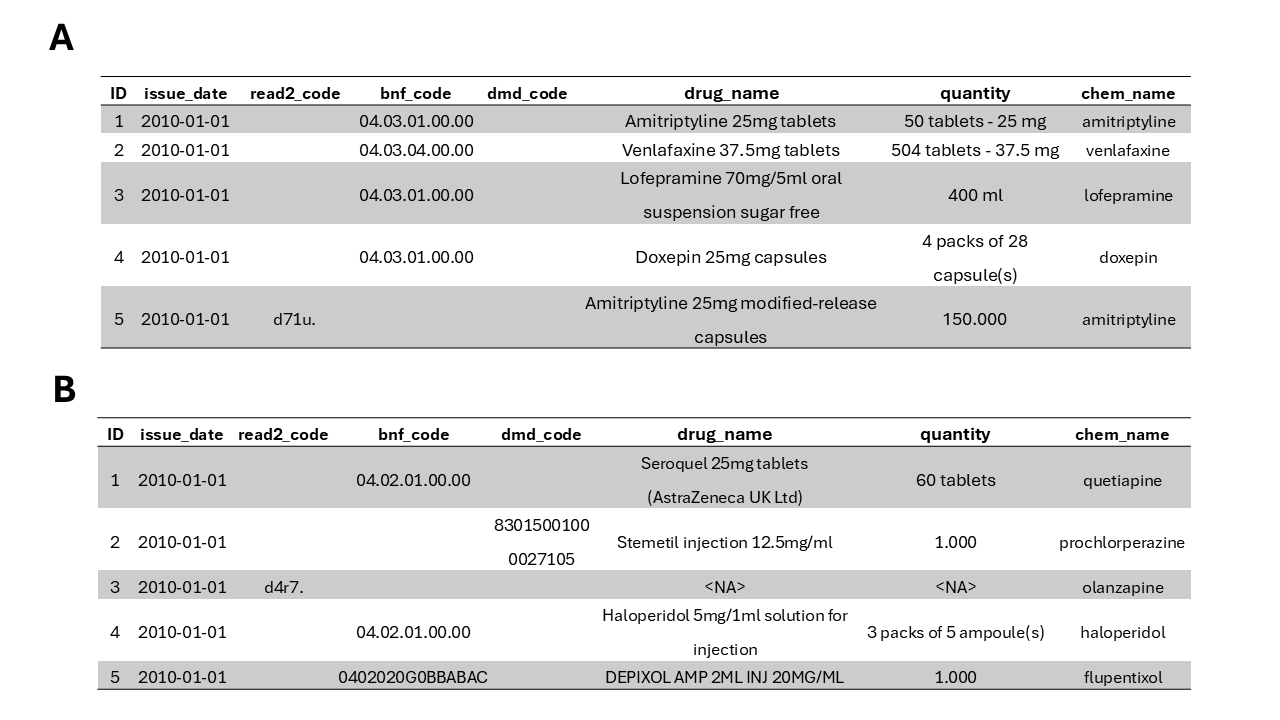

**Abbreviations**

BNF = British National Formulary; dm+d = Dictionary of Medicines and Devices.

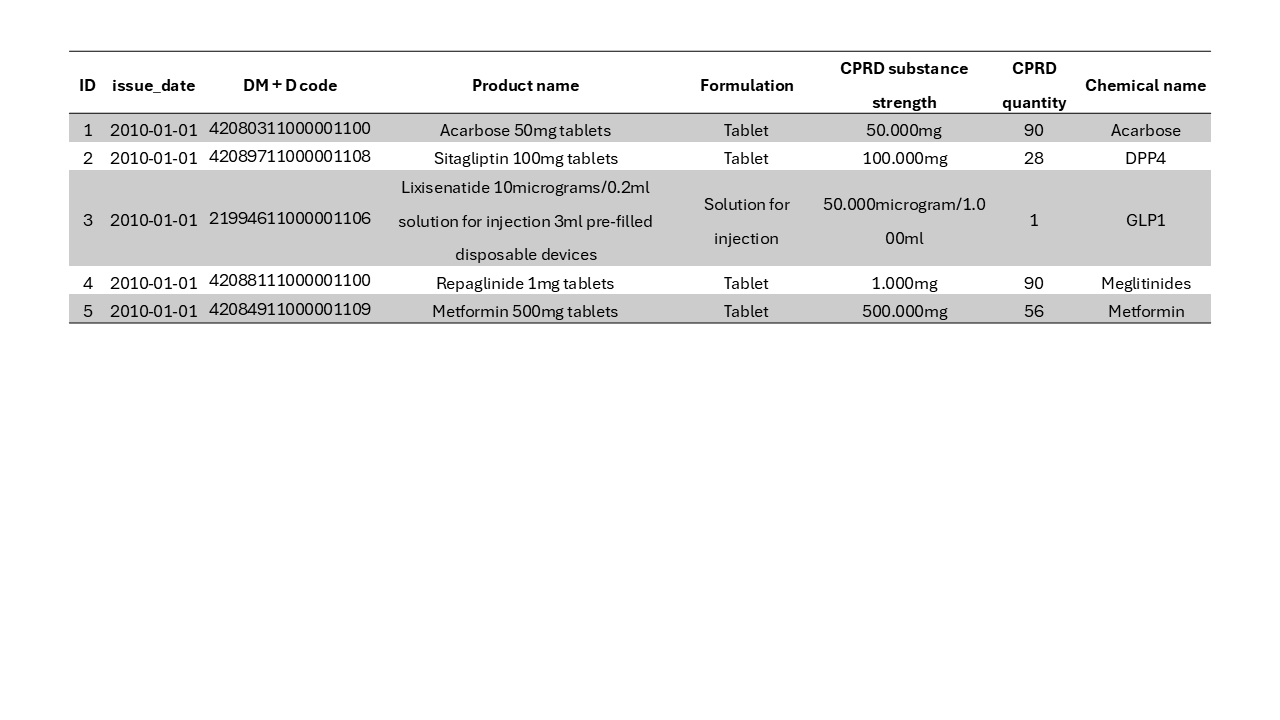
**Supplementary figure S2. Sample prescriptions of oral hypoglycemic agents in CPRD**

**Abbreviations**

CPRD = Clinical Practice Research Datalink; DM+D = Dictionary of Medicines and Devices; DPP-4 = dipeptidyl peptidase-4; GLP-1 = glucagon-like peptide-1.

#### **Supplementary figure S3. Summary of the number of CPRD participants and prescriptions available in CPRD prescription data for oral hypoglycemic agents**

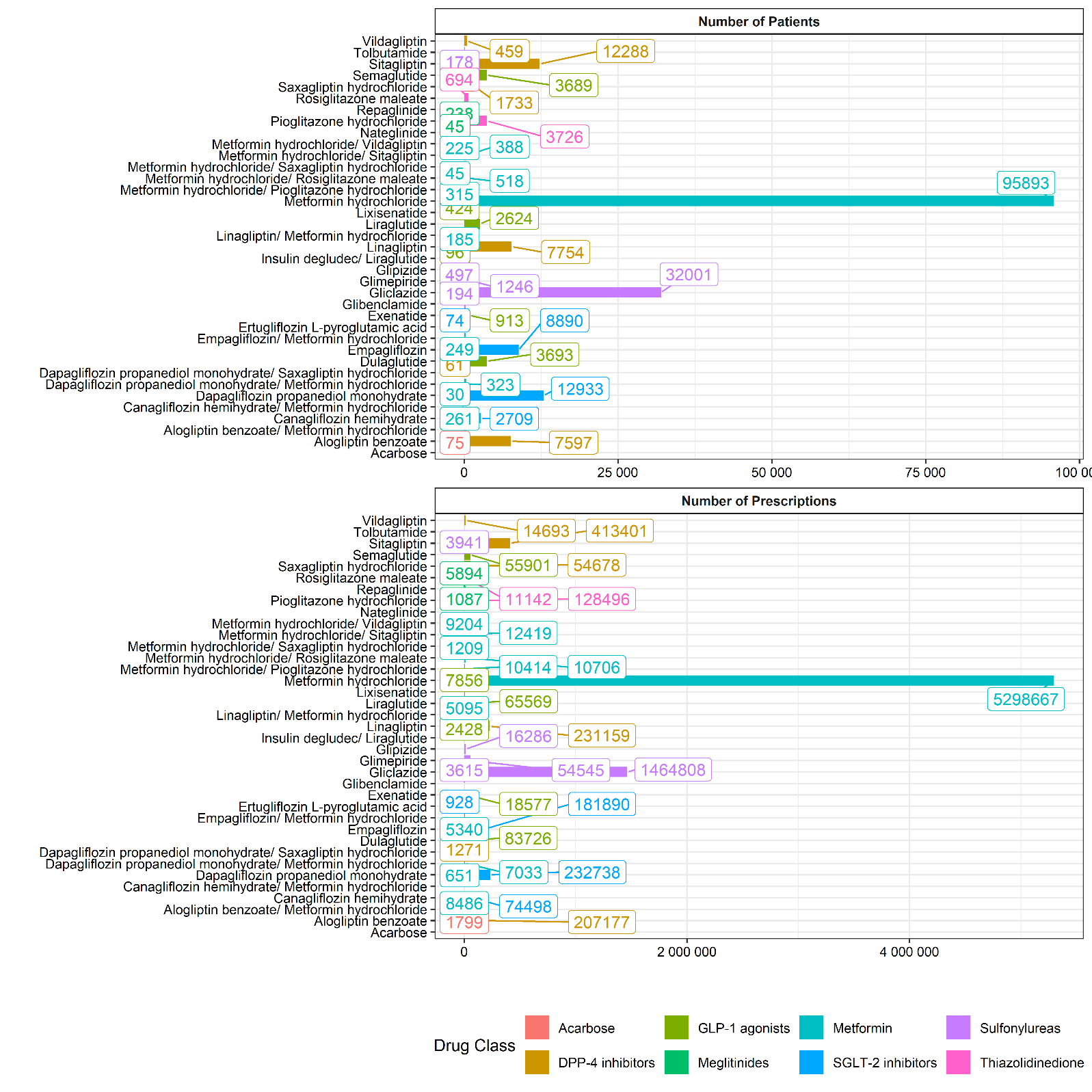

**Abbreviations**

CPRD = Clinical Practice Research Datalink; DPP-4 = dipeptidyl peptidase-4; GLP-1 = glucagon-like peptide-1; SGLT-2 = sodium-glucose cotransporter-2.

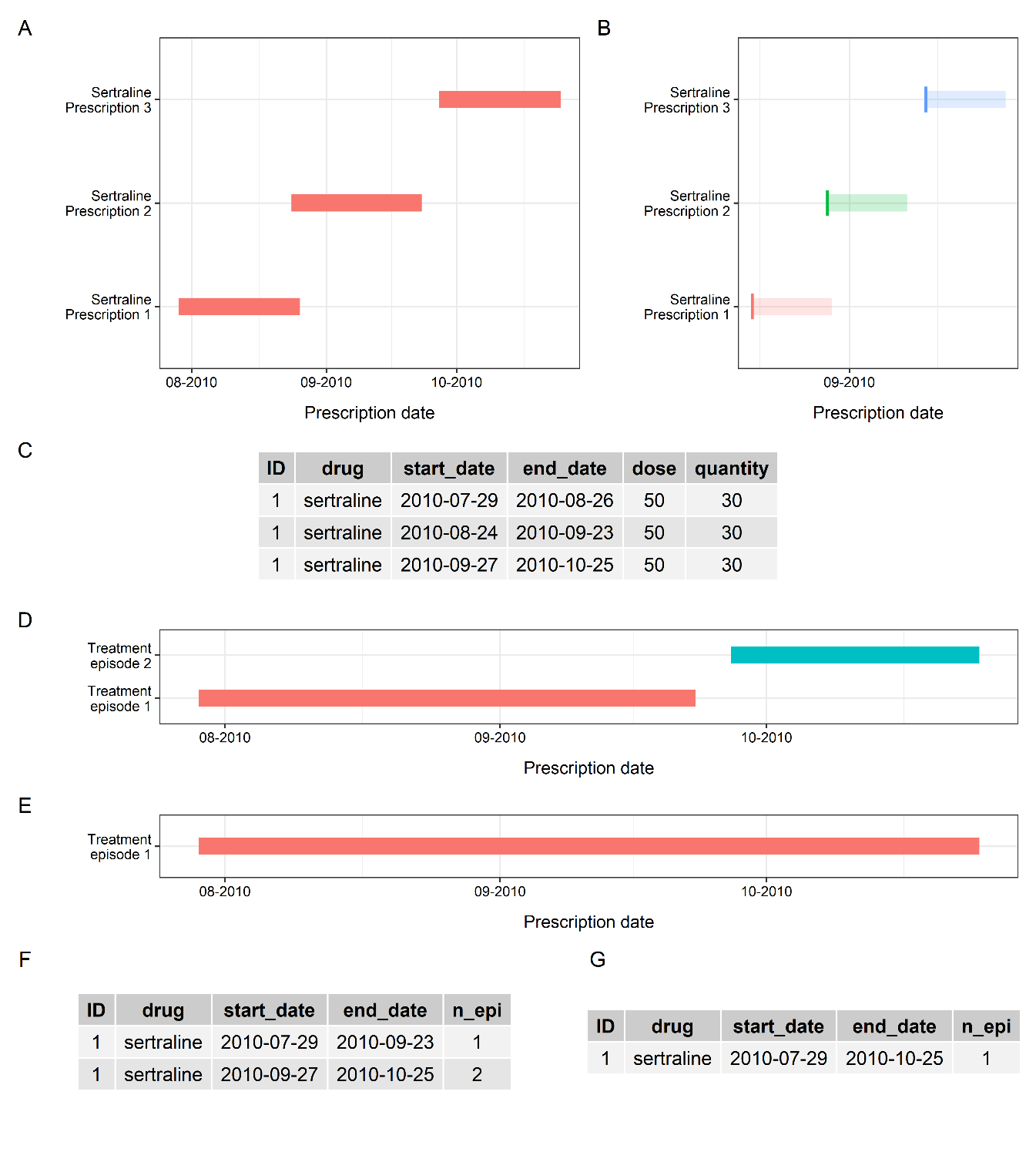

#### **Supplementary figure S4. Exposure ascertainment module functionality for a three-prescription example: (A) sertraline prescriptions (with start and end dates of prescriptions available); (B) sertraline prescriptions (without end dates of prescriptions); (C) sertraline prescriptions (as table); (D) treatment episodes when sertraline prescriptions were merged into prescribing episodes, without gaps allowed between prescriptions; (E) treatment episodes when sertraline prescriptions were merged into prescribing episodes, with a 7-day gap allowed between prescriptions; (F), (G) treatment episodes as tables from (D) and (E), as outputs from *rx_merge()*.**

**Legends**

Details of functions are available on the T-Rx website at: <https://chrislowh.github.io/T-Rx/>.

#### **Supplementary figure S5. Distribution of strengths of (A) antidepressant and (B) antipsychotic prescriptions in UK Biobank primary care records after strength information extracted or imputed by T-Rx. Column labels show strengths and proportions of prescriptions at each strength level.**

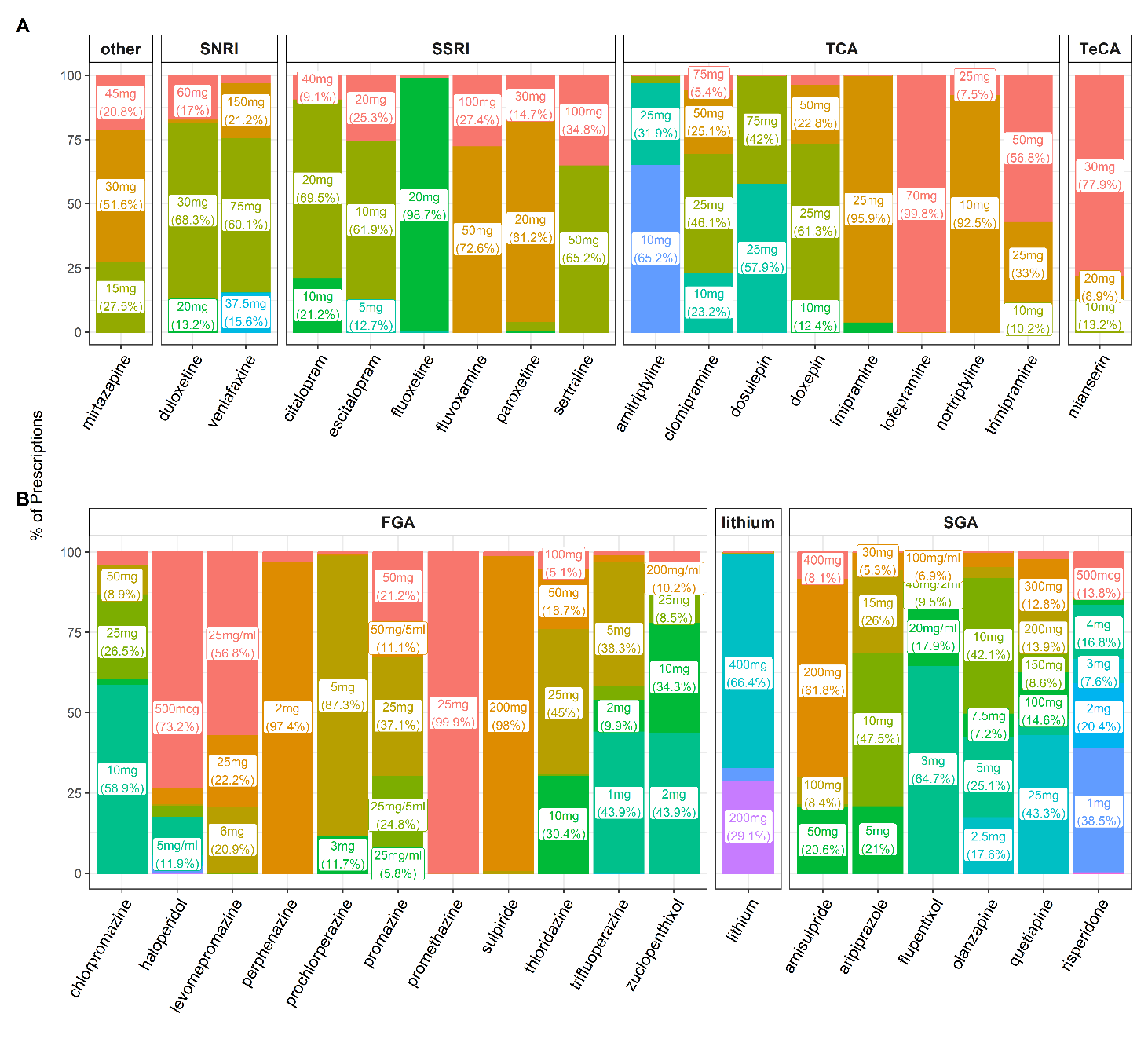

**Legends**

The strength distributions of SSRIs, SNRIs, TCAs, TeCAs and mirtazapine are shown for antidepressants, while those for FGA, SGA and lithium are shown for antipsychotic prescriptions. Only products with more than 500 prescriptions in UKB primary care records were included, with labels only for strengths of products accounting for more than 5% of all products with the same drug name. Details of strength distributions are summarized in **Supplementary tables 9 and 10**.

**Abbreviations**

FGA = first-generation antipsychotics; SGA = second-generation antipsychotics; SNRI = serotonin-norepinephrine reuptake inhibitors; SSRI = selective serotonin reuptake inhibitors; TCA = tricyclic antidepressants; TeCA = tetracyclic antidepressants; UKB = UK Biobank.

#### **Supplementary figure S6. Sankey diagrams to compare the performance of strength extraction functions in T-Rx with (A) oral hypoglycemic agent and (B) statin prescriptions in CPRD**

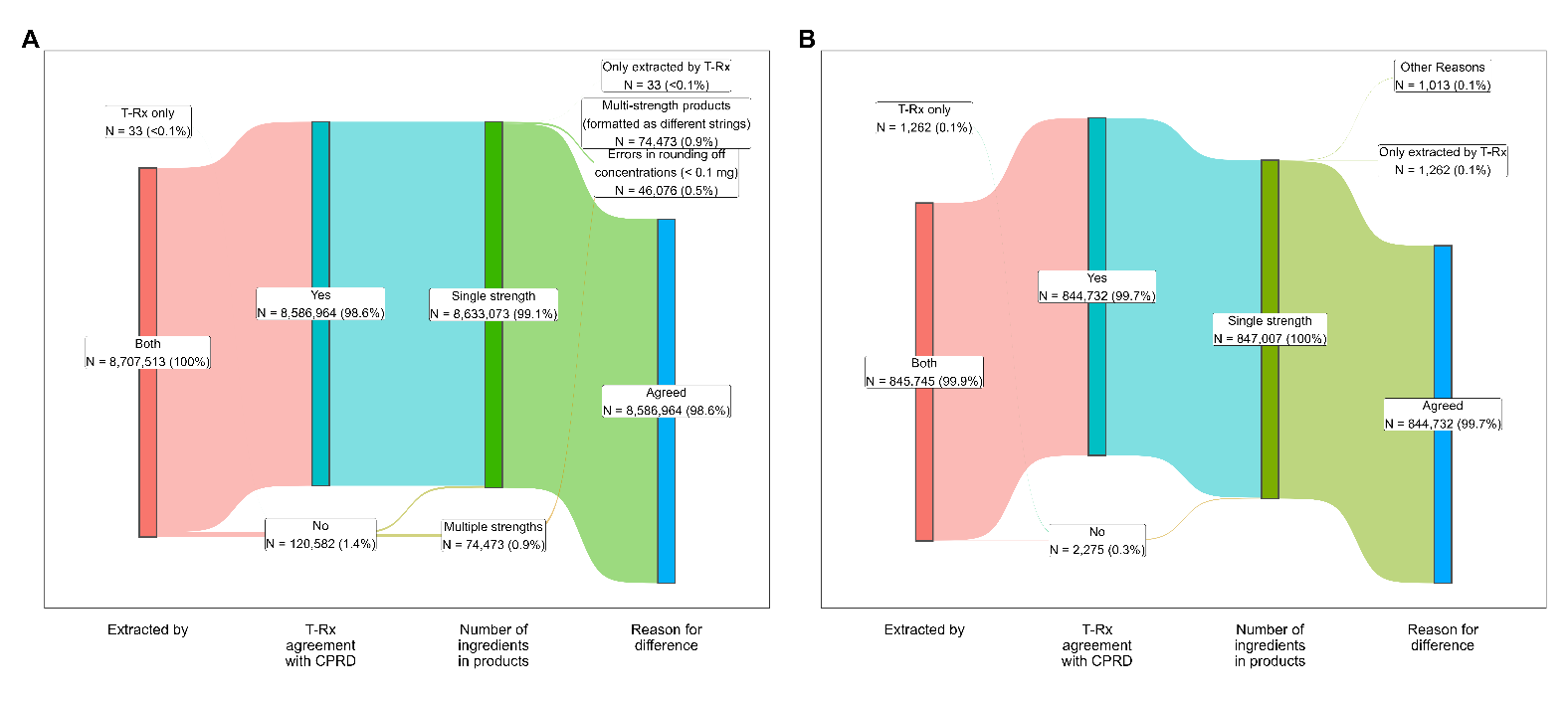

**Legends**

Figures expressed as number of prescriptions (percentage)

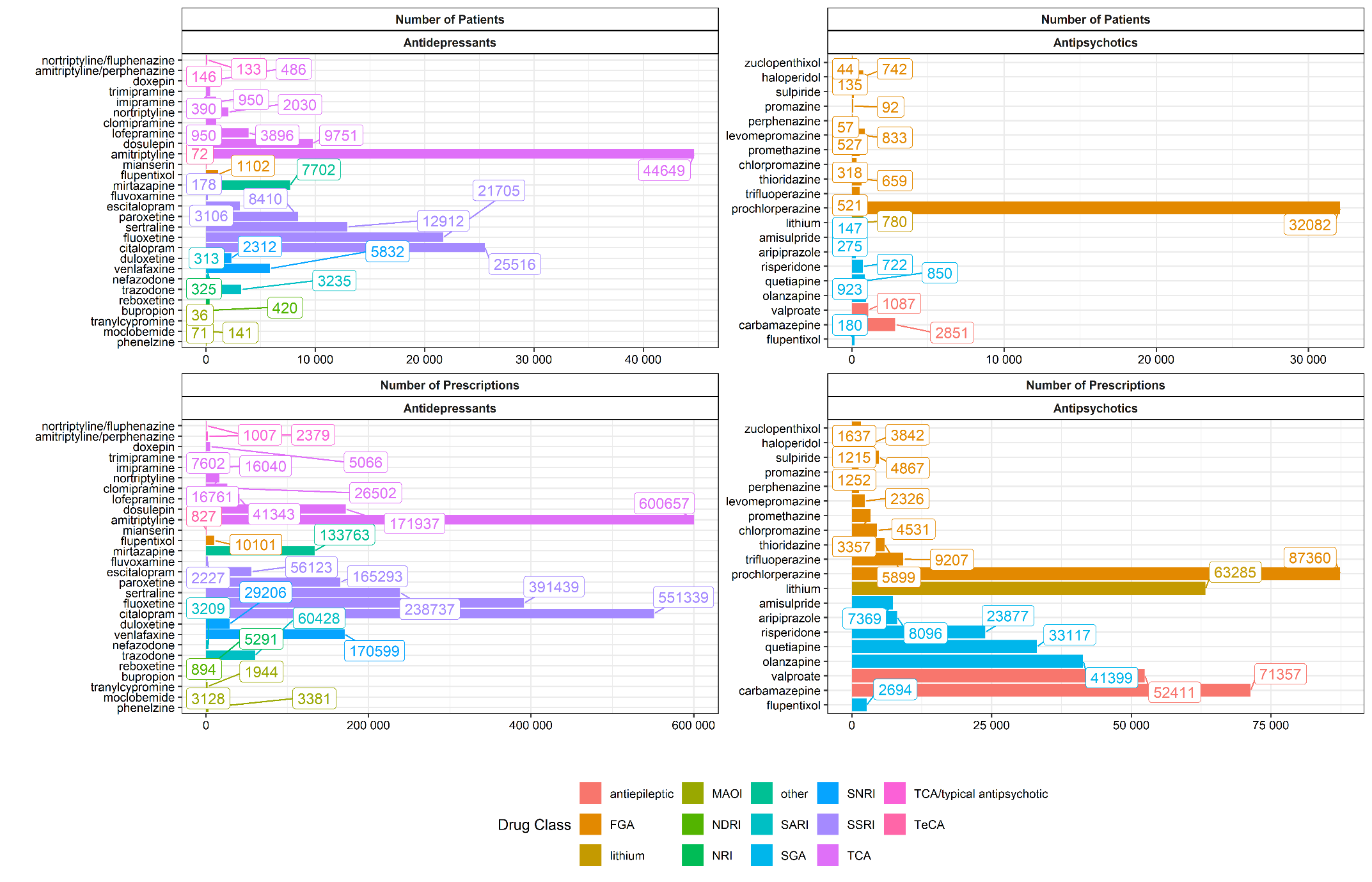

#### **Supplementary figure S7.** **Summary of the number of UK Biobank participants (row 1) and prescriptions (row 2) for antidepressants and antipsychotics/lithium data available in primary care records**

**Legends**

Lithium is illustrated in the panel along with antipsychotic prescriptions for illustration purposes.

**Abbreviations**

FGA = first-generation antipsychotics; MAOI = monoamine oxidase inhibitors; NDRI = norepinephrine-dopamine reuptake inhibitors; NRI = norepinephrine reuptake inhibitors; SARI = serotonin antagonist and reuptake inhibitors; SGA = second-generation antipsychotics; SNRI = serotonin-norepinephrine reuptake inhibitors; SSRI = selective serotonin reuptake inhibitors; TCA = tricyclic antidepressants; TeCA = tetracyclic antidepressants.
